# Forecasting waiting lists for elective procedures and surgery in England: a modelling study

**DOI:** 10.1101/2022.06.20.22276651

**Authors:** Dmitri Nepogodiev, Radhika Acharya, Daoud Chaudhry, James C Glasbey, Benjamin Harris, Elizabeth Li, Kate Jolly, Aneel Bhangu

## Abstract

**Introduction:** The aim of this study was to forecast the total need for elective procedures in England by 2030.

**Methods:** We used publicly available activity data from NHS Digital to estimate procedure-level shortfalls in elective procedures performed during the pandemic period (January 2020 to March 2022) compared to what would be expected based on pre-pandemic trends. We also estimated the procedure-level composition of the NHS waiting list immediately preceding the pandemic (December 2019). The total need for elective procedures in March 2022 was calculated by summing the pandemic shortfall with the pre-pandemic NHS waiting list. We projected the need for elective procedures through to January 2030 for four scenarios: current capacity (surgical volume remains at the same level as in February-March 2022), pessimistic scenario (elective procedure volume increases to pre-pandemic levels by July 2023 followed and remains at this level until 2030), central scenario (elective procedure volume returns to pre-pandemic levels by December 2022 followed by a 2% increase per year), optimistic scenario (elective procedure volume returns to pre-pandemic levels by December 2022 followed by a 4% increase per year)

**Results:** We estimated the total need for elective procedures in England in March 2022 was 4,347,469. Of these 4,347,469 patients, 3,304,513 (76.0%) were on a hidden waiting list. The greatest need was for General Surgery (1,522,366), Orthopaedics (976,875), and Ophthalmology (391,683). The procedures with the greatest need were sigmoidoscopy and colonoscopy (568,838), gastroscopy (447,830), cataract surgery (314,790), lower limb joint replacement (224,363), and interventional cardiology (349,300). We projected that at current capacity, the total number of elective procedures needed would increase to 14,608,195 by 2030. In the pessimistic scenario, elective procedure volume total elective procedures needed would increase to 8,507,087, in the central scenario it would increase to 5,420,999, and in the optimistic scenario it would decrease to 2,584,664 procedures.

**Discussion:** The estimate of 4.3 million elective procedures needed in England is considerably higher than the official NHS waiting list, reflecting a large hidden waiting list. Even in the most optimistic scenarios there will be a substantially larger waiting list in 2030 than pre-pandemic.

## Introduction

Prior to the COVID-19 pandemic NHS elective services in England regularly experienced service disruption due to increasing emergency care pressures, for example, due to winter pressures^1^. This resulted in increasing cancellation rates for elective procedures^2^. During the first COVID-19 wave, the NHS stopped most routine elective surgery, in order to redeploy staff and resources to support the acute COVID-19 response^3–4^. During subsequent waves, the NHS was unable to return to baseline pre-pandemic elective procedure volume due to a combination of stapatient safety concerns and staffing pressures^5–6^.

According to NHS waiting list data, 6.3 million patients were waiting for elective treatment in England in March 2022^7^. However, the waiting list does not fully reflect population need for elective procedures, since fewer new patients were seen in clinic by hospital consultants during the pandemic, as a result of reduced referrals and reduced clinic capacity due to redeployment of resources to support the acute COVID-19 response. This has resulted in a hidden waiting list comprising people who have symptoms or disease requiring elective procedures, but who have not been placed on the elective waiting list^8^.

In February 2022, the UK Government has committed £1.5 billion to financing elective surgery hubs to tackle the elective procedure backlog in England^9^. Whilst there is an ambition to increase surgical capacity above pre-pandemic levels, the full extent, including the hidden waiting list, of current and future need for elective procedures is not known. The aim of this study was to project the total number of patients who will need elective procedures in England by January 2030 based on a series of scenarios.

## Methods

In order to project future need for elective procedures, we first needed to determine baseline need. We estimated the number of patients who needed elective procedures in England in March 2022; this was taken as baseline need because March 2022 was the most recent month for which hospital activity data were available at the time of analysis. We then projected monthly figures for the number of patients who will need elective procedures in England through to 2030.

### Definitions

<u>Elective procedure</u>: this is used as a collective term for surgical operations and endoscopic, interventional cardiology, or interventional radiology procedures. An elective surgical operation was defined as an operation performed by a surgeon in an operating theatre on a planned admission to hospital. This definition is consistent with previous studies^6, 10^. Obstetric operations were excluded as they do not contribute to the main waiting list. Both diagnostic and therapeutic endoscopy, interventional cardiology, and interventional radiology were included if performed in an operating theatre, endoscopy suite, or interventional radiology suite, on a planned admission to hospital. Both day-case procedures and procedures with an overnight admission were included. Minor procedures that are normally performed outside a theatre, endoscopy suite, or interventional radiology suite (e.g. paracentesis, lumbar puncture, joint injection), non-procedural therapeutics (e.g. drug infusion), and non-interventional imaging were excluded. A breakdown of the 1,139 OPCS Classification of Interventions and Procedures codes fulfilling inclusion criteria is provided in Supplementary Table 1.

Calculations were performed at the level of individual OPCS codes, but to aid interpretation, the 1,139 OPCS codes were combined in to 130 procedure categories, which in turn were further summed to 16 sub-specialties and 10 specialties (Supplementary Table 1).

<u>Day-case elective procedure</u>: day-case procedures are completed without an overnight stay in hospital. For this analysis, we classified procedures as day-case or as requiring overnight admission. Using AHES-APC (see below), we reviewed length of hospital stay for each OPCS code in 2018-19 and classified them as day-case if they had an average length of hospital stay under 1 day, or 50% or more of cases were performed as day-cases.

<u>Incident need for elective procedures</u>: The number of new patients each year who develop symptoms or disease that require an elective procedure. The incident need rate is the incident need per 1,000 population.

<u>Pandemic shortfall in elective procedures</u>: the reduction in the number of elective procedures performed during the pandemic period (January 2020 to March 2022) compared to what would be expected based on pre-pandemic trends, adjusted for population growth and ageing.

<u>Total need for elective procedures</u>: the total number elective procedures needed in England, at a given point in time. This count includes all patients regardless of whether or not they are on an NHS waiting list.

<u>NHS waiting list</u>: patients who are on the NHS waiting list for an elective procedure.

<u>Hidden waiting list</u>: patients who need elective procedures, but who have not been added to the NHS waiting list by a hospital consultant for reasons related to the COVID-19 pandemic. This might occur in the following circumstances:

- The patient did not see their general practitioner (GP) and therefore they were not referred to a consultant.
- The patient did see their GP, who decided to not make a referral to a consultant due to factors related to the pandemic.
- The GP did make a referral to a consultant, but the patient has not yet been seen by the consultant due to the pandemic significantly increasing waiting times for clinic.
- The patient did see a hospital consultant, but, due to factors related to the pandemic, have not yet been added to the NHS waiting list.
- However, if a patient was seen by a hospital consultant and was added them to the NHS waiting list, this patient would appear on the NHS waiting list rather than the hidden waiting list.

Based on these definitions, total need was calculated as the sum of the NHS waiting list and hidden waiting list:

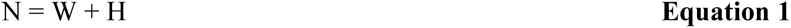

Where

N = Total need for elective procedures at baseline in a given month

W = Number of patients waiting for elective procedures on the NHS waiting list in a given month

H = Hidden waiting list in a given month

### Conceptual framework

#### Key assumptions

This analysis is based on the following assumptions:

- Prior to the COVID-19 pandemic, the NHS waiting list for elective care included all people who needed elective procedures; i.e. there was no hidden waiting list pre-pandemic before the pandemic.
- Age-sex specific incident need rates for elective procedures remained constant during the pandemic period. Consequently, if there was a reduction in elective procedure activity during the pandemic period (pandemic shortfall) this would result in a backlog of patients, increasing total need for elective procedures.
- There is no attrition to individuals’ need for elective procedures over time. This means that all patients who need elective procedures stay on the waiting list regardless of how long they have to wait to have their procedure.
- Age and sex specific incident need rates for elective procedures will remain constant through to 2030. Our modelling takes in to account projected changes in England’s population structure over time, meaning that the number of elective procedures needed each year (incident need) may increase over time, even though we have assumed that age-sex specific incident need rates do not change.

#### Approach to modelling

We projected the number of patients who will need elective procedures in England each month from April 2022 to December 2029. The following calculation was performed for each month:

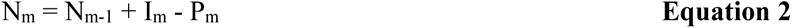

Where

Nm = Total need for elective procedures at the end of month m

Nm-1 = Total need for elective procedures at the end of the month preceding month m

Im = Incident need for elective procedures in month m. This was calculated as one twelfth of annual incident need in the relevant year. Annual incident need was adjusted for projected changes in population structure over time

Pm = Number of elective procedures performed in month m

Baseline total need for elective procedures was estimated for March 2022 based on real-world NHS activity data. Total need in March 2022 was calculated as sum of the pre-pandemic waiting list and the pandemic shortfall in elective procedures:

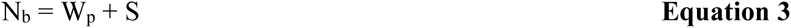

Where

Nb = Total need for elective procedures at baseline in March 2022

Wp = Number of patients on the NHS waiting list for elective procedures immediately before the pandemic in December 2019

S = Pandemic shortfall in elective procedures

### Data sources

This study used the following NHS (National Health Service) England data which are publicly available from NHS Digital:

- Monthly Hospital Episode Statistics for Admitted Patient Care (MHES-APC) activity data for April 2018 to March 2022^11^.
- Annual Hospital Episode Statistics for Admitted Patient Care (AHES-APC) activity data for 2018-19 and 2020-21^11^.
- NHS England waiting list data for March 2015 to March 2022^7^.
- NHS reference costs for 2019-20 (most recent available)^12^.

In addition, the following data were accessed from the Office for National Statistics (ONS):

- Population (age and sex) structure data for mid-2018 to mid-2020^13^.
- Population (age and sex) structure projections for 2021 to 2029^14^.
- Health-system level population data for mid-2020 (most recent available)^15^.

#### Hospital Episode Statistics

Hospital Episode Statistics (HES) captures data for inpatient NHS patient episodes across all NHS and private hospitals in England^11^. HES has monthly (MHES-APC) and annual (AHES- APC) data releases. AHES-APC data are released around September for the preceding NHS year (April to March) and MHES-APC data are released monthly on a rolling basis.

Hospital Episode Statistics (HES) provides information on finished consultant episodes and finished admission episodes. A finished consultant episode is a period of care under a particular consultant. A patient may have multiple finished consultant episodes during a finished admission episode, a continuous period of care within a particular hospital.

AHES-APC provides a breakdown of finished consultant episodes at procedure-level (OPCS classification). In contrast, MHES-APC only provides a breakdown for finished consultant episodes at specialty-level and whilst a subtotal is given for finished consultant episodes with procedures performed, this includes a wider range of treatments than our definition of elective procedures; for example, simple injections, minor procedures (e.g. paracentesis, lumbar puncture), drug infusions. Therefore, when possible, it is preferable to use AHES- APC data since this provides a more granular breakdown.

The totals for finished consultant episodes provided by both AHES-APC and MHES-APC are not broken down in to elective versus emergency episodes. Instead, both datasets provide subtotals for the number of finished admission episodes that are elective versus emergency episodes. Therefore, we used the proportion of finished admission episodes that are elective episodes as a surrogate for the proportion of finished consultant episodes that are elective.

We used the following equation to estimate the number of elective finished consultant episodes:

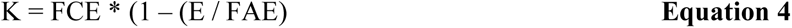

Where:

K = Number of elective finished consultant episodes FCE = Total number of finished consultant episodes

E = Number of finished admission episodes that were emergencies FAE = Total number of finished admission episodes

### Pandemic shortfall in elective procedures

The methodology for estimating the shortfall in elective procedures expanded on methodology previously published in The Lancet^6^.

For the purpose of this study, we considered the COVID-19 pandemic to have started on 1 January 2020; the first COVID-19 case was reported in Wuhan on 31 December 2019. The most recent available MHES-APC data was for March 2022. Therefore, it was possible to estimate the shortfall in elective procedures over the period January 2020 to March 2022 (27 months).

We calculated the shortfall in elective procedures in the following way for the pandemic period (January 2020 to March 2022):

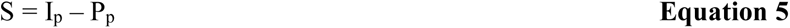

Where:

S = Pandemic shortfall in January 2020 to March 2022

Ip = Incident need for elective procedures during the pandemic period

Pp = Number of procedures completed during the pandemic period

#### Calculation of incident need

Elective procedure activity in 2018-19 (last pre-pandemic year for which AHES-APC data is available) was used as the baseline to estimate incident need for elective procedures.

However, as England has a growing and aging population, incident need for elective procedures in absolute terms can be expected to increase over time. To take in to account projected changes in population structure, we estimated age-sex incident need rates and applied these to ONS population structure projections for 2020-29.

Age and sex specific incident need rates were calculated based on the AHES-APC data for 2018-19. The following equation was used to calculate age and sex specific incident need rates:

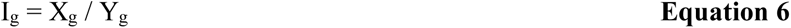

Where:

Ig = Incident need rate for elective procedures for a specific age-sex group

Xg = Number of elective procedures performed for a specific age-sex group in 2018-19

Yg = Total population for a specific age-sex group in England in mid-2018

We calculated total incident need for each year by summing incident need for each age-sex group:

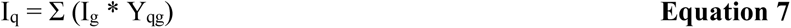

Where:

Iq = Total incident need for elective procedures in year q

Ig = Incident need rate for elective procedures for age-sex group

Yqg = Population for age-sex group in England in year q

To estimate incident need for elective procedures during the pandemic period (January 2020 to March 2022) age and sex specific incident need rates were calculated for the following groups (age-sex categorisation 1): female <18 years, female 18-39 years, female 40-64 years, female 65-79 years, female ≥80 years, male <18 years, male 18-39 years, male 40-64 years, male 65-79 years, male ≥80 years. Calculations were performed at OPCS code level to enable estimation of procedure-level pandemic shortfalls and therefore procedure-level total need in March 2022.

The same methodology was used to estimate incident need for elective procedures in April 2022 to December 2029, with the exception that we calculated overall total need for each year, rather than procedure-level need. This allowed us to use more granular age-sex groups for the calculation of age-sex specific incident need rates (age-sex categorisation 2); we calculated age-sex specific incident need rates separately for females and for males, with the following age breakdown: 0-4, 5-9, 10-14, 15-19, 20-24, 25-29, 30-34, 35-39, 40-44, 45-49, 50-54, 55-59, 60-64, 65-69, 70-74, 75-79, 80-84, 85-89, ≥90 years.

#### Calculation of pandemic shortfall using AHES-APC data

The AHES-APC dataset was used in preference to MHES-APC when possible, as it enables data to be extracted for elective procedures meeting the study inclusion criteria. Therefore, the following data sources were used:

- Pre-pandemic baseline number and breakdown of elective procedures was based on AHES-APC data for 2018-19 (the last full NHS year before the pandemic).
- For the pandemic period:
- `o` Data for April 2020 to March 2021 was based on AHES-APC data for 2020- 21 (the only HES-APC dataset where the full reporting period was during the pandemic).
- `o` Data for January to March 2020, and April 2021 to March 2022 were based on MHES-APC data. The most recent available MHES-APC data are for March 2022.

The AHES-APC dataset includes a breakdown of finished consultant episodes by age and separately by sex. This allowed us to calculate shortfall in 2020-21 for each OPCS code for ten age-sex groups (age-sex categorisation 1, see above) based on Equation 5.

#### Calculation of pandemic shortfall using MHES-APC data

We first summed monthly MHES-APC data for April 2018 to March 2019 (pre-pandemic baseline), and for January to March 2020 and April 2021 to March 2022 (15 month pandemic period for which AHES-APC data is not available) to calculate specialty-level totals.

As MHES-APC includes a wider range of treatments than the elective procedures included in this study, we had to estimate the number of elective procedures completed in January to March 2020 and April 2021 to March 2022 from the MHES-APC data. To do this, for each specialty, we calculated the number of elective procedures in 2018-19 in AHES-APC as a proportion of total patients recorded in MHES-APC in 2018-19. We applied this proportion to MHES-APC total for the 15 pandemic months to calculate a specialty-level breakdown of elective procedures performed during this period.

The specialty-level shortfall in procedures during January to March 2020 and April 2021 to March 2022 was calculated by subtracting specialty-level estimates of elective procedures performed from the calculated incident need for elective procedures during these months.

We estimated procedure-level breakdown for the shortfall during January to March 2020 and April 2021 to March 2022 based on the assumption that the pattern of this shortfall would be consistent with the shortfalls observed in 2020-21. To do this, we took the procedure-level shortfalls calculated for 2020-21 and summed them at specialty-level (for procedures where an increase rather than decrease (shortfall) in activity was observed in 2020-21, the shortfall was set as zero). We then calculated the proportion of the shortfall accounted for by each procedure within its specialty. These proportions were then applied to specialty-level shortfalls for January to March 2020 and April 2021 to March 2022 to achieve a procedure-level breakdown for that period.

### NHS waiting list

NHS waiting list data were accessed from NHS Digital to estimate the number of patients on the NHS waiting list who were waiting for elective procedures in December 2019 (last pre-pandemic month) and March 2022^7^.

Monthly NHS Waiting list data are reported by specialty for three categories:

- Incomplete pathways: patients who have been referred for treatment and are on the waiting list. This includes both patients waiting for clinic review, as well as those who are waiting for hospital admission for treatment (e.g. surgery, endoscopy).
- Admitted pathways: patients whose treatment was completed that month and whose treatment included an admitted care. This includes both day-case and inpatient admissions for a wider range of treatments than our definition of elective procedures (i.e. includes surgery, endoscopy, interventional radiology and cardiology, but also simple injections, lumbar puncture, drug infusions etc).
- Non-admitted pathways: patients whose treatment was completed that month and whose treatment did not involve admitted care (e.g. discharged from outpatient clinic).

For each specialty category in the NHS waiting list data, we estimated the number of incomplete pathways for that would translate to admitted pathways when completed using the following equation:

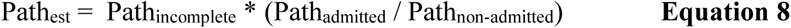

Where:

Pathest = Number of patients on NHS waiting list that month who will require admitted care

Pathincomplete = Number of patients on NHS waiting list that month whose pathways are incomplete

Pathadmitted = Number of patients who completed their pathway that month and whose treatment included admitted care

Pathnon-admitted = Number of patients who completed their pathway that month and whose treatment did not include admitted care

We made the following changes to the specialty categories to facilitate combination of NHS waiting list and pandemic shortfall estimates:

- We combined Gastroenterology with General Surgery.
- We combined Thoracic Medicine with Cardiothoracic Surgery.
- We excluded the following specialties from our calculations as they would not typically meet our definition for elective procedures: General Medicine, Dermatology, Neurology, Rheumatology, Geriatric Medicine, Other Medical Services, Other Mental Health Services, Other Paediatric Services, Other Services. Although some procedures that would be on the Oral Surgery waiting list are included in this study, the overwhelming majority of procedures are simple extractions that are excluded. Therefore we excluded the Oral Surgery figures from our calculations.
- Prior to 2022, the waiting list data included an ’Other’ category. In the March 2022 data this was disaggregated in to five subcategories: one surgical [Other Surgical Services] and four non-surgical. We calculated the proportion of the ’Other’ waiting list cases in March 2022 that were ’Other Surgical Services’ and applied this proportion to the totals for ’Other’ waiting list cases in 2019 and earlier, to estimate ’Other Surgical Services’ in those months. The estimates for pandemic shortfalls that we produced did not include an ’Other’ category, so we distributed the ’Other’ patients to the other (included) specialty categories in the waiting list data. Patients were distributed according to the proportion of the total waiting list (excluding ’Other’) accounted for by each specialty.

#### Composition of NHS waiting list

The NHS waiting list data are only publicly available at specialty-level. Therefore, in order to estimate total need in March 2022 at procedure-level we created estimates for the procedure-level composition of the NHS waiting list in December 2019 (Equation 3).

The AHES-APC dataset provides procedure-level data for:

- The number of elective procedures performed on patients admitted from waiting lists. Some elective procedures are planned rather than waiting list cases, meaning that the patient is given a procedure date at the time of booking (e.g. may happen for follow-up procedures).
- The mean number of days on the waiting list.

We used the 2018-19 AHES-APC data to multiply the number of waiting list admissions by their mean wait, to estimate at procedure-level an aggregated total for the number of days waited by patients for that procedure that year. Using the NHS waiting list data described above, we then calculated, at specialty-level, the proportion of the total waiting time within each specialty accounted for by each procedure. We then applied these proportions to the total number of patients on the NHS waiting list in December 2019 by specialty, to estimate the number of patients on the NHS waiting list in December 2019 at a procedure-level.

### Estimated cost to address need for elective procedures

We projected the cost to address the need for elective procedures based on the NHS reference costs for 2019-20^12^, the most recent available. NHS providers submit cost data to NHS England which calculates an average cost per care episode. Reference costs are provided by health resource group (HRG), a grouping similar treatment which require comparable healthcare resources. HRGs are determined by both the treatment a patient received and also their demographics (age) and comorbidities. There is no readily available cross-reference of HRGs against OPCS codes. Therefore, for each of the 130 procedures included in this study we identified the most appropriate HRG code. We aimed to produce a conservative estimate, therefore, when there was a choice of possible relevant HRG codes we selected the least expensive possibility. When applicable, we selected HRG codes for adult and those applying to patients with a comorbidity score of 1 (Supplementary Table 3). This process was completed by two investigators and any discrepancies were resolved through discussion.

We calculated the total cost for each procedure:

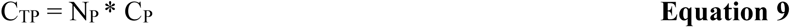

Where:

CTP = Total cost for procedure group

NP = Total need per procedure

CP = Procedure-specific cost per procedure group

We summed procedure-level costs to calculate total specialty-level costs.

### ICS-level breakdown

In order to estimate a regional breakdown for need for elective procedures, NHS waiting list data for March 2022 was accessed at Integrated Care System (ICS) level^7^. The ICS structure will be formally introduced in July 2022. Each ICS brings together hospital providers with other health and social care bodies within a geographic region. There are 42 ICSs in England with their population ranging from half a million to three million. In addition, there is a separate waiting list for nationally commissioned services.

The ICS-level NHS waiting list data was available broken down by specialty. Specialties were included in the analysis as described above. In order to estimate the total need for surgery at ICS-level, the hidden waiting list was estimated for each specialty for each ICS plus the nationally commissioned services. This was achieved by calculating for each ICS the proportion of all patients on the national NHS waiting list for a particular specialty in that ICS (e.g. Dorset has 11,674 patients on the General Surgery NHS waiting list, accounting for 1.4% of all patients on the national General Surgery NHS waiting list). This proportion was then multiplied by the national estimate for the hidden waiting list for that specialty and added to the NHS waiting list figure to calculate total need (e.g. since the national hidden waiting list for General Surgery is 1,793,946 for Dorset this was multiplied by 1.4% and the result added to 11,674, resulting in an estimate of 25,208 General Surgery procedures needed in Dorset). This produced specialty-level estimates of total need for each of the 42 ICSs and also for nationally commissioned services.

It was assumed that for each specialty the waiting list for nationally commissioned services in each ICS would be proportionate to the waiting list for locally commissioned services. We calculated for each ICS the proportion of all patients on the national NHS waiting list for that specialty (excluding nationally commissioned services from the denominator). This proportion was then multiplied by the total for the relevant specialty for nationally commissioned services, and this was added to the existing estimate for need based on locally commissioned services to calculate an overall total need at specialty-level for each ICS. These figures were rounded to the nearest integer to allow easier interpretation.

For each ICS, specialty-level estimates for total need were summed to calculate an overall total need for elective procedures. To facilitate comparison across ICSs, the need for elective procedures was calculated per 1,000 population:

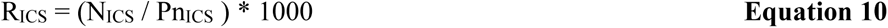

Where:

RICS = Elective procedures needed in ICS per 1,000 population

NICS = Total elective procedures needed in ICS

PnICS = ICS population in mid-2020

ICS-level projections for costs to address the need for elective procedures was calculated at specialty-level and summed to produce an overall figure. Using the national estimates, an average cost per procedure was calculated for each specialty. For each specialty, this figure was multiplied by the number of procedures needed in each ICS to estimate cost for that specialty in that ICS. Costs were summed across all specialties in each ICS to calculate a total cost per ICS. To aid interpretation a cost per capita was calculated:

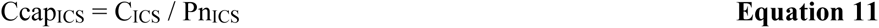

Where:

CcapICS = Cost per capita to address the need for elective procedures in ICS

CICS = Total cost to address the need for elective procedures in ICS

PnICS = ICS population in mid-2020

Data was also aggregated at a regional-level based on the seven regions defined by NHS England (East of England, London, Midlands, North East & Yorkshire, North West, South East, South West)

### Forecasting total need to 2030

We conducted an expert survey to ascertain expectations for the speed of recovery for elective services and the potential to expand beyond pre-pandemic volume. We used an anonymous online survey to collect responses from members of the CovidSurg research network. The survey instrument is included in Supplement 1. A total of 47 clinicians across 15 specialities submitted responses and the results are summarised in Supplementary Table 4.

Based on the results of the expert survey we have modelled four scenarios to estimate the number of people in England who will need elective procedures through to 2030.

These scenarios were

- Current capacity: surgical volume remains at the same level as in February-March 2022, from April 2022 up until January 2030.
- Pessimistic scenario: elective procedure volume gradually increases from current levels up to pre-pandemic levels by July 2023 followed by a plateauing of this rate through to January 2030.
- Central scenario: elective procedure volume returns to pre-pandemic levels by December 2022 followed by a 2% increase per year.
- Optimistic scenario: elective procedure volume returns to pre-pandemic levels by December 2022 followed by a 4% increase per year.

Scenarios were modelled on a monthly basis, with the total need for selective procedures calculated for each month between April 2022 and December 2029.

We also determined the monthly increase in surgical volume (assuming a constant rate of increase) required to fulfil the need for surgery and invasive diagnostics in full by January 2030.

## Results

We estimated the total need for elective procedures in England in March 2022 was 4,347,469. This is the highest need since at least 2007 (Figure 1). The greatest need was for General Surgery (35.0%, 1,522,366 of 4,347,469), Orthopaedics (22.5%, 976,875 of 4,347,469), and Ophthalmology (9.0%, 391,683 of 4,347,469, Table 1). There was substantial geographic variation (Figure 2), with the highest need in the South West (93.2 elective procedures needed per 100,000) followed by the North West (89.7 per 100,000), Midlands (83.9 per 100,000), South East (77.6 per 100,000), East of England (73.8 per 100,000), North East and Yorkshire (69.6 per 100,000), and London (56.7 per 100,000).

**Figure 1:**
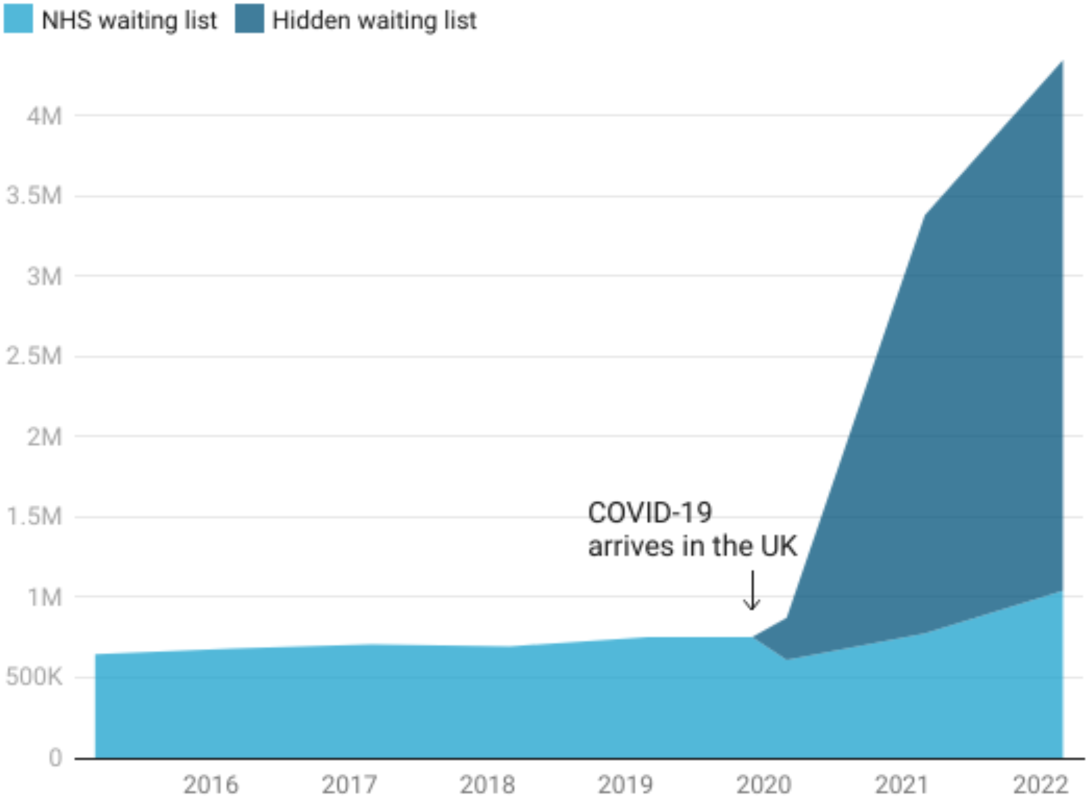
Number of elective procedures needed in England (2015-2022) Estimates for number of elective procedures needed were based on analysis of Consultant-led Referral to Treatment Waiting Times as detailed in main text. Figure created in Datawrapper.

**Figure 2:**
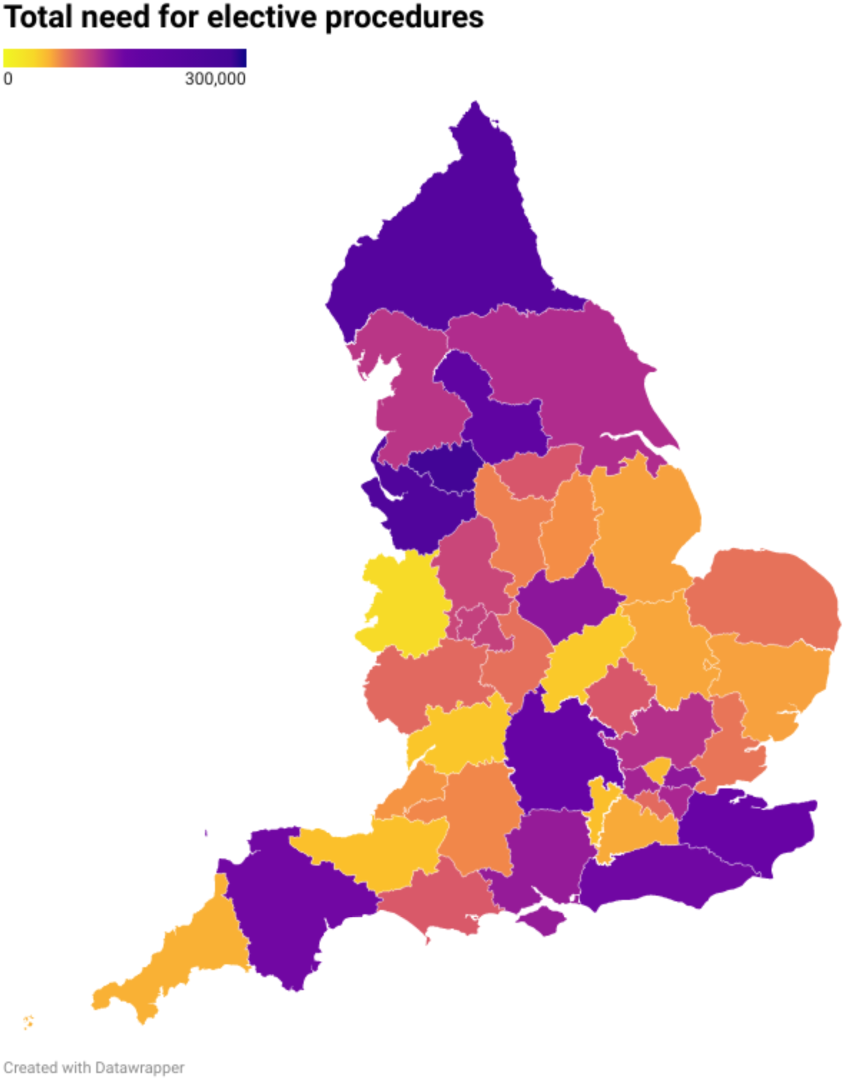
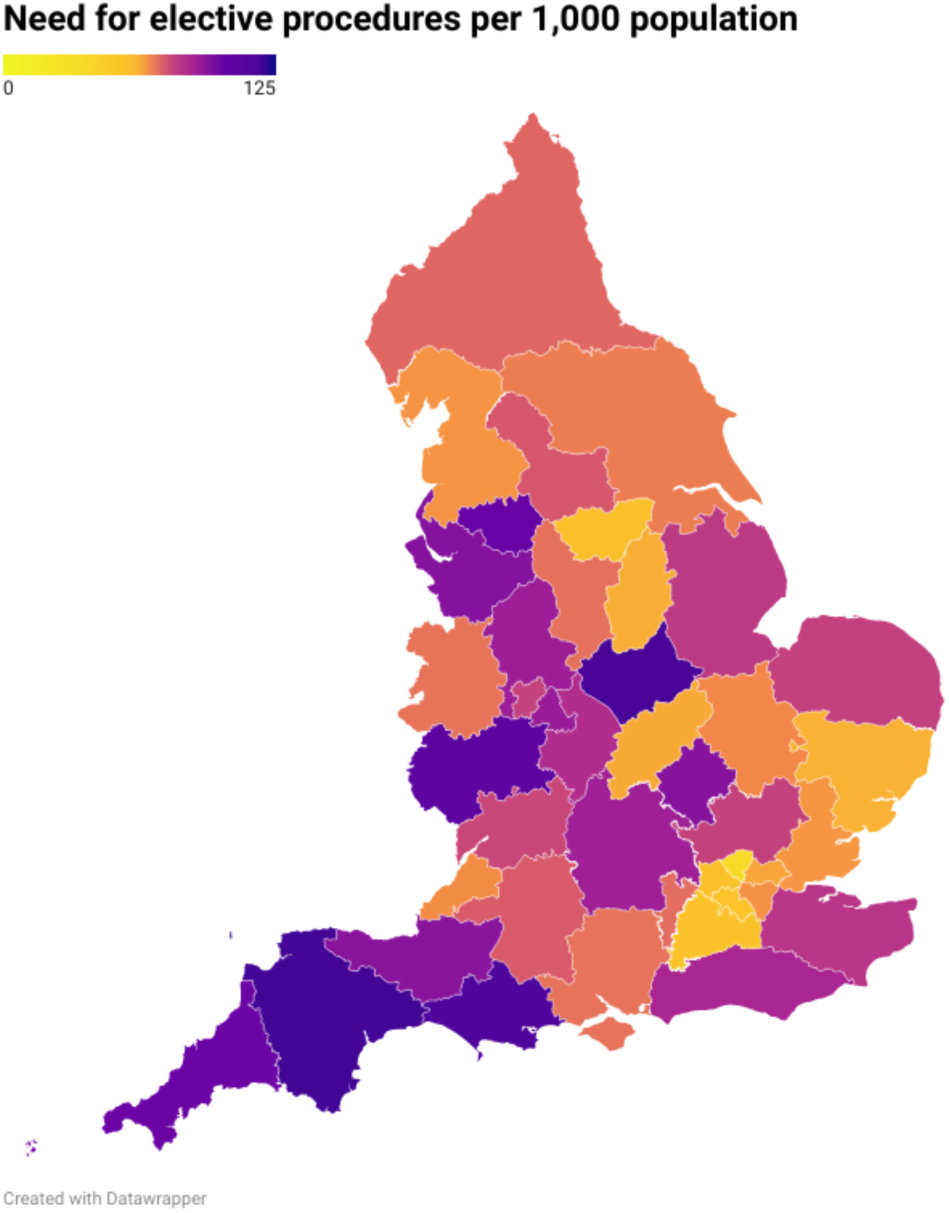
Total need for elective procedures in England per 1,000 population

**Table 1:** Specialty-level breakdown of estimated total need for elective procedures in England and estimated hidden waiting list in March 2022

| **Specialty** | **Total need** | **Hidden waiting list** |
| --- | --- | --- |
| General Surgery | 1,522,366 | 1,289,440 |
| Orthopaedics | 976,875 | 747,749 |
| Ophthalmology | 391,683 | 168,361 |
| Urology | 385,971 | 284,279 |
| Head & Neck surgery | 332,281 | 263,447 |
| Gynaecology | 246,973 | 175,394 |
| Plastic Surgery | 231,333 | 182,570 |
| Cardiology | 154,453 | 109,479 |
| Cardiothoracic Surgery | 88,865 | 76,286 |
| Neurosurgery | 16,669 | 7,508 |
| **Total** | **4,347,469** | **3,304,513** |

### Hidden waiting list

Of the 4,347,469 required elective procedures, 1,042,956 (24.0%) of patients were on the NHS waiting list and 3,304,513 (76.0%) were on a hidden waiting list. The largest hidden waiting lists were for General Surgery (1,289,440) and Orthopaedics (747,749).

### Day-case procedures

Overall, 84.9% (3,692,377 of 4,347,469) need was for day-case procedures. For six sub- specialties (Cardiology, Colorectal Surgery, Head & Neck Surgery, Oesophagogastric Surgery, Ophthalmology, Plastic Surgery) >95% of total need was for day-cases (Table 2).

**Table 2:**
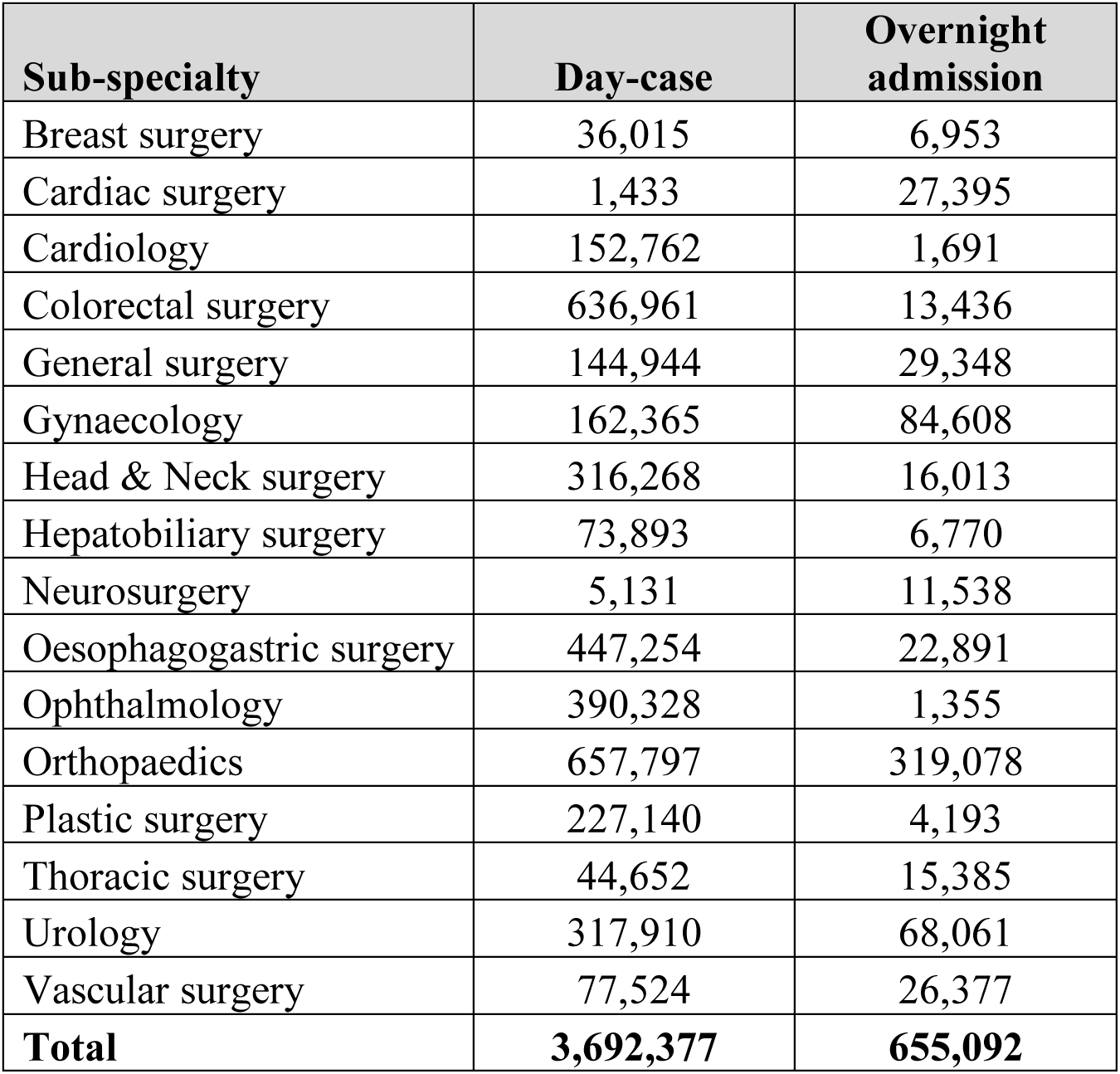
Sub-specialty-level breakdown of estimated total need for elective procedures in England in March 2022

### Key procedures

Overall, the greatest need was for sigmoidoscopy/ colonoscopy (568,838), gastroscopy (447,830), cataract surgery (314,790), lower limb joint replacement (224,363), and interventional cardiology (349,300). These top five procedures accounted for 39.3% of total need (1,710,274 of 4,347,469). The top 20 procedures (Table 3) accounted for 68.8% (2,992,395 of 4,347,469) of total need.

**Table 3:** Top 20 elective procedures by need in England in March 2022

| Procedure name | Day-case | Overnight admission | Total | Rank |
| --- | --- | --- | --- | --- |
| Sigmoidoscopy, colonoscopy | 568,886 | - | 568,838 | 1 |
| Gastroscopy | 446,125 | 1,705 | 447,830 | 2 |
| Cataract surgery | 314,790 | - | 314,790 | 3 |
| Lower limb joint replacement | - | 224,363 | 224,363 | 4 |
| Interventional cardiology | 152,762 | 1,691 | 154,453 | 5 |
| Ureteroscopy, cystoscopy & related procedures | 117,817 | 18,444 | 136,261 | 6 |
| Excision of lesion of skin | 128,131 | - | 128,131 | 7 |
| Operations on bladder | 90,486 | 23,263 | 113,749 | 8 |
| Procedures on peripheral nerves | 105,915 | - | 105,915 | 9 |
| Operations on ear | 86,216 | 1,347 | 87,563 | 10 |
| Spinal nerve root procedure | 78,130 | - | 78,130 | 11 |
| Operations on nose | 77,855 | 4 | 77,859 | 12 |
| Groin hernia repair | 76,792 | - | 76,792 | 13 |
| Arthroscopic procedures | 75,431 | - | 75,431 | 14 |
| Surgery on ligament, tendon, muscle | 72,068 | 1,658 | 73,726 | 15 |
| Hysteroscopy & related procedures | 68,744 | - | 68,744 | 16 |
| Other day-case spinal surgery | 68,697 | - | 68,697 | 17 |
| Division or excision of bone | 63,069 | 5,322 | 68,391 | 18 |
| Tonsillectomy | 61,993 | - | 61,993 | 19 |
| Proctology | 60,739 | - | 60,739 | 20 |

Total need for Sigmoidoscopy, colonoscopy is lower than day-case need as the need for overnight admissions for this procedure was calculated as -48

### Projected cost to address need for elective procedures

The projected cost to fully address the need for elective procedures was £9,207,798,071. The greatest costs were for Orthopaedics (£3,751,715,700), Urology (£722,558,421), and Head & Neck Surgery (£585,055,737, Table 4).

**Table 4:** Projected cost to address need for elective procedures

| **Sub-specialty** | **Total cost** |
| --- | --- |
| Breast surgery | £221,703,319 |
| Cardiac surgery | £318,800,210 |
| Cardiology | £372,181,343 |
| Colorectal surgery | £540,488,976 |
| General surgery | £465,424,135 |
| Gynaecology | £555,252,435 |
| Head & Neck surgery | £585,055,737 |
| Hepatobiliary surgery | £227,444,145 |
| Neurosurgery | £130,995,974 |
| Oesophagogastric surgery | £335,191,264 |
| Ophthalmology | £394,740,194 |
| Orthopaedics | £3,751,715,700 |
| Plastic surgery | £210,128,413 |
| Thoracic surgery | £106,740,125 |
| Urology | £722,558,422 |
| Vascular surgery | £269,377,678 |
| **Total** | **£9,207,798,071** |

### Surgical activity in February to March 2022

The pre-pandemic baseline for the number of procedures expected in a two-month period was 953,390. We estimated that 810,674 elective procedures were performed in February to March 2022. This indicates a shortfall of 142,717 procedures, equivalent to 15.0% of pre-pandemic elective volume.

### Forecasting total need for elective procedures to 2030

We modelled that if age-sex specific incident need rates for elective procedures remain constant, based on ONS population projections in the seven years and nine months from April 2022 to December 2029, 47,957,079 elective procedures will be needed (Supplemental Table 5), in addition to the existing pandemic shortfall of 4,347,469. Therefore, the total number of elective procedures that will be needed through to 2030 is 52,304,550.

We modelled that if surgical volume remains at current levels, the total number of elective procedures needed would increase to 14,608,195 by January 2030 (Figure 3). In the pessimistic scenario, if elective procedure volume gradually increases from current levels up to pre-pandemic levels by July 2023 followed by a plateauing of this rate, the total number of elective procedures needed would increase to 8,507,087 by January 2030. In the central scenario, if elective procedure volume returns to pre-pandemic levels by December 2022 followed by a 2% increase per year, the total number of elective procedures needed in January 2030 would be 5,420,999. Finally, in the optimistic scenario, if elective procedure volume returns to pre-pandemic levels by December 2022 followed by a 4% increase per year, 2,584,664 procedures would be needed in January 2030.

**Figure 3:**
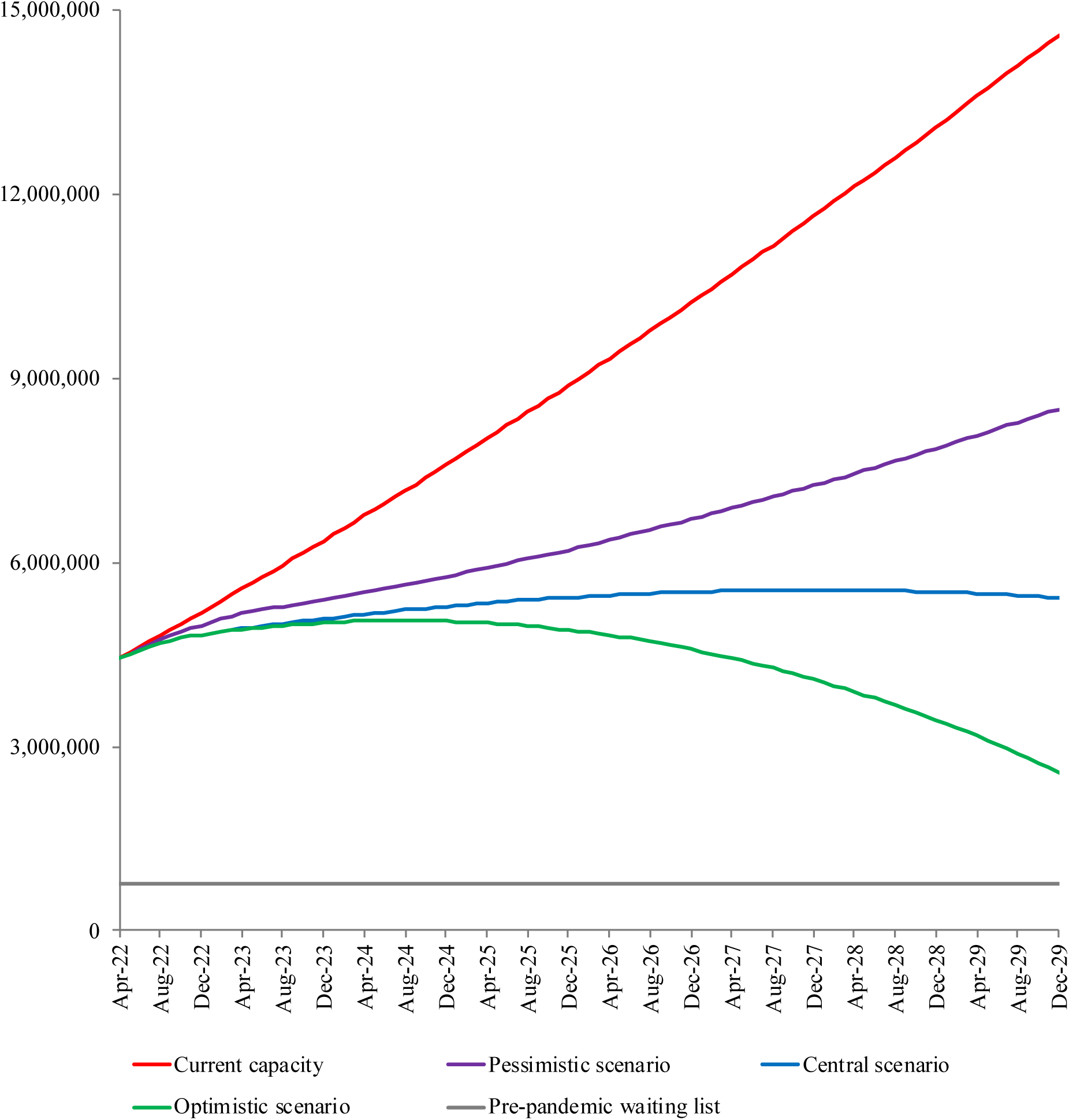
Forecasts for total need for elective procedures in England in different scenarios. The pre-pandemic waiting list in December 2019 was 753,116. Data is tabulated in Supplemental Table 6. Figure created in Datawrapper.

In order to fulfil the need for elective procedures in full by January 2030, elective procedure volume would need to increase by 8.4% (as a proportion of pre-pandemic volume) each year, reaching a maximum of 50% above the pre-pandemic baseline in January 2030.

## Discussion

We estimated that 4.3 million procedures were needed in England in March 2022. This is very considerably higher than the NHS waiting list; an estimated 3.3 million patients were on a hidden waiting list. Our projects suggest that the number of people needing elective procedures is likely to remain very considerably higher than the pre-pandemic waiting list peak until at least 2030. Unless elective procedure volume increases at least 2% above pre-pandemic levels each year, the number of people needing procedures will actually increase over time due to population growth and ageing. Based on our expert survey, the 8% annual increase in activity required to eliminate waiting lists even by 2030 is unlikely to be achievable.

### Impact of waiting lists on population health

The hidden waiting list includes people who need elective procedures and would have been referred and treated pre-pandemic. However, as a result of the pandemic these patients may have not sought medical attention. These patients will be delayed in receiving treatment and some will suffer worse health as a result. For example, patients with hip osteoarthritis needing a joint replacement are likely to experience increasing symptoms resulting in deterioration in their quality of life and their ability to complete their work and social activities. This in turn may in turn result in lifestyle changes. For patients with life threatening diseases such as cancer, delayed diagnosis and treatment could reduce chances of being successfully cured.

### Inequalities in the need for elective procedures

There is sharp variation in the overall need for elective procedures between England’s regions, with the need in the South West being almost double that in London. At a sub- regional level there is variation between different integrated care systems, but we were not able to explore variation at a more granular postcode level. It is possible that some patients living in more deprived neighbourhoods and particularly vulnerable patients such as those with mental health conditions, refugees, and intravenous drug users may find it more difficult to navigate the process to get elective treatment at a time when health services are under particularly high pressure. Moreover, a shift from NHS to private provision in more affluent neighbourhoods could relieve some pressure on NHS services in those areas, whilst services in more deprived areas remain under heavy pressure. It is important that funding is awarded to integrated care systems according to their actual needs; this analysis suggests where is wide variation in per capita funding needs. Health equity impact assessments^16^ should be conducted to ensure that planned initiatives to address the need for elective procedures are equitable both at national and local levels.

### Strategies to address need for elective procedures

As long waiting lists are likely to continue for the foreseeable future, difficult policy decisions are needed around the prioritisation and models of delivery for elective procedures. Consideration should be given to deprioritising procedures of lower clinical value, in order to release resources to focus on finding patients on the hidden waiting list who have serious disease requiring rapid treatment.

A key insight from this study is that most of the elective procedures needed are day-case procedures. This is likely to reflect the prioritisation of life saving surgery, such as cancer surgery, during the pandemic resulting in only small backlogs of major surgery. An important factor that could limit the ability to ramp up inpatient surgery is an ongoing pressure on general hospital beds in the NHS, particularly by high levels of emergency admissions.

However, day-case procedures present greater flexibility in ramping up activity, since they are not dependent on availability of either general or intensive care beds. Initiatives such as high intensity theatre lists have been piloted to focus on high volume low complexity day-case activity^17^, though further research is needed to determine their optimum format. However, an important caveat to any focus on day-case activity is the need to tackle the orthopaedic backlog.

### Strengths and limitations

This is the first study to focus on the need for elective procedures and to provide granular breakdowns at specialty-level and procedure-level. Previous studies have focussed on the overall NHS England waiting list, but this overlooks the large heterogeneity of treatments that patients are waiting for^9, 18–20^. The complexity of patient pathways for surgery, endoscopy, interventional cardiology and interventional radiology is far greater than for simpler treatments like drug infusions. Surgery, endoscopy, and interventional radiology all require specific suites equipped with expensive specialist machines, and staffed by highly trained multidisciplinary teams. Any initiative to increase elective procedure volume must therefore be carefully planned.

There are several limitations to this study. An assumption was made that the pre-pandemic NHS waiting list captured all patients who needed elective procedures, but it is likely that even pre-pandemic there were some patients who needed elective procedures who had not sought medical attention or had not been appropriately referred or listed. If this is the case, this would lead to us underestimating the current hidden waiting list and total need. NHS waiting list data is not fully cleaned. There may be some double counting of the same patient on different waiting lists, or the counting of patients who no longer require elective treatment, for example, because they have been admitted and treated as an emergency or because they have died. This would potentially result in a small over-estimation of the total need for elective procedures. However, because most of the total need is as a result of the pandemic shortfall, rather than the pre-pandemic NHS waiting list, the impact of inaccuracies in the NHS waiting list would be small overall.

We assumed that as waiting times for surgery increase there is no attrition to the need for surgery, but patients who would usually have been treated during the pandemic may no longer still need treatment; for example, if they have had successful non-operative treatment, have had spontaneous resolution of their underlying condition, or died. This would lead to an overestimation of the current need for surgery.

We assumed that age-sex specific incidence of symptoms of conditions requiring elective procedures would remain stable over time, but this may change as a result of wider social and economic circumstances, mediated through the wider determinants of health; for example, the need for cancer surgery is likely to grow if obesity rates continue to increase.

For our geographic analyses, we have assumed that the size of the hidden waiting list relative to the NHS waiting list is consistent across all NHS systems. If some systems have performed better at capturing patients in need of procedures on their NHS waiting lists, we may be over- estimating their total need, whilst under-estimating need in other systems that have unexpectedly large hidden waiting lists. However, this would not impact the accuracy of national-level estimates.

## Supporting information

Supplement 1 (Expert survey)

## Data Availability

All data produced in the present study are available upon reasonable request to the authors

## Research ethics

This study used publicly available, anonymised data only and is therefore exempt from the requirement for ethical review.

## Research reporting checklist

There is no appropriate checklist available for this modelling study.

## Data Availability Statement

All data produced in the present study are available upon reasonable request to the authors. Data requests can be emailed to.

---

**Supplemental Table 1:**
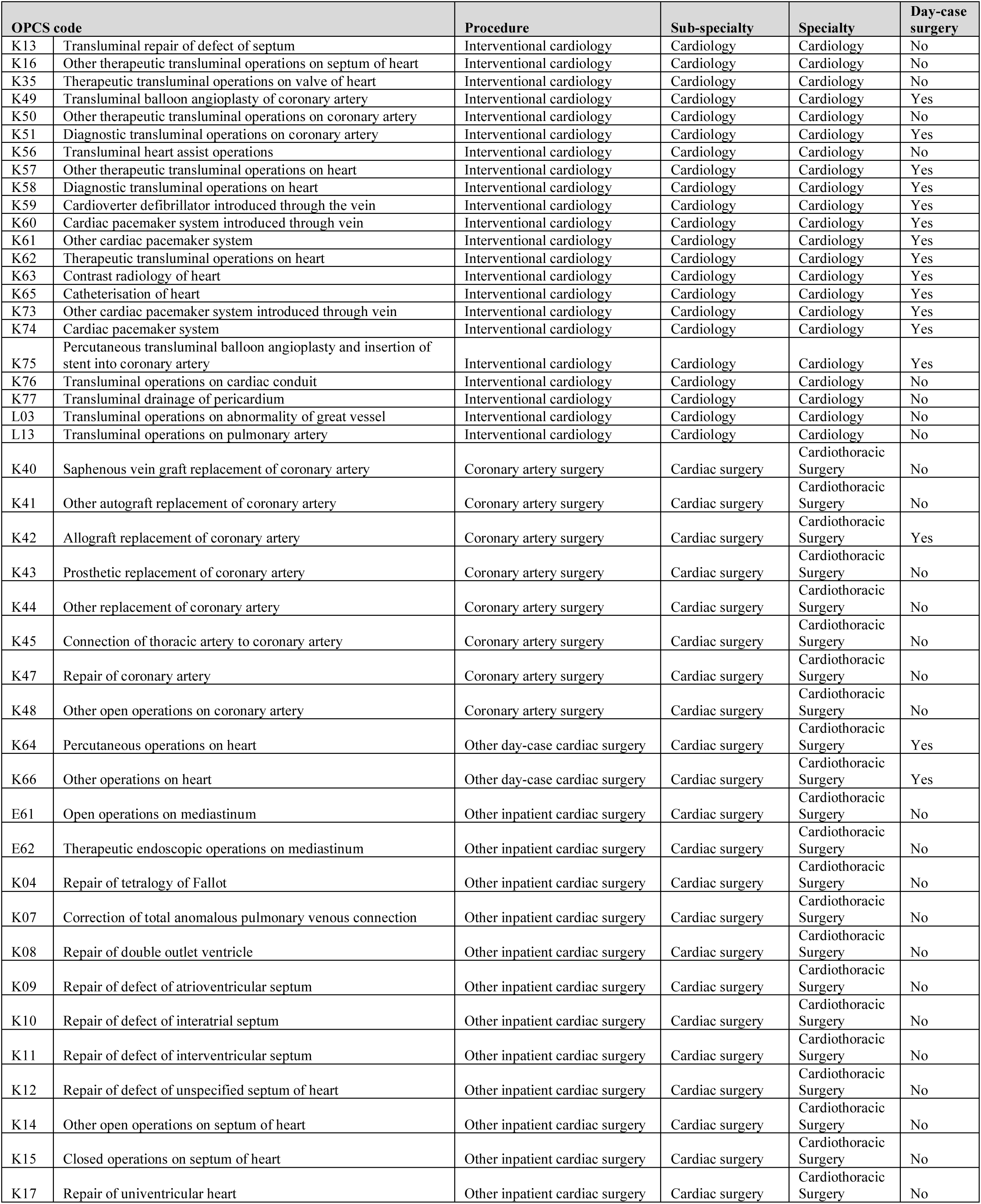
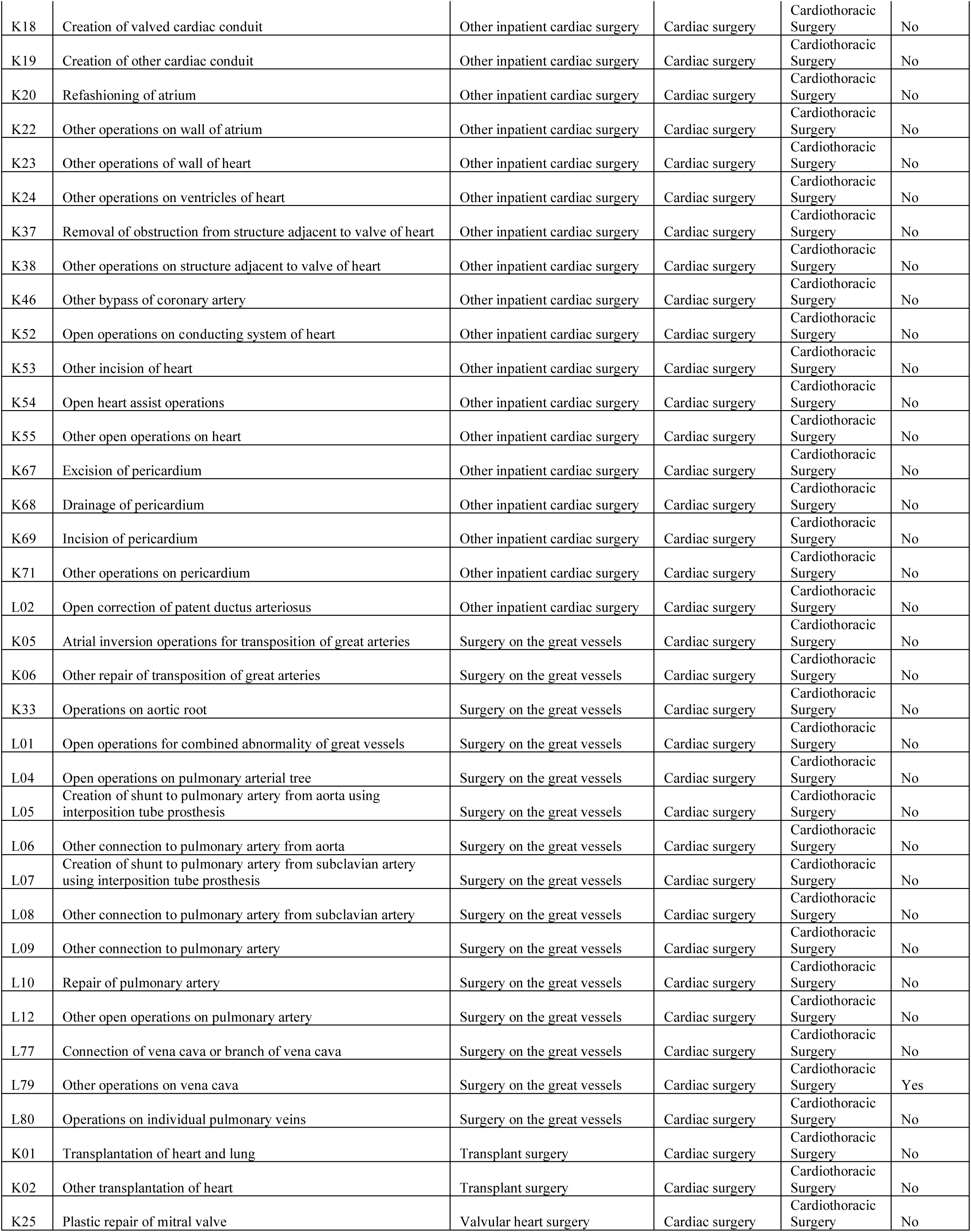
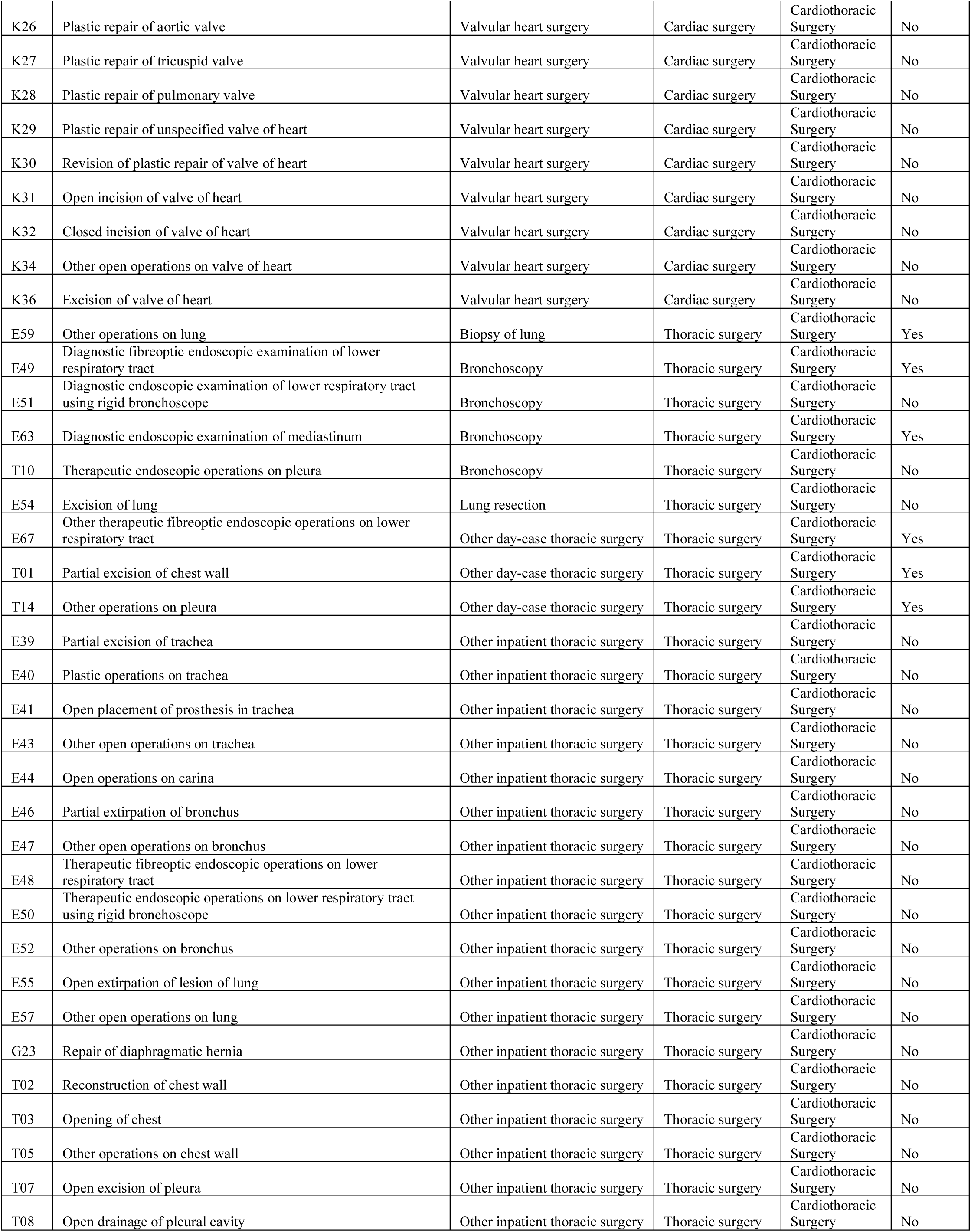
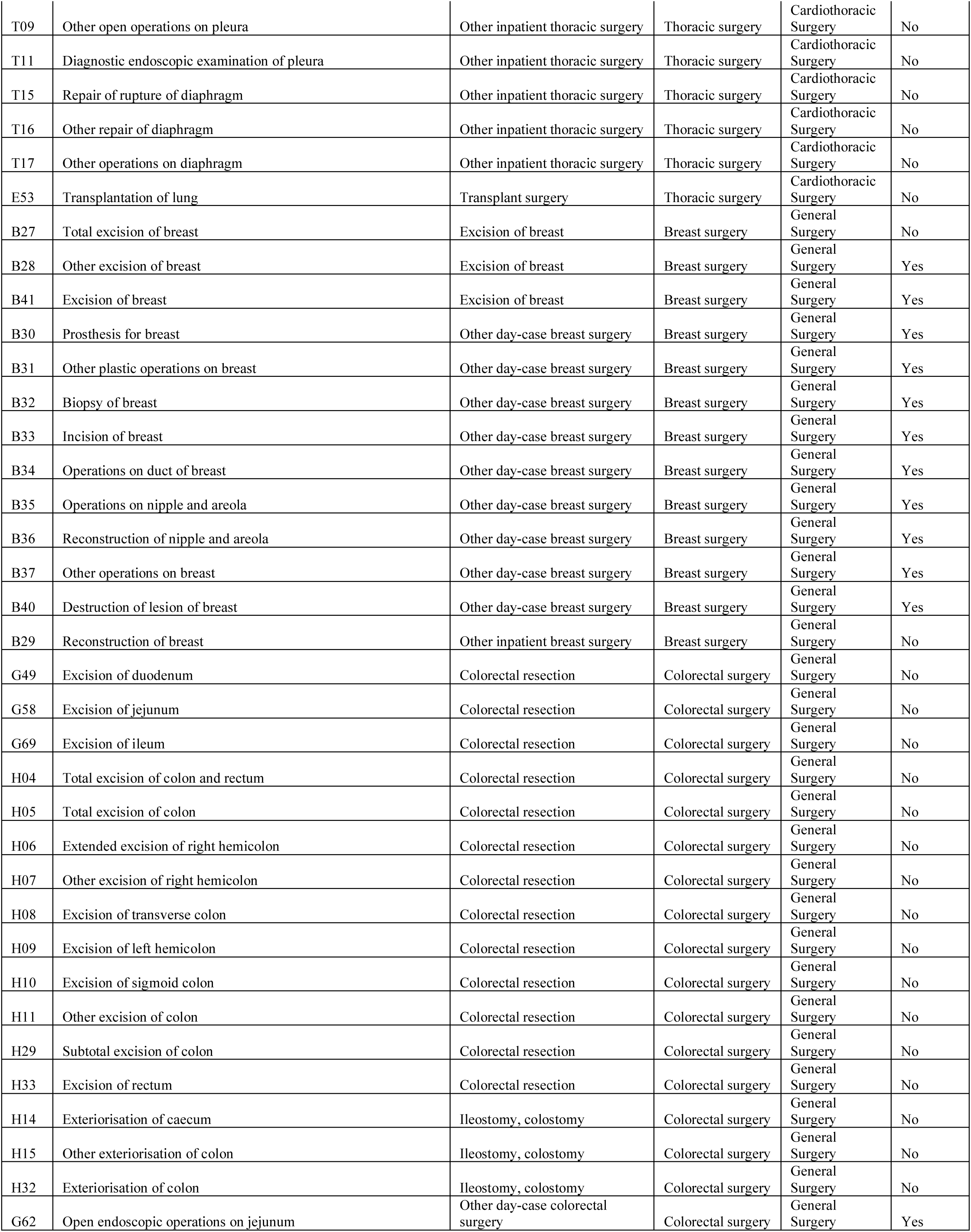
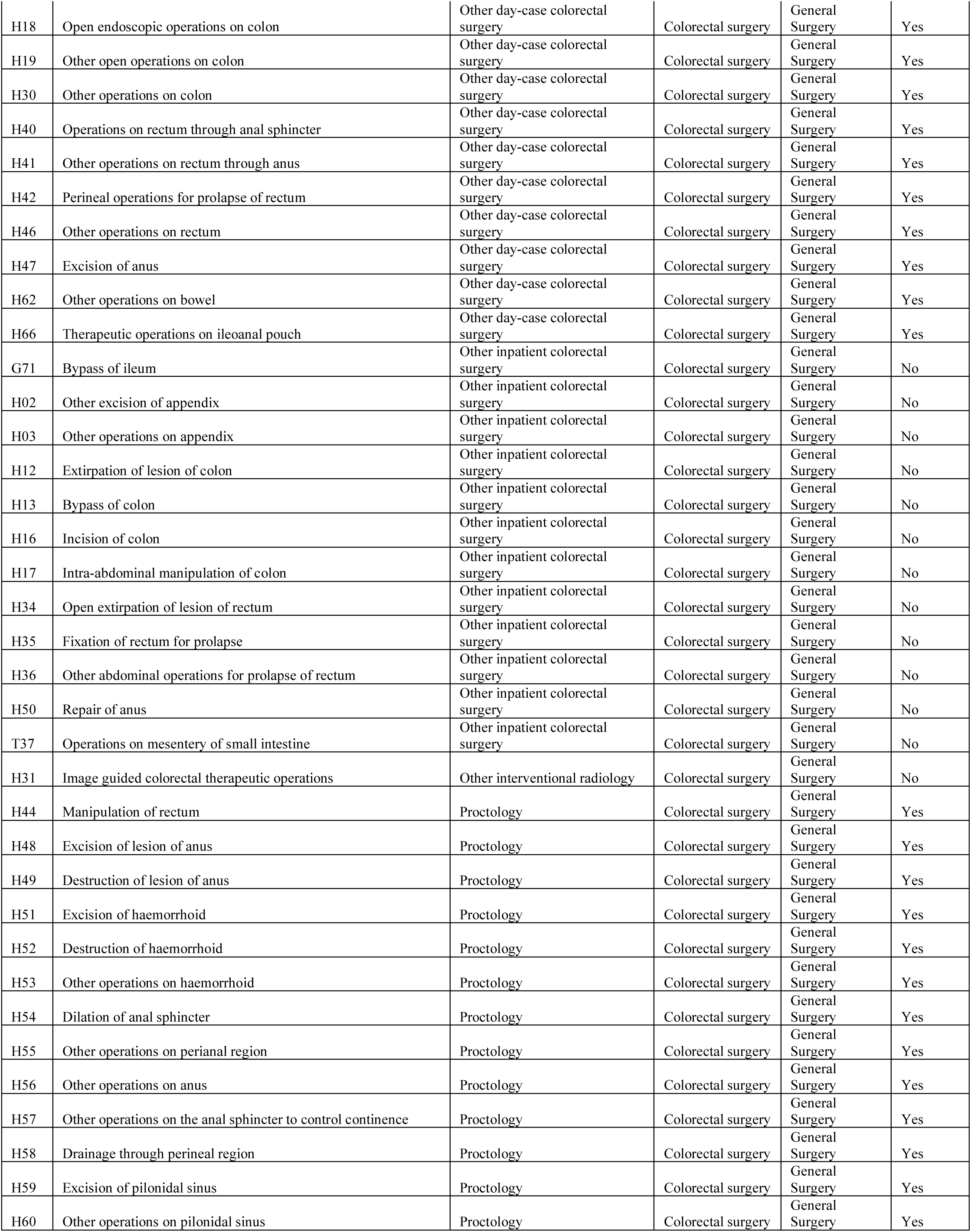
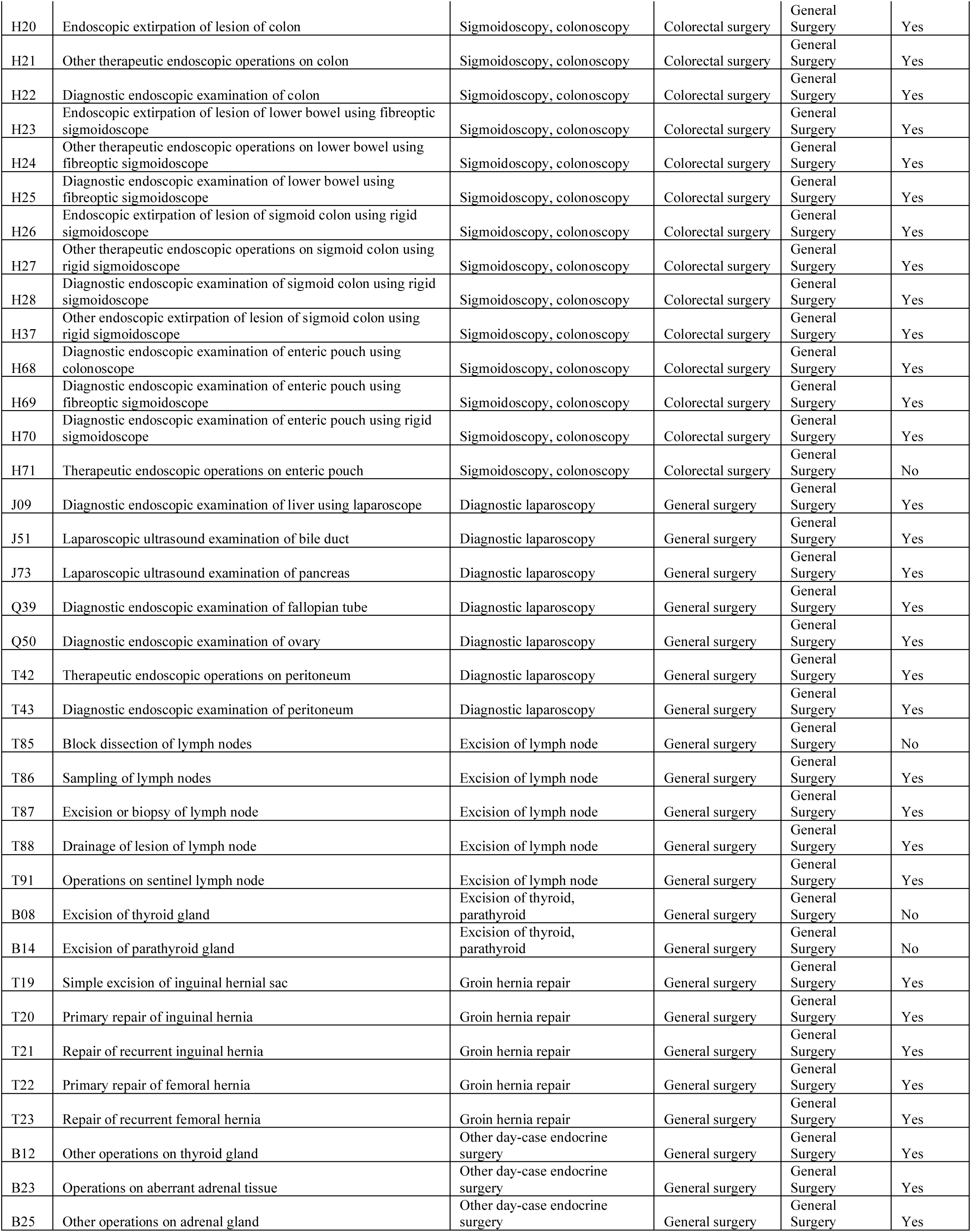
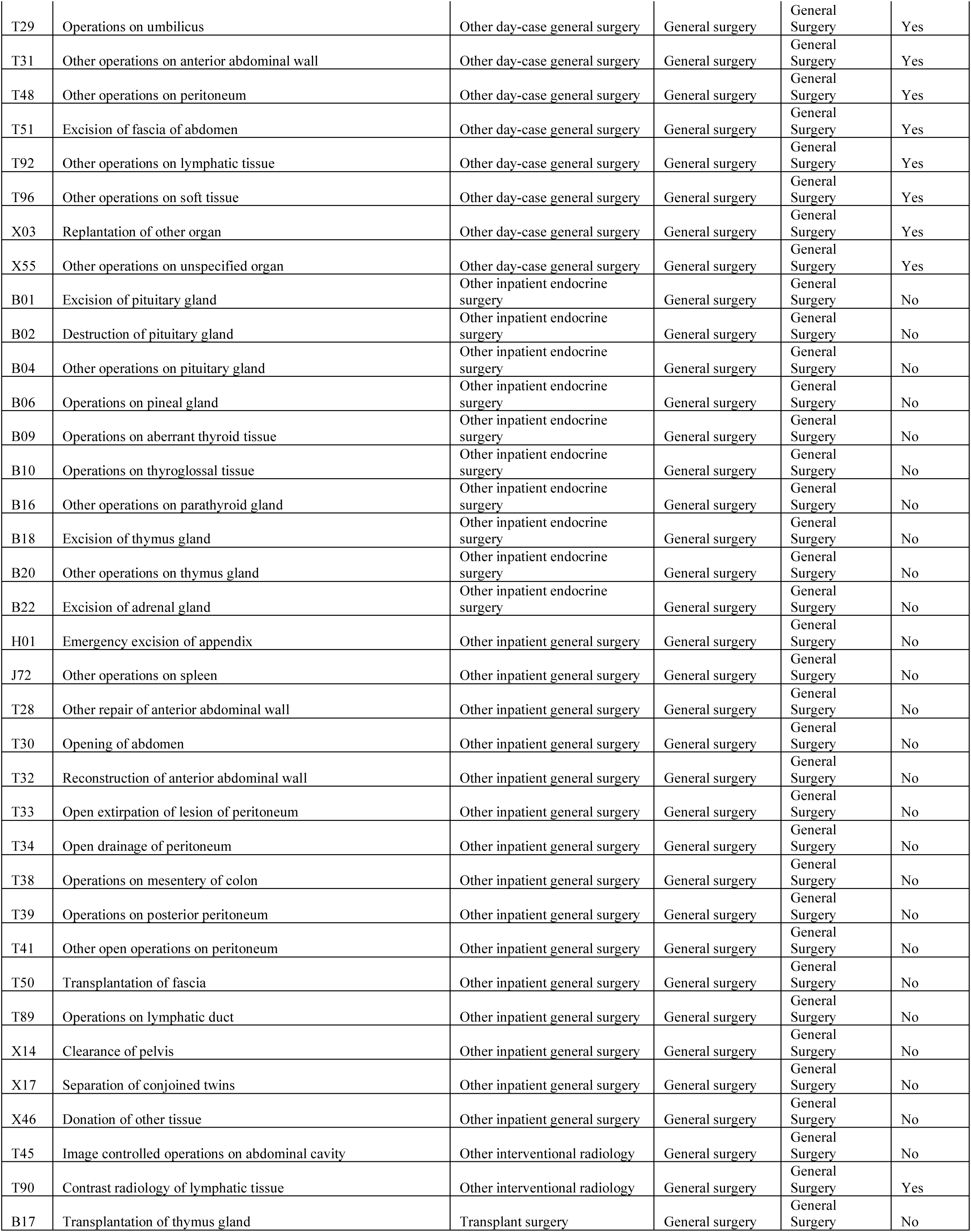
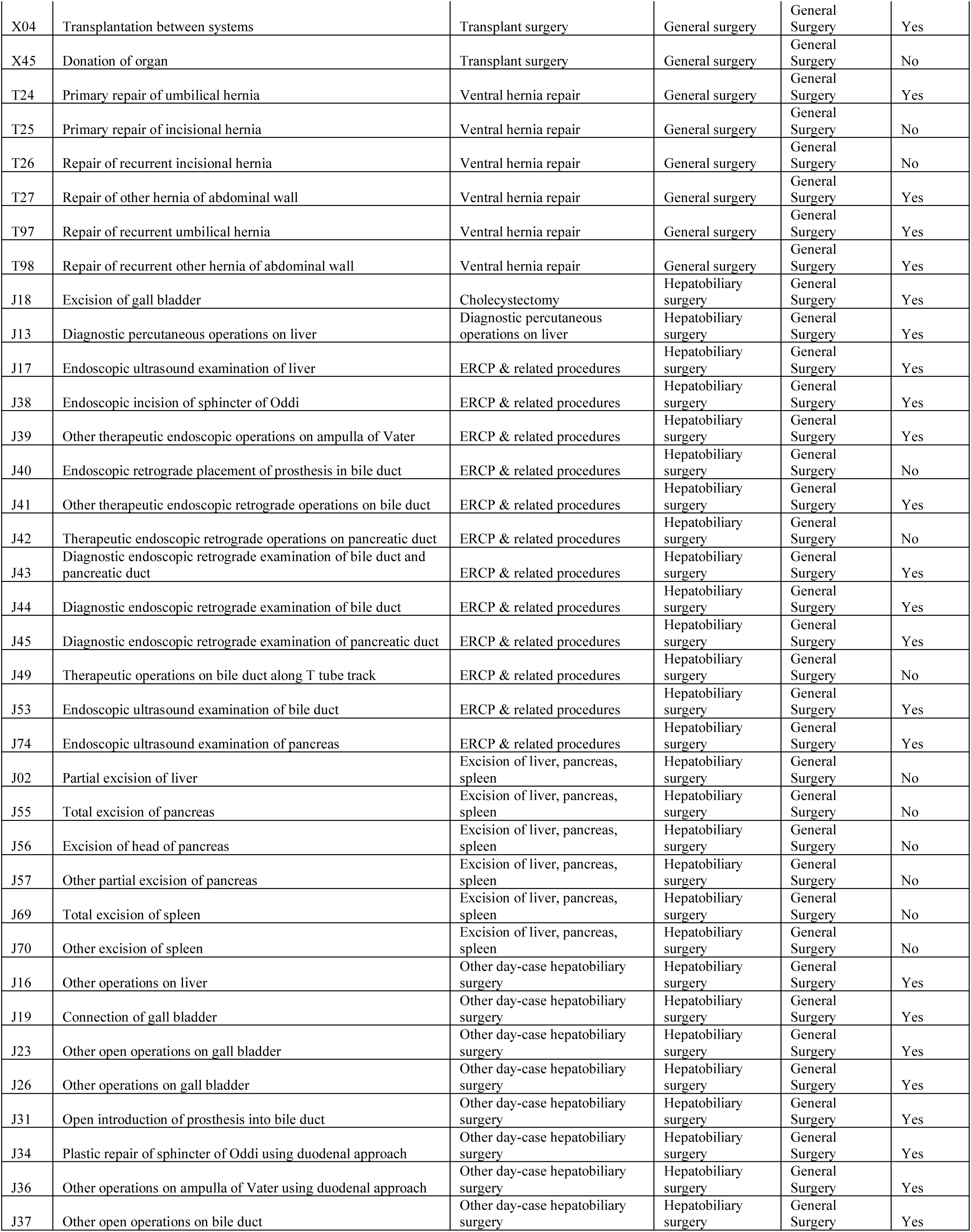
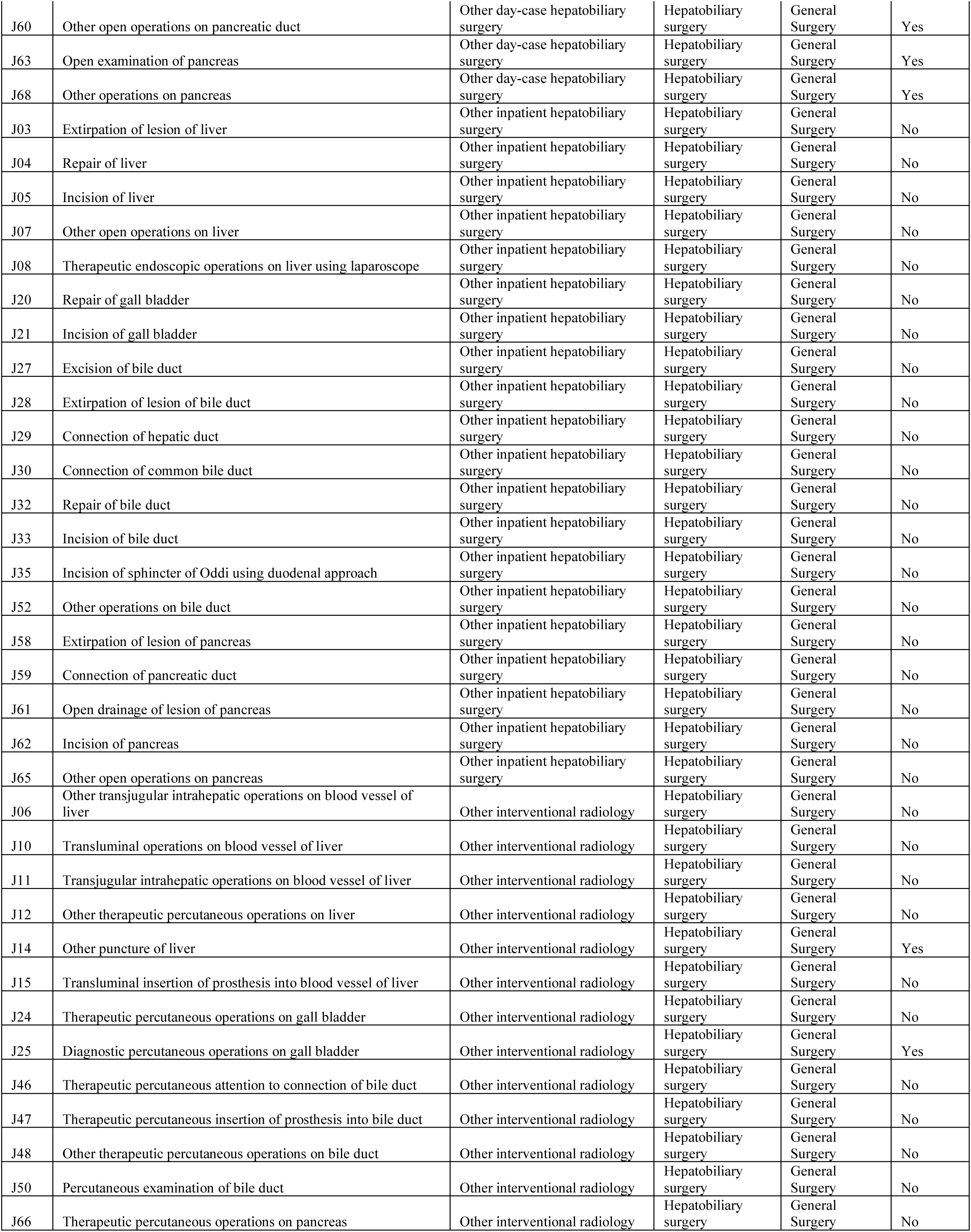
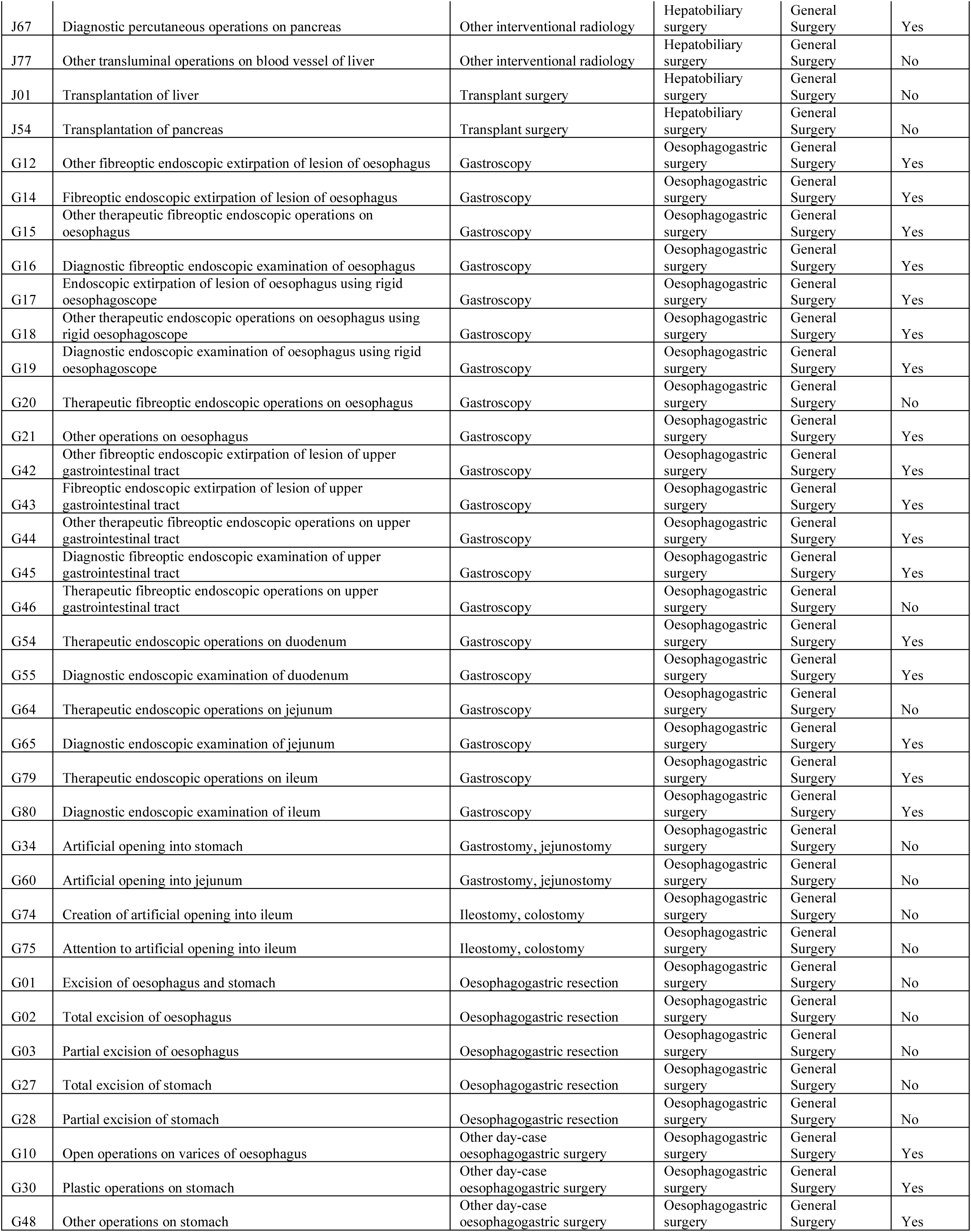
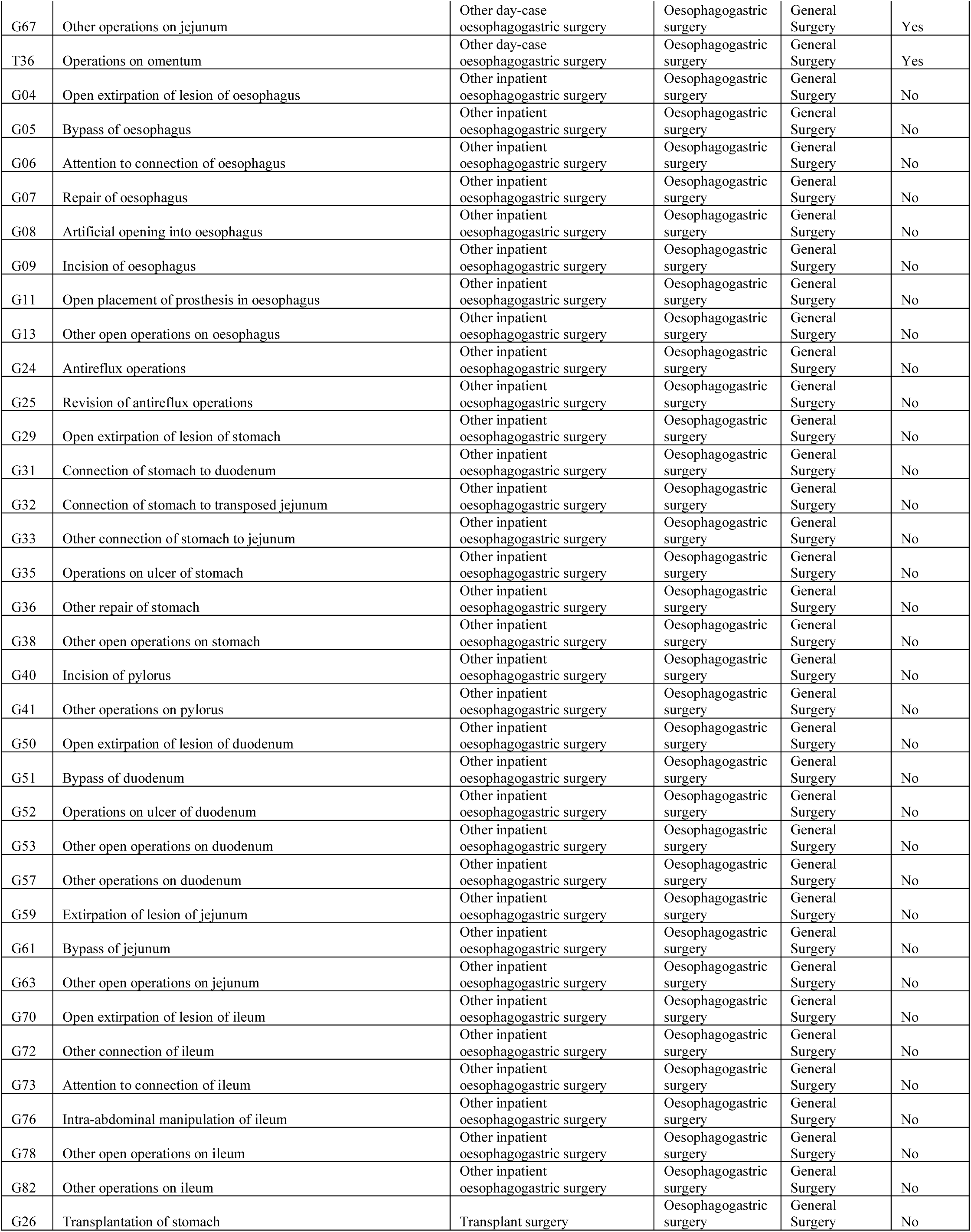
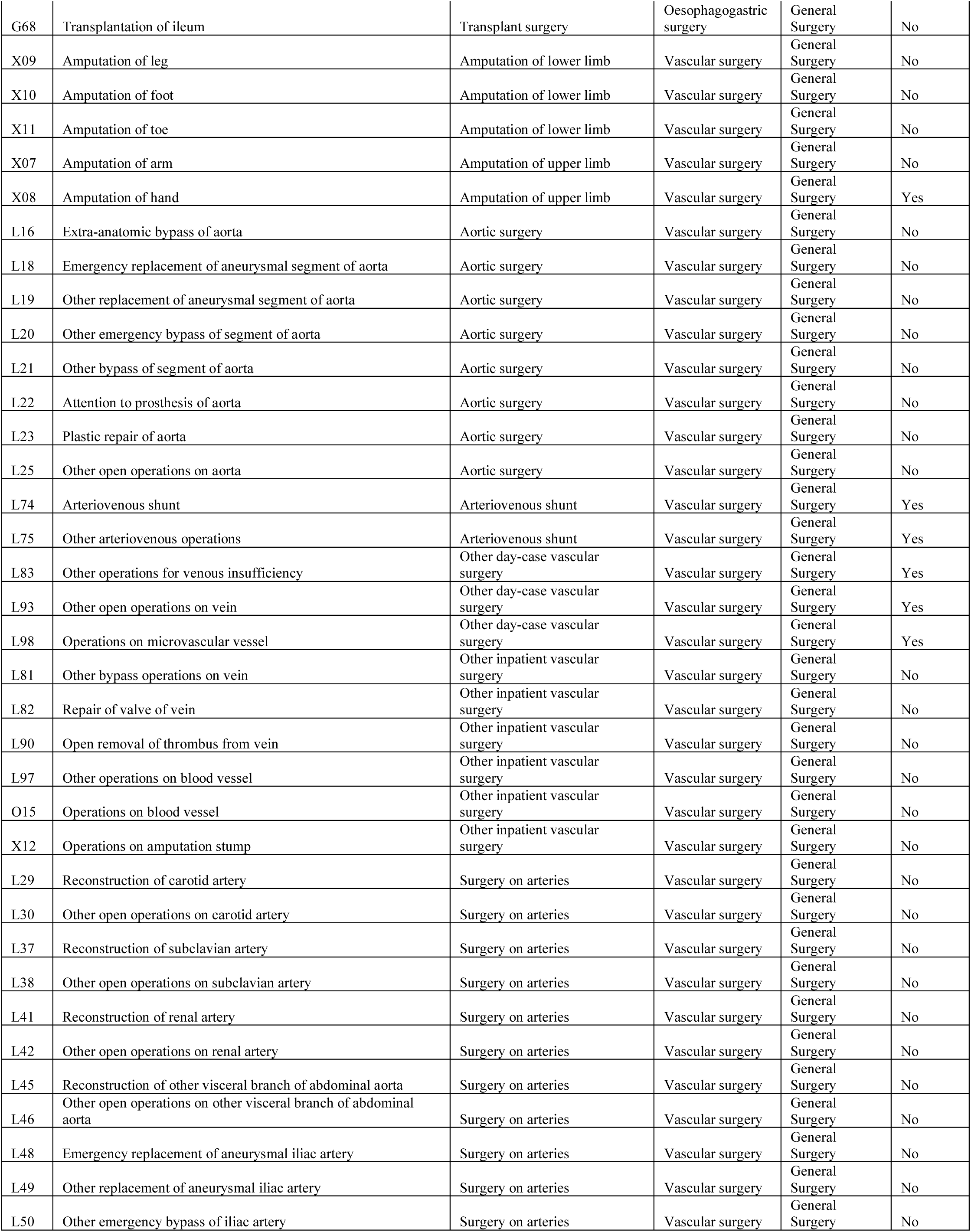
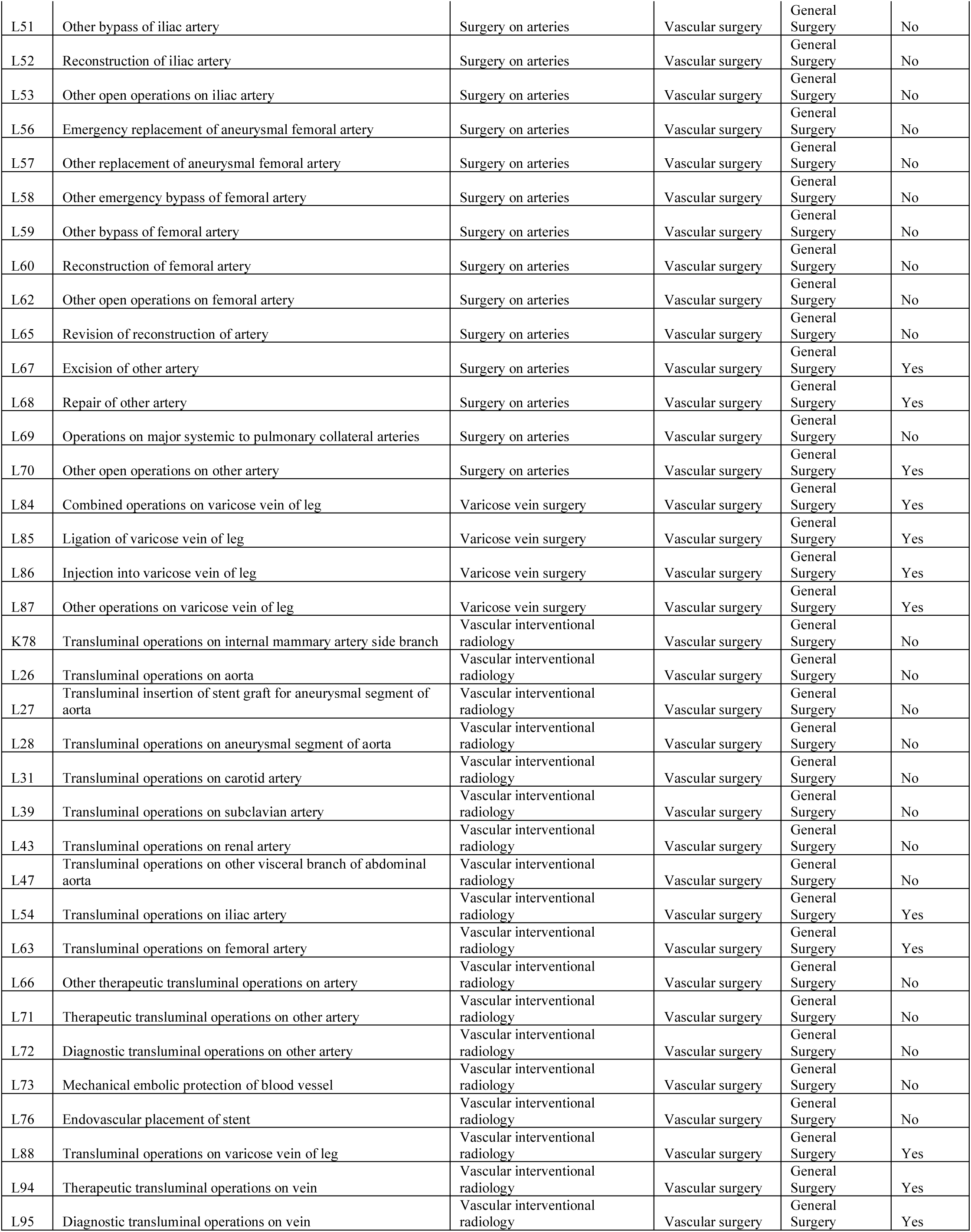
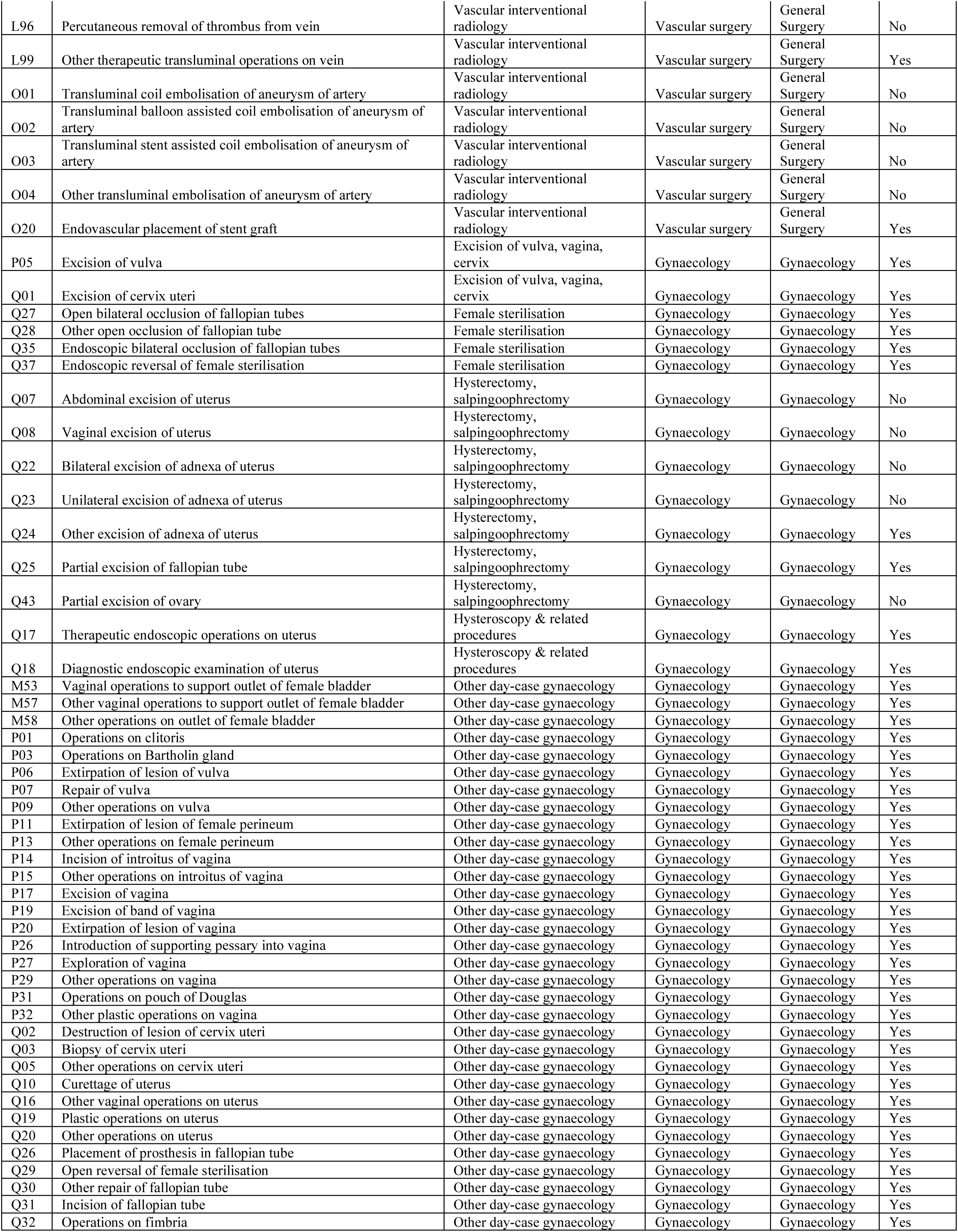
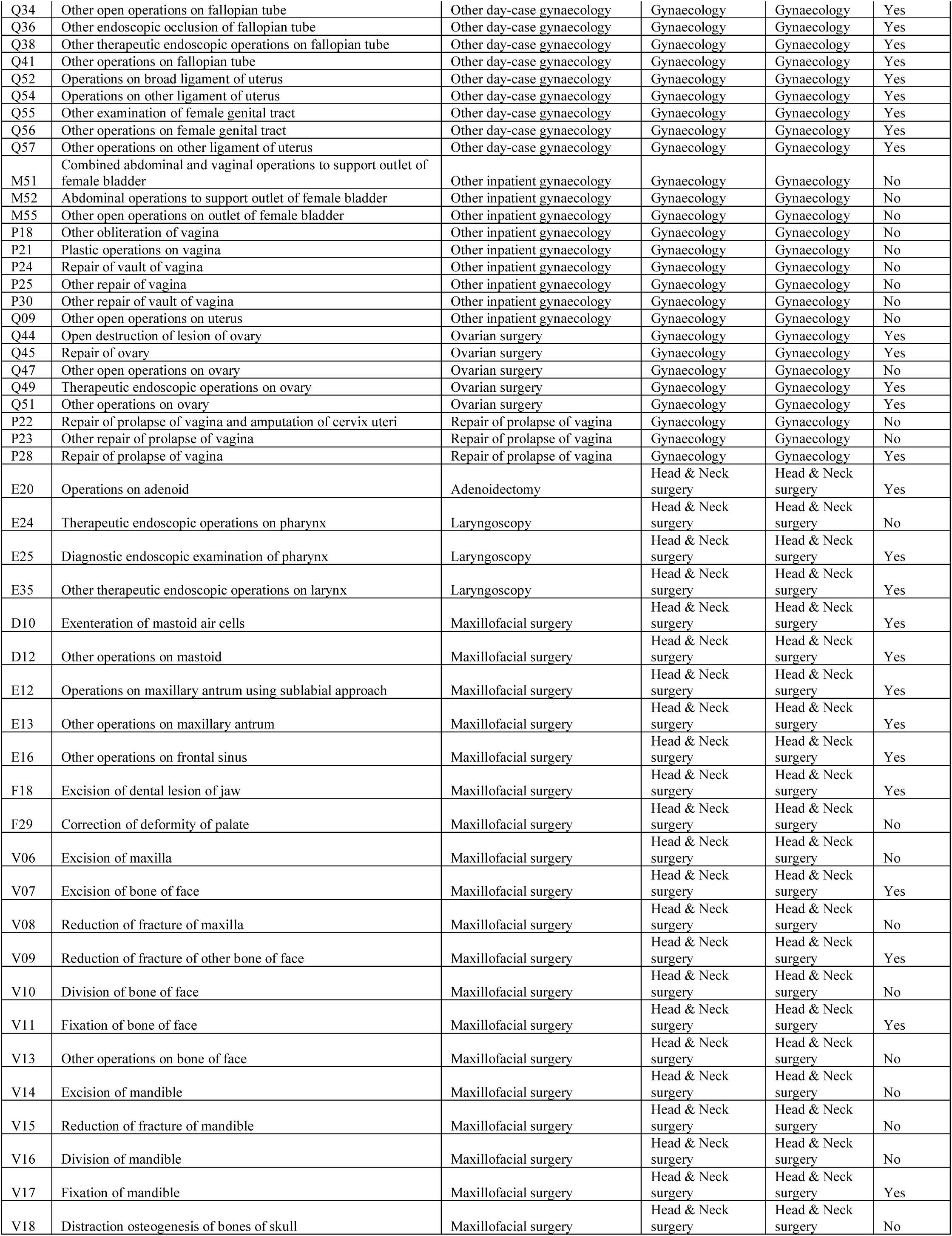
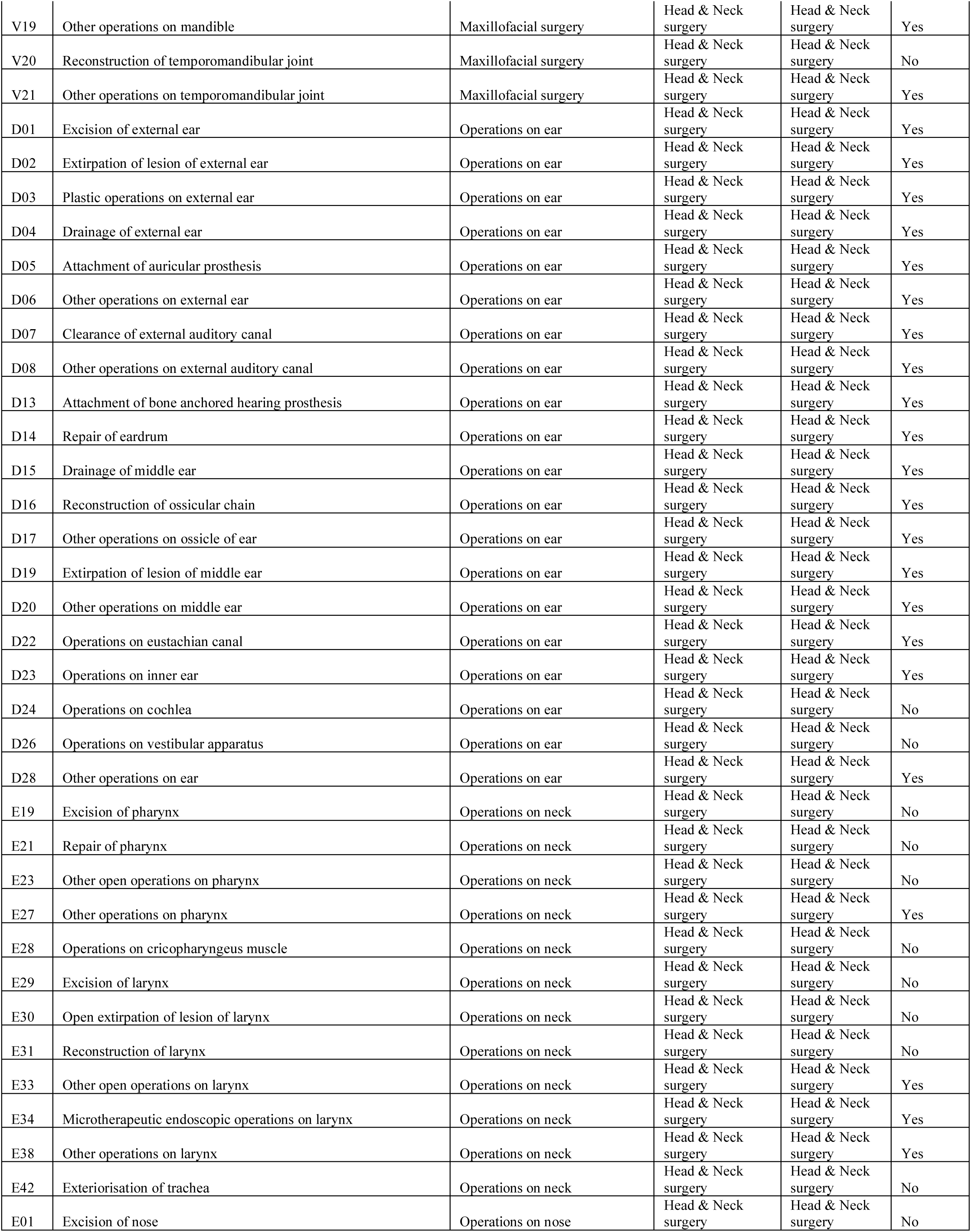
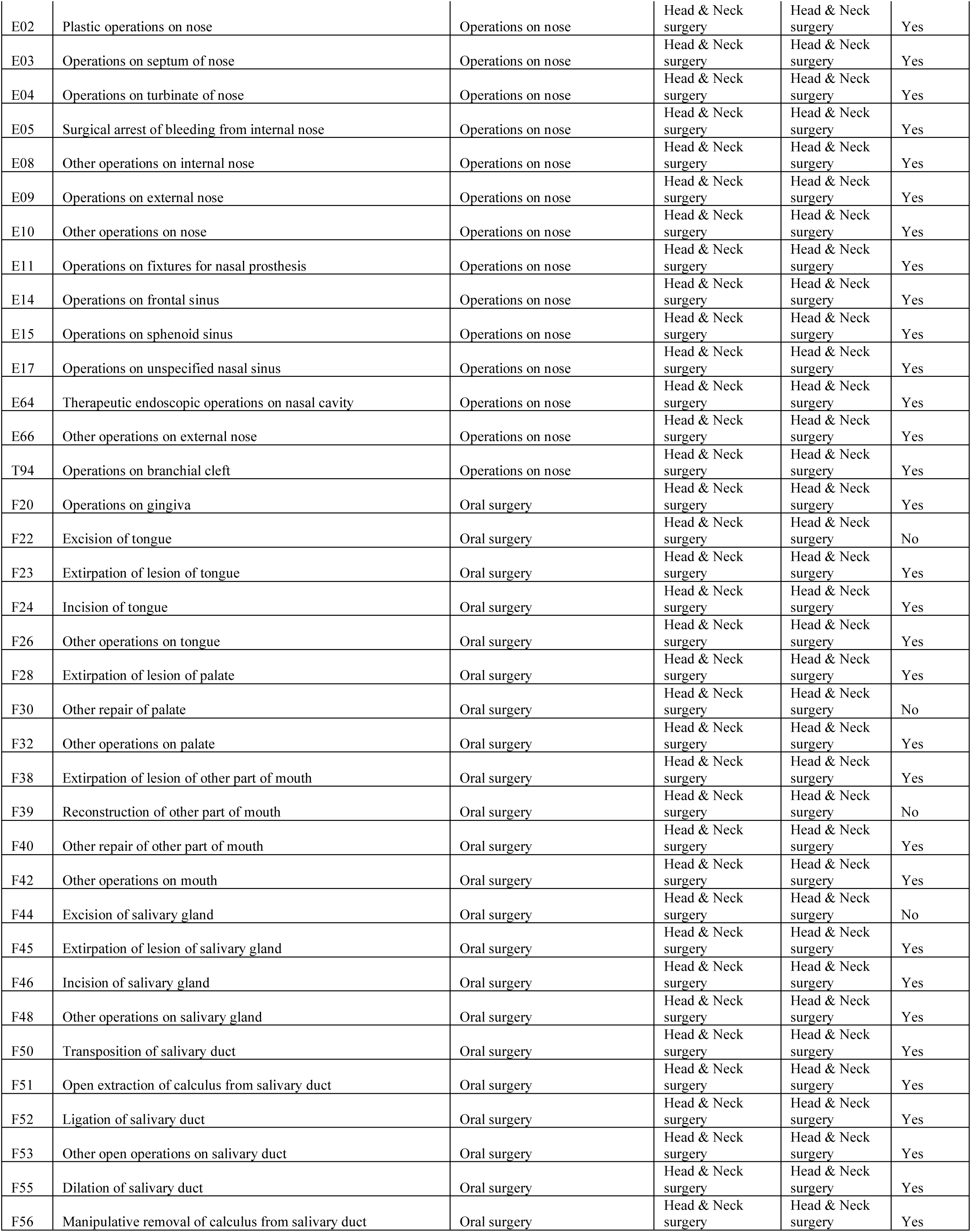
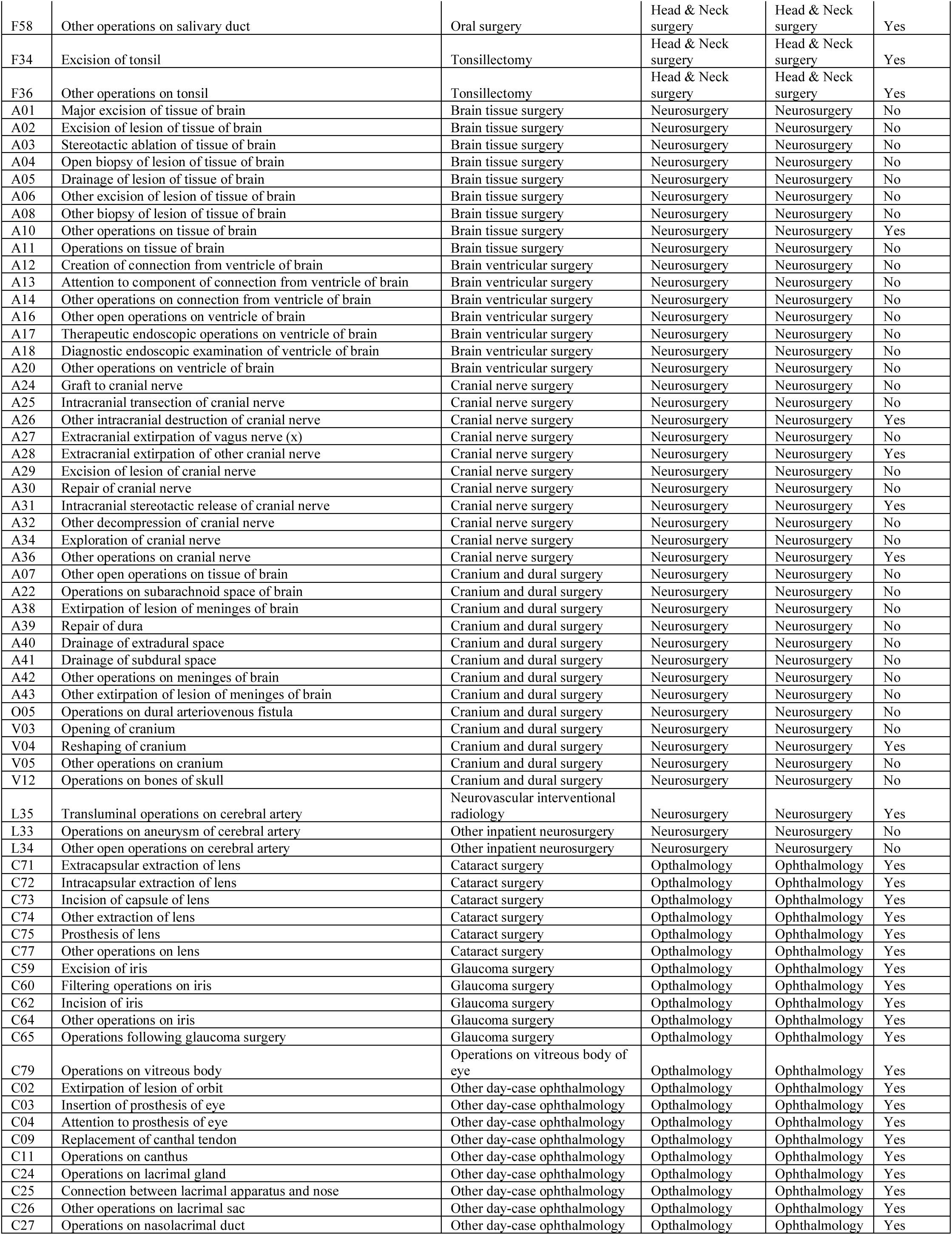
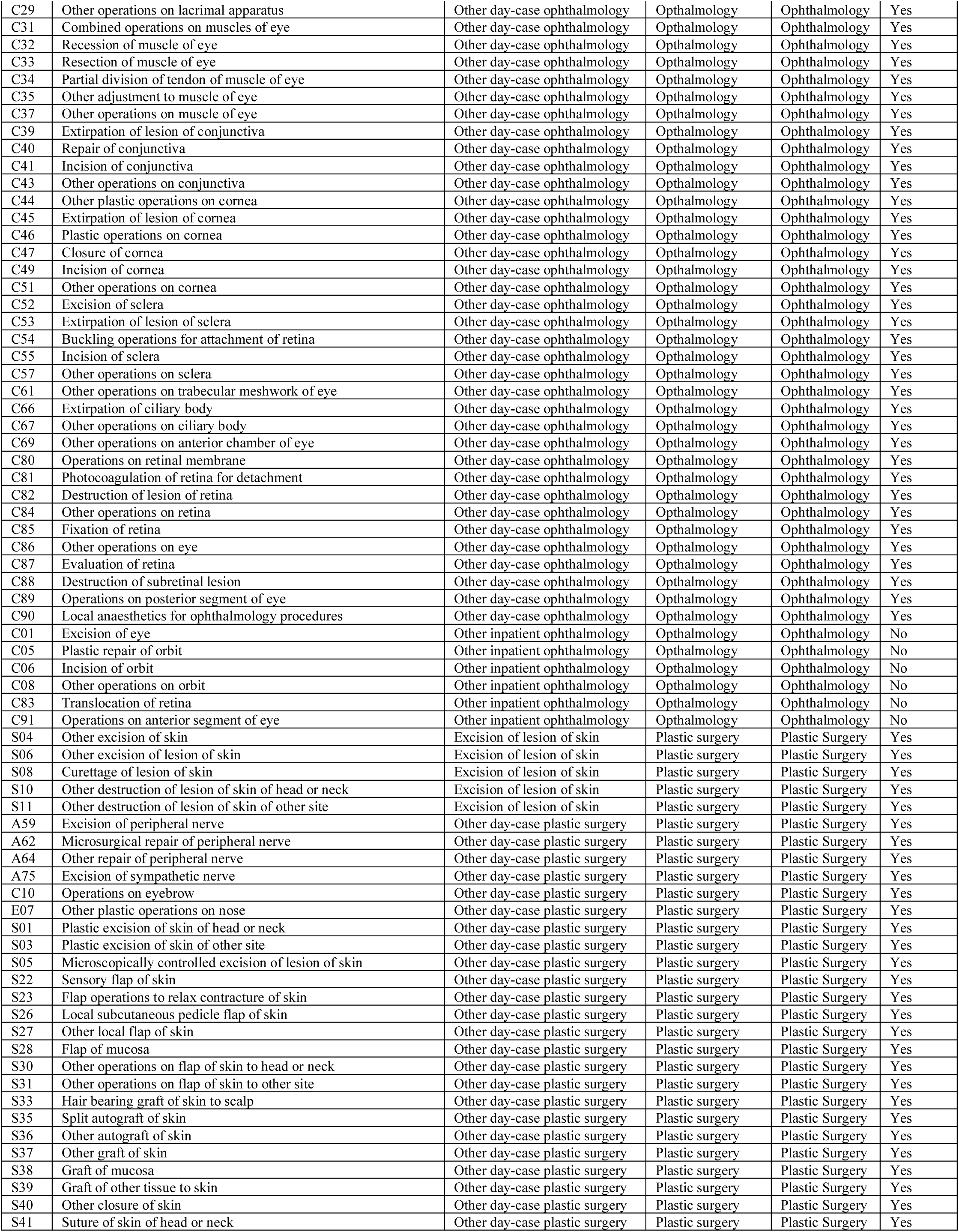
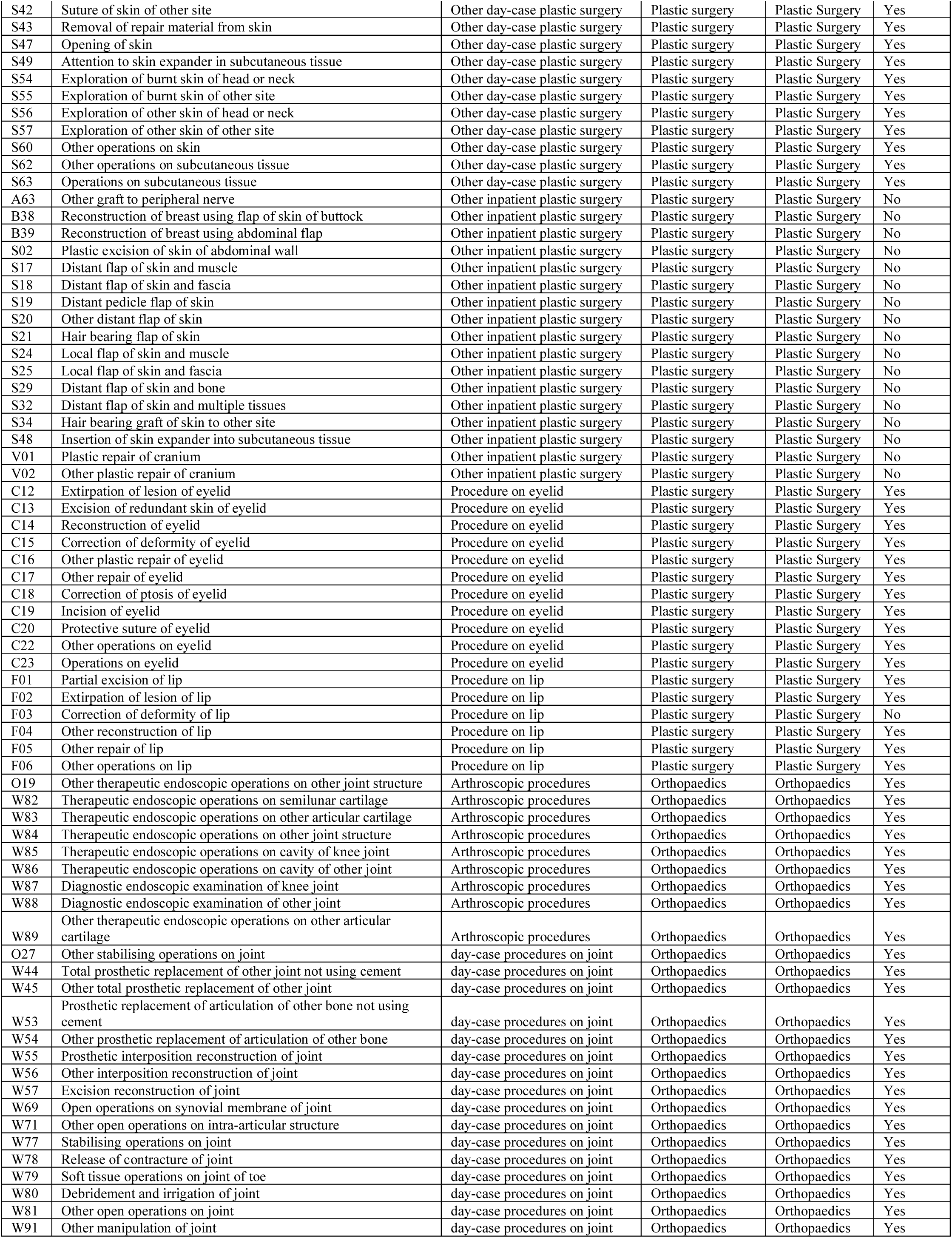
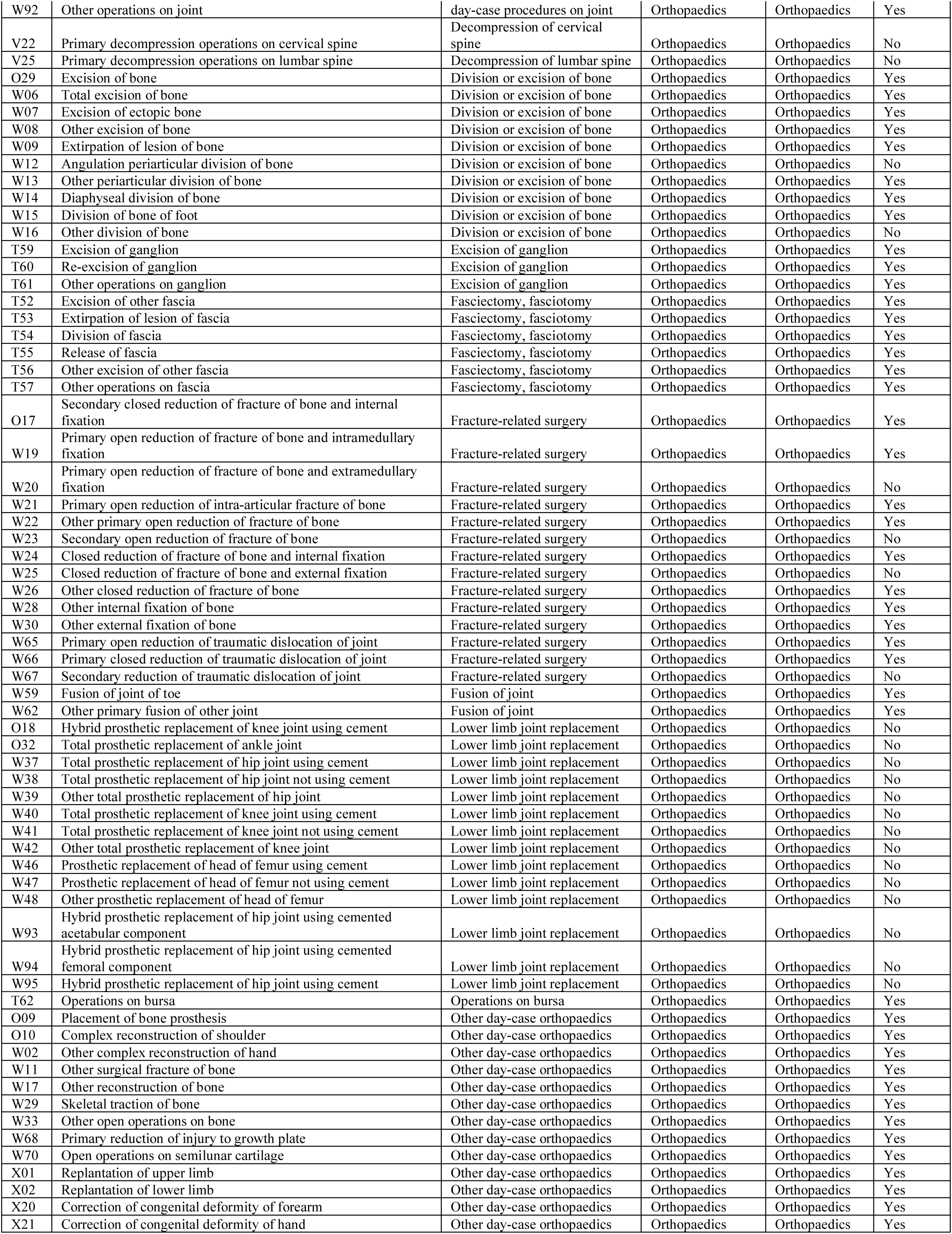
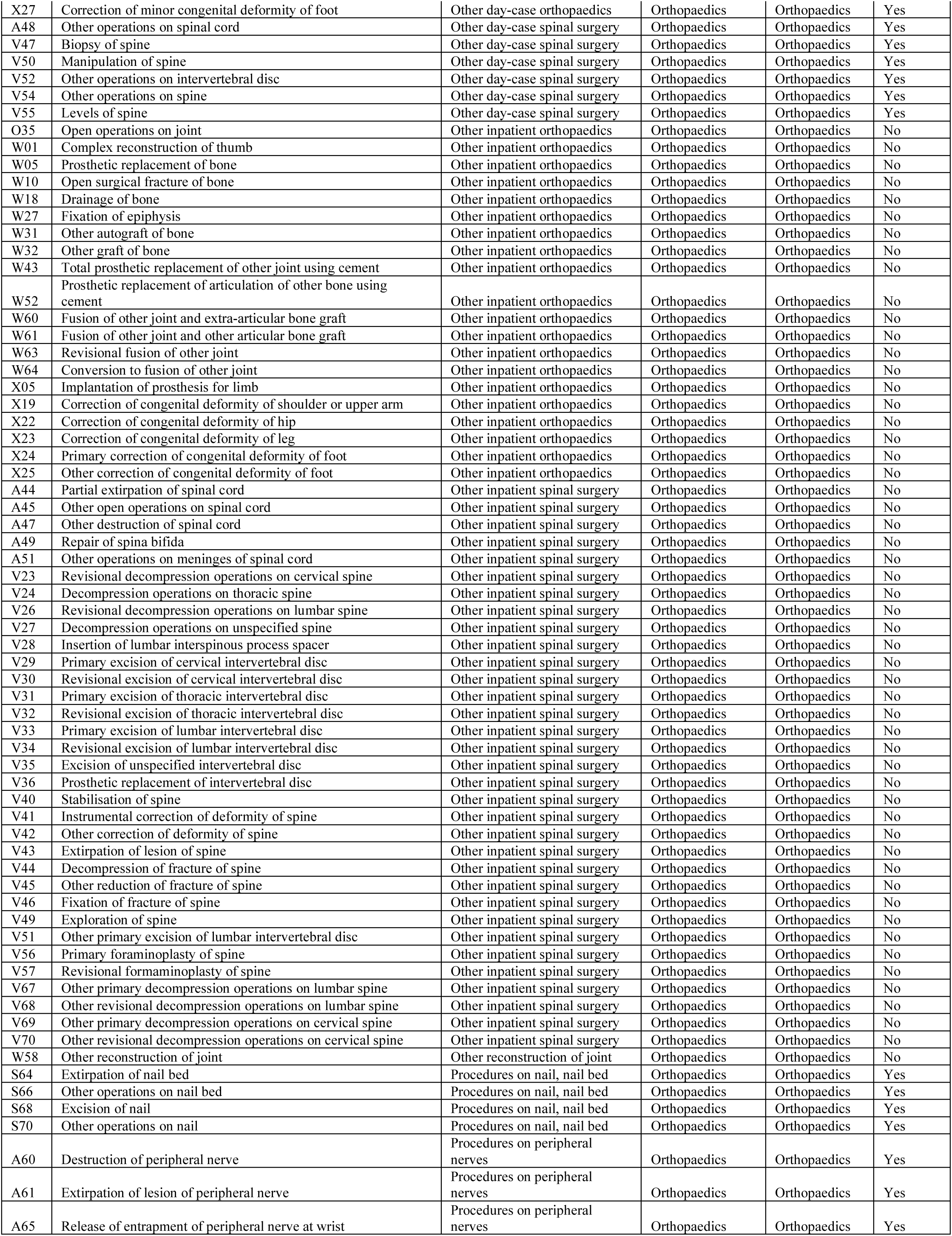
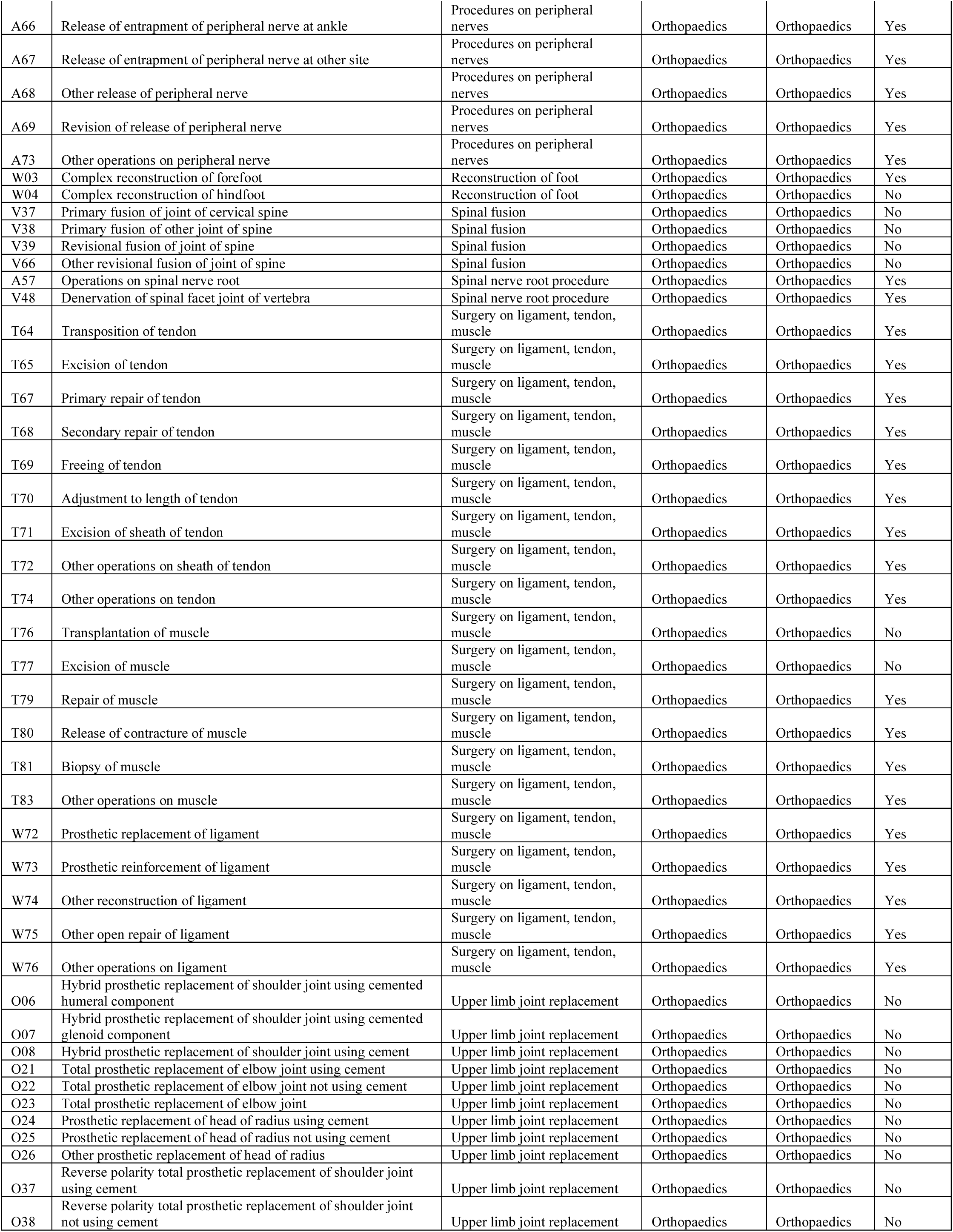
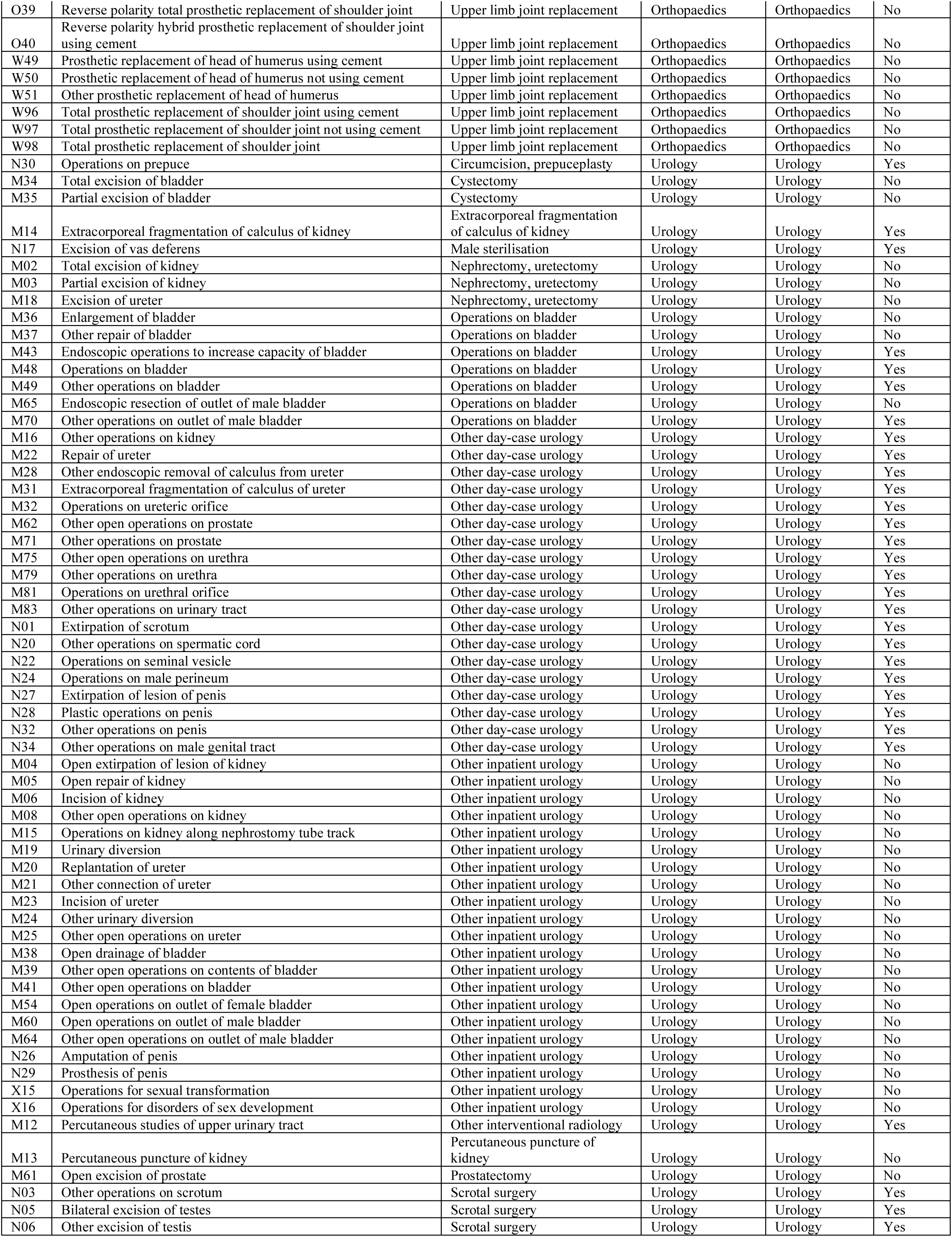
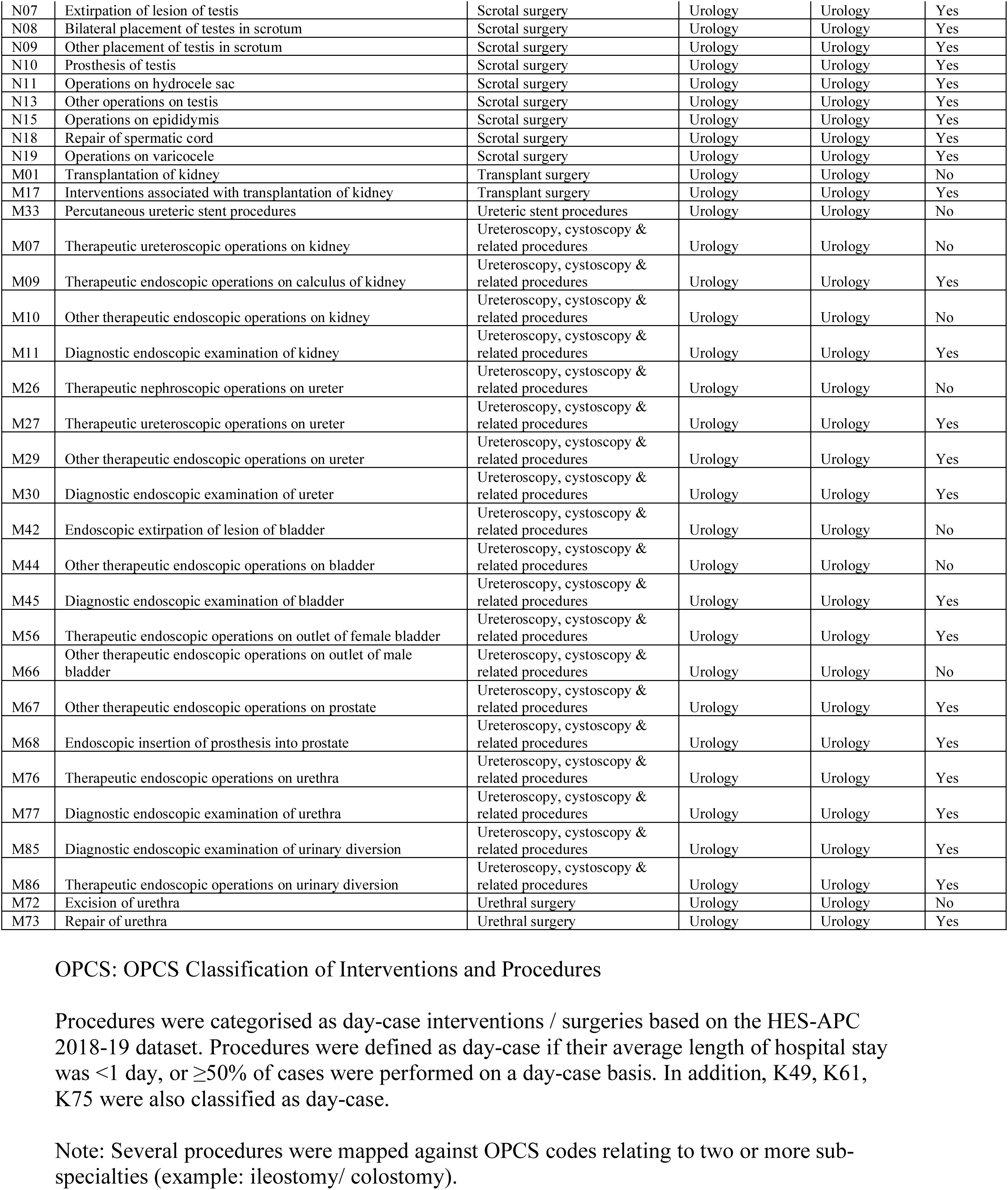
Categorisation of included OPCS codes

**Supplementary Table 2:** Baseline data taken from AHES-APC datasets

| OPCS code | Estimated number of elective procedures |  | Mean length of hospital stay in 2018-19 | Proportion of procedures completed as day-cases in 2018-19 | Average wait (days) on the waiting list for patients operated in 2018-19 |
| --- | --- | --- | --- | --- | --- |
|  | 2018-19 | 2020-21 |  |  |  |
| A01 | 216 | 130 | 7.7 | 0.5% | 72.4 |
| A02 | 3,963 | 3,531 | 6.9 | 0.2% | 23.1 |
| A03 | 39 | 16 | 3.3 | 0.0% | 63.5 |
| A04 | 526 | 387 | 6.0 | 9.9% | 13.3 |
| A05 | 139 | 127 | 17.5 | 0.7% | 29.9 |
| A06 | 27 | 22 | 3.9 | 11.2% | 65.0 |
| A07 | 98 | 69 | 14.4 | 1.0% | 95.9 |
| A08 | 642 | 536 | 5.4 | 10.0% | 10.9 |
| A10 | 773 | 460 | 5.3 | 71.9% | 19.3 |
| A11 | 502 | 318 | 10.4 | 0.6% | 61.6 |
| A12 | 1,376 | 928 | 10.6 | 1.0% | 59.0 |
| A13 | 356 | 298 | 5.3 | 7.6% | 34.9 |
| A14 | 819 | 1,107 | 5.9 | 31.4% | 36.9 |
| A16 | 93 | 116 | 11.6 | 0.0% | 38.8 |
| A17 | 306 | 274 | 7.3 | 0.3% | 40.6 |
| A18 | 14 | 9 | 8.2 | 0.0% | 20.6 |
| A20 | 614 | 380 | 13.1 | 13.7% | 62.5 |
| A22 | 26 | 9 | 9.8 | 38.0% | 33.8 |
| A24 | 38 | 12 | 5.4 | 7.9% | 116.8 |
| A25 | 8 | 4 | 4.6 | 12.5% | 139.6 |
| A26 | 427 | 267 | 0.9 | 70.5% | 66.9 |
| A27 | 5 | 5 | 4.3 | 41.7% | 24.3 |
| A28 | 54 | 30 | 1.1 | 76.0% | 54.8 |
| A29 | 374 | 254 | 8.5 | 4.3% | 87.3 |
| A30 | 20 | 16 | 2.8 | 29.6% | 66.9 |
| A31 | 48 | 14 | 1.0 | 64.6% | 30.4 |
| A32 | 511 | 268 | 3.9 | 2.9% | 106.3 |
| A34 | 8 | 0 | 3.3 | 12.9% | 66.3 |
| A36 | 557 | 438 | 0.5 | 84.7% | 85.1 |
| A38 | 1,303 | 1,182 | 8.6 | 0.2% | 49.7 |
| A39 | 192 | 93 | 7.0 | 2.1% | 86.4 |
| A40 | 55 | 22 | 10.9 | 0.0% | 121.8 |
| A41 | 973 | 663 | 10.7 | 0.4% | 15.7 |
| A42 | 47 | 41 | 7.4 | 10.6% | 43.8 |
| A43 | 205 | 174 | 11.5 | 0.5% | 56.0 |
| A44 | 612 | 491 | 9.0 | 0.3% | 46.5 |
| A45 | 84 | 30 | 11.1 | 4.8% | 64.0 |
| A47 | 27 | 21 | 2.0 | 22.2% | 18.3 |
| A48 | 3,869 | 1,847 | 0.9 | 73.9% | 86.9 |
| A49 | 278 | 173 | 7.6 | 1.1% | 99.6 |
| A51 | 305 | 236 | 8.3 | 19.7% | 98.4 |
| A57 | 48,974 | 19,695 | 0.4 | 98.0% | 104.8 |
| A59 | 417 | 178 | 0.7 | 84.8% | 98.2 |
| A60 | 5,976 | 3,802 | 0.2 | 98.5% | 135.1 |
| A61 | 4,052 | 1,539 | 0.3 | 88.4% | 92.4 |
| A62 | 2,453 | 2,110 | 0.7 | 87.6% | 10.2 |
| A63 | 36 | 23 | 2.1 | 42.2% | 47.5 |
| A64 | 960 | 727 | 0.4 | 91.4% | 5.2 |
| A65 | 44,564 | 20,323 | 0.0 | 99.0% | 72.3 |
| A66 | 117 | 43 | 0.4 | 80.3% | 86.0 |
| A67 | 4,739 | 2,200 | 0.1 | 93.0% | 86.3 |
| A68 | 1,197 | 533 | 0.5 | 78.3% | 86.1 |
| A69 | 1,479 | 604 | 0.1 | 95.6% | 91.7 |
| A73 | 37,046 | 16,201 | 0.3 | 97.6% | 108.8 |
| A75 | 170 | 41 | 1.5 | 56.0% | 78.9 |
| B01 | 147 | 71 | 6.4 | 0.0% | 55.6 |
| B02 | 0 | 0 | - | 0.0% | 0.0 |
| B04 | 1,116 | 878 | 5.7 | 0.3% | 65.0 |
| B06 | 26 | 29 | 6.6 | 0.0% | 25.3 |
| B08 | 11,210 | 7,799 | 1.7 | 5.6% | 72.2 |
| B09 | 19 | 9 | 2.3 | 0.0% | 33.8 |
| B10 | 860 | 388 | 1.1 | 28.6% | 102.8 |
| B12 | 2,246 | 1,532 | 1.2 | 88.6% | 27.3 |
| B14 | 4,751 | 2,753 | 1.2 | 23.8% | 107.9 |
| B16 | 85 | 65 | 1.1 | 29.4% | 103.7 |
| B17 | 4 | 9 | 17.3 | 0.0% | 1.0 |
| B18 | 411 | 339 | 4.1 | 1.0% | 33.7 |
| B20 | 25 | 24 | 7.5 | 28.0% | 49.2 |
| B22 | 870 | 633 | 3.6 | 1.3% | 62.5 |
| B23 | 2 | 4 | 1.0 | 50.0% | 5.0 |
| B25 | 219 | 185 | 3.1 | 56.6% | 17.0 |
| B27 | 17,937 | 15,009 | 1.8 | 19.6% | 33.3 |
| B28 | 42,943 | 32,922 | 0.2 | 80.4% | 25.3 |
| B29 | 1,416 | 392 | 1.9 | 34.6% | 123.6 |
| B30 | 3,639 | 1,575 | 1.0 | 53.9% | 91.7 |
| B31 | 3,963 | 1,544 | 0.9 | 36.1% | 106.2 |
| B32 | 4,806 | 3,136 | 1.6 | 90.4% | 19.5 |
| B33 | 700 | 384 | 1.6 | 62.1% | 47.4 |
| B34 | 2,511 | 1,671 | 0.1 | 93.1% | 36.0 |
| B35 | 1,165 | 535 | 0.1 | 96.0% | 46.6 |
| B36 | 2,859 | 727 | 0.1 | 94.5% | 121.3 |
| B37 | 4,958 | 2,428 | 0.7 | 79.3% | 119.1 |
| B38 | 7 | 4 | 3.6 | 28.6% | 19.8 |
| B39 | 1,032 | 284 | 5.0 | 2.0% | 229.7 |
| B40 | 9 | 0 | 0.0 | 100.0% | 59.6 |
| B41 | 986 | 615 | 0.2 | 80.9% | 23.5 |
| C01 | 679 | 497 | 2.6 | 17.4% | 47.3 |
| C02 | 268 | 153 | 0.9 | 64.3% | 74.4 |
| C03 | 129 | 58 | 0.4 | 71.3% | 96.0 |
| C04 | 119 | 57 | 0.6 | 70.7% | 70.9 |
| C05 | 262 | 100 | 1.0 | 41.7% | 61.1 |
| C06 | 688 | 410 | 1.6 | 39.9% | 68.5 |
| C08 | 690 | 292 | 1.2 | 26.7% | 25.1 |
| C09 | 436 | 232 | 0.0 | 97.0% | 98.5 |
| C10 | 3,814 | 2,491 | 0.3 | 96.6% | 66.7 |
| C11 | 6,113 | 3,919 | 0.1 | 97.8% | 66.7 |
| C12 | 20,984 | 10,211 | 0.0 | 98.8% | 60.8 |
| C13 | 3,396 | 1,580 | 0.0 | 98.2% | 100.9 |
| C14 | 2,010 | 1,287 | 0.2 | 94.2% | 63.8 |
| C15 | 12,855 | 6,635 | 0.0 | 98.7% | 83.7 |
| C16 | 761 | 422 | 1.0 | 93.4% | 55.2 |
| C17 | 731 | 506 | 0.5 | 93.7% | 67.6 |
| C18 | 5,349 | 2,406 | 0.0 | 98.6% | 111.3 |
| C19 | 504 | 193 | 0.5 | 96.4% | 57.1 |
| C20 | 86 | 43 | 1.7 | 88.5% | 56.7 |
| C22 | 6,730 | 3,142 | 0.1 | 99.2% | 58.5 |
| C23 | 253 | 113 | 0.1 | 96.5% | 97.3 |
| C24 | 301 | 197 | 0.3 | 87.1% | 63.2 |
| C25 | 3,617 | 1,016 | 0.2 | 80.5% | 127.4 |
| C26 | 102 | 47 | 0.5 | 84.6% | 74.4 |
| C27 | 2,880 | 793 | 0.0 | 98.2% | 90.2 |
| C29 | 3,688 | 1,211 | 0.0 | 99.3% | 76.4 |
| C31 | 6,848 | 2,536 | 0.0 | 96.7% | 126.2 |
| C32 | 2,465 | 881 | 0.0 | 96.9% | 121.0 |
| C33 | 1,066 | 357 | 0.0 | 97.6% | 114.0 |
| C34 | 250 | 82 | 0.0 | 98.8% | 95.3 |
| C35 | 312 | 128 | 0.0 | 97.8% | 109.9 |
| C37 | 816 | 379 | 0.1 | 97.9% | 100.0 |
| C39 | 2,710 | 1,193 | 0.1 | 96.9% | 71.6 |
| C40 | 386 | 215 | 0.4 | 90.4% | 62.5 |
| C41 | 44 | 18 | 0.5 | 91.0% | 80.7 |
| C43 | 1,166 | 620 | 0.2 | 98.3% | 44.4 |
| C44 | 2,569 | 1,498 | 0.4 | 71.9% | 113.1 |
| C45 | 1,557 | 883 | 0.3 | 97.5% | 80.6 |
| C46 | 3,462 | 1,910 | 0.7 | 77.1% | 97.9 |
| C47 | 1,787 | 1,080 | 0.6 | 96.8% | 59.7 |
| C49 | 203 | 106 | 0.0 | 97.5% | 78.7 |
| C51 | 3,002 | 1,952 | 0.6 | 97.8% | 80.2 |
| C52 | 465 | 321 | 0.0 | 96.8% | 59.8 |
| C53 | 25 | 8 | 0.0 | 96.0% | 67.3 |
| C54 | 716 | 553 | 0.2 | 85.3% | 27.1 |
| C55 | 39 | 33 | 1.2 | 74.7% | 27.6 |
| C57 | 249 | 182 | 0.7 | 84.9% | 27.9 |
| C59 | 116 | 76 | 0.4 | 88.1% | 47.0 |
| C60 | 9,246 | 6,723 | 0.1 | 92.3% | 65.5 |
| C61 | 2,573 | 2,117 | 0.0 | 97.6% | 66.7 |
| C62 | 2,936 | 1,796 | 0.2 | 98.3% | 62.0 |
| C64 | 229 | 150 | 0.1 | 95.8% | 65.4 |
| C65 | 3,619 | 2,356 | 0.0 | 97.2% | 40.1 |
| C66 | 2,782 | 2,394 | 0.1 | 98.4% | 36.4 |
| C67 | 39 | 66 | 1.3 | 82.1% | 38.0 |
| C69 | 915 | 567 | 0.8 | 90.6% | 24.2 |
| C71 | 3,811 | 1,260 | 0.1 | 98.5% | 51.3 |
| C72 | 29 | 15 | 0.0 | 96.6% | 92.6 |
| C73 | 17,893 | 18,361 | 0.0 | 99.6% | 66.1 |
| C74 | 278 | 242 | 0.7 | 75.9% | 37.4 |
| C75 | 423,494 | 233,483 | 0.0 | 99.4% | 81.6 |
| C77 | 150 | 65 | 0.0 | 97.6% | 55.1 |
| C79 | 106,809 | 98,757 | 0.1 | 97.4% | 41.1 |
| C80 | 1,208 | 917 | 0.2 | 88.2% | 60.9 |
| C81 | 857 | 578 | 0.2 | 93.5% | 26.9 |
| C82 | 8,536 | 6,158 | 0.3 | 96.8% | 45.9 |
| C83 | 0 | 2 | - | 0.0% | 0.0 |
| C84 | 241 | 149 | 0.7 | 66.4% | 31.6 |
| C85 | 1,811 | 1,099 | 0.1 | 93.5% | 23.9 |
| C86 | 4,117 | 1,887 | 0.3 | 97.4% | 67.3 |
| C87 | 1,058 | 1,571 | 0.4 | 97.7% | 61.5 |
| C88 | 59 | 38 | 0.0 | 100.0% | 4.3 |
| C89 | 2,247 | 1,953 | 0.0 | 99.2% | 38.2 |
| C90 | 39 | 21 | 0.0 | 97.4% | 97.9 |
| C91 | 0 | 4 | - | 0.0% | 0.0 |
| D01 | 2,088 | 1,178 | 0.3 | 93.3% | 71.3 |
| D02 | 16,307 | 11,422 | 0.1 | 98.2% | 53.7 |
| D03 | 1,584 | 528 | 0.4 | 80.9% | 115.8 |
| D04 | 172 | 83 | 1.4 | 79.8% | 19.0 |
| D05 | 31 | 12 | 2.3 | 90.6% | 75.2 |
| D06 | 3,503 | 2,114 | 0.5 | 96.8% | 39.6 |
| D07 | 3,849 | 1,688 | 1.4 | 93.0% | 32.9 |
| D08 | 940 | 333 | 1.8 | 81.9% | 69.4 |
| D10 | 3,076 | 1,419 | 1.2 | 53.8% | 123.9 |
| D12 | 1,191 | 525 | 0.9 | 64.3% | 125.6 |
| D13 | 1,930 | 560 | 0.1 | 94.2% | 95.0 |
| D14 | 8,614 | 3,191 | 0.3 | 78.8% | 126.2 |
| D15 | 23,216 | 4,385 | 0.3 | 95.3% | 86.4 |
| D16 | 1,334 | 419 | 0.2 | 78.9% | 133.6 |
| D17 | 682 | 219 | 0.3 | 72.0% | 133.7 |
| D19 | 797 | 326 | 0.7 | 81.3% | 81.2 |
| D20 | 2,986 | 847 | 0.3 | 96.1% | 69.0 |
| D22 | 399 | 108 | 0.2 | 95.5% | 114.2 |
| D23 | 184 | 35 | 0.2 | 91.3% | 61.2 |
| D24 | 1,392 | 823 | 1.0 | 38.3% | 65.8 |
| D26 | 97 | 26 | 1.3 | 43.3% | 114.6 |
| D28 | 1,263 | 425 | 0.3 | 96.1% | 70.9 |
| E01 | 101 | 107 | 4.0 | 13.9% | 35.5 |
| E02 | 3,180 | 1,086 | 0.5 | 65.3% | 132.9 |
| E03 | 19,887 | 5,878 | 0.2 | 85.2% | 98.4 |
| E04 | 4,424 | 1,431 | 0.2 | 88.0% | 107.3 |
| E05 | 2,338 | 808 | 1.7 | 82.2% | 71.9 |
| E07 | 2,382 | 546 | 0.3 | 77.8% | 143.8 |
| E08 | 7,490 | 2,928 | 0.4 | 81.4% | 86.5 |
| E09 | 27,351 | 19,272 | 0.1 | 97.8% | 64.4 |
| E10 | 605 | 363 | 0.3 | 95.9% | 41.1 |
| E11 | 27 | 26 | 0.4 | 77.8% | 64.1 |
| E12 | 86 | 36 | 0.8 | 80.3% | 75.7 |
| E13 | 4,944 | 2,072 | 0.5 | 79.1% | 96.8 |
| E14 | 3,283 | 1,279 | 0.6 | 75.0% | 107.6 |
| E15 | 363 | 182 | 1.8 | 60.0% | 92.2 |
| E16 | 158 | 69 | 1.9 | 59.6% | 89.3 |
| E17 | 759 | 431 | 0.7 | 76.3% | 86.5 |
| E19 | 361 | 335 | 17.6 | 4.1% | 20.5 |
| E20 | 6,155 | 1,454 | 0.2 | 87.7% | 99.9 |
| E21 | 166 | 61 | 2.3 | 6.0% | 135.9 |
| E23 | 331 | 144 | 5.4 | 23.0% | 80.3 |
| E24 | 1,076 | 502 | 2.0 | 40.4% | 73.3 |
| E25 | 7,539 | 4,549 | 1.2 | 84.0% | 29.1 |
| E27 | 650 | 376 | 2.9 | 71.8% | 46.4 |
| E28 | 260 | 119 | 2.0 | 33.9% | 90.1 |
| E29 | 369 | 359 | 21.2 | 7.0% | 22.9 |
| E30 | 47 | 19 | 3.8 | 42.9% | 55.1 |
| E31 | 141 | 91 | 6.1 | 20.6% | 56.9 |
| E33 | 566 | 251 | 1.1 | 75.1% | 81.0 |
| E34 | 5,286 | 3,020 | 0.8 | 75.3% | 44.9 |
| E35 | 1,893 | 1,157 | 1.7 | 64.7% | 48.6 |
| E38 | 1,005 | 624 | 0.2 | 94.7% | 64.7 |
| E39 | 34 | 36 | 6.6 | 32.1% | 59.9 |
| E40 | 25 | 16 | 7.5 | 8.0% | 76.5 |
| E41 | 232 | 151 | 3.8 | 42.7% | 50.3 |
| E42 | 1,992 | 1,813 | 19.0 | 26.1% | 44.7 |
| E43 | 158 | 84 | 4.9 | 24.7% | 74.0 |
| E44 | 0 | 2 | - | 0.0% | 0.0 |
| E46 | 76 | 58 | 6.2 | 5.3% | 32.6 |
| E47 | 33 | 26 | 13.8 | 27.4% | 14.1 |
| E48 | 1,270 | 819 | 2.9 | 41.1% | 22.4 |
| E49 | 36,042 | 15,554 | 1.5 | 81.4% | 15.4 |
| E50 | 804 | 607 | 3.1 | 33.7% | 25.4 |
| E51 | 1,200 | 727 | 2.0 | 46.9% | 50.0 |
| E52 | 170 | 92 | 4.2 | 10.6% | 26.7 |
| E53 | 52 | 32 | 37.6 | 1.9% | 15.5 |
| E54 | 10,201 | 7,986 | 6.5 | 0.1% | 22.6 |
| E55 | 1,809 | 1,166 | 6.1 | 0.1% | 33.8 |
| E57 | 51 | 25 | 7.4 | 5.9% | 25.2 |
| E59 | 12,095 | 9,925 | 1.0 | 76.9% | 12.2 |
| E61 | 463 | 394 | 5.6 | 11.9% | 29.4 |
| E62 | 80 | 57 | 2.6 | 10.0% | 45.8 |
| E63 | 16,673 | 13,000 | 0.6 | 88.8% | 10.6 |
| E64 | 877 | 689 | 0.4 | 80.4% | 96.1 |
| E66 | 366 | 329 | 0.3 | 85.8% | 64.1 |
| E67 | 29 | 2 | 3.4 | 62.1% | 27.9 |
| F01 | 124 | 87 | 1.3 | 83.1% | 46.7 |
| F02 | 9,189 | 5,182 | 0.1 | 97.6% | 56.3 |
| F03 | 663 | 374 | 1.2 | 20.7% | 105.4 |
| F04 | 146 | 83 | 0.9 | 72.1% | 74.3 |
| F05 | 1,438 | 804 | 0.3 | 94.0% | 61.7 |
| F06 | 2,832 | 1,558 | 0.2 | 97.8% | 40.0 |
| F18 | 3,095 | 1,621 | 0.2 | 90.7% | 78.4 |
| F20 | 3,170 | 1,899 | 0.1 | 97.4% | 43.9 |
| F22 | 1,014 | 899 | 8.2 | 10.2% | 27.5 |
| F23 | 4,035 | 2,435 | 0.4 | 84.5% | 40.9 |
| F24 | 6,717 | 3,968 | 0.2 | 94.7% | 25.0 |
| F26 | 4,566 | 2,009 | 1.0 | 71.0% | 68.4 |
| F28 | 1,682 | 907 | 0.6 | 86.7% | 49.6 |
| F29 | 1,178 | 688 | 1.5 | 3.5% | 127.4 |
| F30 | 240 | 98 | 1.6 | 19.6% | 145.0 |
| F32 | 2,734 | 1,219 | 0.2 | 91.5% | 48.5 |
| F34 | 45,021 | 15,636 | 0.4 | 65.9% | 96.2 |
| F36 | 1,793 | 1,150 | 1.1 | 81.0% | 29.4 |
| F38 | 4,178 | 2,112 | 1.3 | 82.5% | 46.3 |
| F39 | 74 | 44 | 4.7 | 37.9% | 87.3 |
| F40 | 108 | 50 | 0.8 | 85.3% | 72.6 |
| F42 | 8,050 | 3,703 | 0.4 | 97.0% | 35.3 |
| F44 | 4,332 | 2,484 | 1.8 | 12.2% | 81.0 |
| F45 | 1,072 | 547 | 0.8 | 44.5% | 77.3 |
| F46 | 55 | 34 | 3.6 | 65.0% | 83.3 |
| F48 | 1,742 | 1,193 | 1.3 | 92.7% | 44.7 |
| F50 | 14 | 7 | 0.5 | 50.0% | 131.8 |
| F51 | 535 | 196 | 0.2 | 88.1% | 86.9 |
| F52 | 44 | 14 | 0.8 | 65.9% | 127.3 |
| F53 | 77 | 50 | 1.1 | 79.1% | 86.5 |
| F55 | 285 | 86 | 0.1 | 93.7% | 114.2 |
| F56 | 134 | 39 | 0.3 | 92.6% | 80.8 |
| F58 | 171 | 103 | 2.2 | 84.9% | 92.5 |
| G01 | 1,403 | 1,008 | 12.7 | 0.1% | 31.2 |
| G02 | 87 | 62 | 12.6 | 0.0% | 22.9 |
| G03 | 212 | 163 | 12.9 | 1.9% | 25.7 |
| G04 | 27 | 21 | 5.5 | 3.7% | 72.8 |
| G05 | 18 | 12 | 11.9 | 0.0% | 57.5 |
| G06 | 25 | 11 | 12.8 | 27.9% | 63.1 |
| G07 | 169 | 137 | 18.8 | 5.9% | 38.6 |
| G08 | 16 | 16 | 16.3 | 30.8% | 35.4 |
| G09 | 531 | 311 | 2.9 | 5.1% | 98.5 |
| G10 | 38 | 16 | 1.3 | 84.3% | 37.5 |
| G11 | 14 | 10 | 8.6 | 21.4% | 12.5 |
| G12 | 63 | 59 | 0.5 | 79.4% | 29.3 |
| G13 | 8 | 7 | 4.4 | 12.3% | 21.8 |
| G14 | 426 | 214 | 0.7 | 83.3% | 50.7 |
| G15 | 4,876 | 3,885 | 2.1 | 65.5% | 32.6 |
| G16 | 9,655 | 6,265 | 0.9 | 86.8% | 28.3 |
| G17 | 15 | 7 | 0.9 | 73.3% | 32.5 |
| G18 | 462 | 257 | 1.0 | 61.6% | 55.8 |
| G19 | 512 | 265 | 1.0 | 83.2% | 39.3 |
| G20 | 7 | 23 | 3.7 | 27.8% | 7.5 |
| G21 | 9,211 | 4,318 | 0.8 | 86.4% | 43.9 |
| G23 | 1,975 | 990 | 3.7 | 7.0% | 123.8 |
| G24 | 2,628 | 960 | 3.0 | 13.1% | 129.3 |
| G25 | 219 | 114 | 3.6 | 12.4% | 115.1 |
| G26 | 1 | 0 | 49.7 | 0.0% | 0.0 |
| G27 | 349 | 324 | 11.3 | 0.3% | 24.9 |
| G28 | 3,307 | 1,283 | 3.9 | 0.7% | 118.9 |
| G29 | 296 | 236 | 4.2 | 4.4% | 41.9 |
| G30 | 1,384 | 362 | 0.9 | 59.0% | 82.1 |
| G31 | 30 | 23 | 9.6 | 6.7% | 143.4 |
| G32 | 1,840 | 626 | 2.6 | 3.2% | 135.4 |
| G33 | 2,363 | 913 | 5.8 | 6.6% | 101.1 |
| G34 | 7,423 | 5,754 | 6.0 | 47.2% | 42.1 |
| G35 | 8 | 7 | 8.1 | 0.0% | 79.3 |
| G36 | 90 | 47 | 8.1 | 10.0% | 82.3 |
| G38 | 932 | 592 | 2.8 | 47.7% | 86.8 |
| G40 | 178 | 174 | 3.7 | 4.5% | 48.1 |
| G41 | 13 | 14 | 8.8 | 46.4% | 24.3 |
| G42 | 1,965 | 1,527 | 0.3 | 80.3% | 40.4 |
| G43 | 18,182 | 11,949 | 2.1 | 69.5% | 37.2 |
| G44 | 26,398 | 18,781 | 4.6 | 59.4% | 32.8 |
| G45 | 685,821 | 415,882 | 0.5 | 91.8% | 28.5 |
| G46 | 402 | 352 | 4.1 | 41.8% | 32.7 |
| G48 | 515 | 231 | 1.3 | 77.0% | 77.4 |
| G49 | 55 | 59 | 15.0 | 10.9% | 31.9 |
| G50 | 39 | 26 | 8.2 | 10.2% | 42.0 |
| G51 | 96 | 80 | 14.3 | 1.0% | 49.4 |
| G52 | 5 | 9 | 8.7 | 0.0% | 9.0 |
| G53 | 109 | 94 | 11.2 | 4.6% | 62.4 |
| G54 | 327 | 270 | 4.8 | 51.6% | 26.8 |
| G55 | 283 | 148 | 0.7 | 73.4% | 25.6 |
| G57 | 37 | 16 | 5.1 | 35.1% | 29.7 |
| G58 | 205 | 215 | 13.0 | 1.9% | 55.4 |
| G59 | 20 | 22 | 9.5 | 10.2% | 32.5 |
| G60 | 1,045 | 1,059 | 8.9 | 43.3% | 33.0 |
| G61 | 59 | 39 | 10.2 | 3.4% | 99.9 |
| G62 | 4 | 2 | 0.5 | 75.0% | 72.0 |
| G63 | 68 | 19 | 10.8 | 36.7% | 28.1 |
| G64 | 1,658 | 449 | 5.9 | 38.2% | 18.1 |
| G65 | 199 | 134 | 1.5 | 74.8% | 49.8 |
| G67 | 978 | 2,149 | 6.6 | 56.6% | 31.8 |
| G68 | 1 | 0 | 22.1 | 0.0% | 0.0 |
| G69 | 1,403 | 1,011 | 13.2 | 1.1% | 60.3 |
| G70 | 128 | 89 | 7.0 | 10.9% | 60.9 |
| G71 | 64 | 45 | 13.1 | 0.0% | 51.7 |
| G72 | 146 | 71 | 12.1 | 0.0% | 130.1 |
| G73 | 353 | 291 | 11.9 | 4.5% | 72.8 |
| G74 | 1,045 | 748 | 14.3 | 0.1% | 59.1 |
| G75 | 5,015 | 2,628 | 7.4 | 2.6% | 90.0 |
| G76 | 48 | 24 | 8.5 | 0.0% | 64.5 |
| G78 | 138 | 96 | 11.0 | 13.8% | 67.8 |
| G79 | 139 | 94 | 3.2 | 66.9% | 48.6 |
| G80 | 11,448 | 6,940 | 0.5 | 92.8% | 36.1 |
| G82 | 61 | 16 | 4.0 | 35.8% | 46.8 |
| H01 | 827 | 798 | 2.8 | 16.8% | 43.2 |
| H02 | 1,386 | 1,263 | 2.7 | 32.8% | 71.4 |
| H03 | 163 | 104 | 5.0 | 28.9% | 101.4 |
| H04 | 563 | 383 | 10.6 | 0.0% | 74.0 |
| H05 | 484 | 358 | 11.7 | 0.8% | 57.5 |
| H06 | 1,406 | 1,344 | 10.2 | 0.1% | 24.2 |
| H07 | 8,399 | 7,668 | 8.4 | 0.1% | 28.8 |
| H08 | 175 | 143 | 12.1 | 0.0% | 35.2 |
| H09 | 1,135 | 938 | 9.4 | 0.1% | 29.1 |
| H10 | 1,937 | 1,382 | 9.2 | 0.2% | 49.6 |
| H11 | 552 | 351 | 12.4 | 1.1% | 54.3 |
| H12 | 99 | 74 | 3.3 | 37.3% | 48.7 |
| H13 | 36 | 28 | 13.8 | 0.0% | 27.6 |
| H14 | 151 | 116 | 5.1 | 45.7% | 79.9 |
| H15 | 4,394 | 2,783 | 8.7 | 2.7% | 84.2 |
| H16 | 17 | 15 | 8.8 | 17.9% | 75.9 |
| H17 | 46 | 16 | 5.6 | 28.3% | 39.1 |
| H18 | 14 | 4 | 2.1 | 79.6% | 47.8 |
| H19 | 152 | 82 | 9.3 | 51.2% | 45.8 |
| H20 | 196,045 | 150,592 | 0.1 | 97.7% | 39.0 |
| H21 | 1,062 | 785 | 1.7 | 76.8% | 37.7 |
| H22 | 419,132 | 280,442 | 0.1 | 97.6% | 35.8 |
| H23 | 58,210 | 25,684 | 0.1 | 97.9% | 25.7 |
| H24 | 1,863 | 1,512 | 3.3 | 65.7% | 30.8 |
| H25 | 310,417 | 132,825 | 0.4 | 94.5% | 25.6 |
| H26 | 685 | 279 | 0.6 | 93.7% | 31.1 |
| H27 | 86 | 62 | 4.1 | 69.7% | 36.7 |
| H28 | 702 | 346 | 1.6 | 86.7% | 54.3 |
| H29 | 606 | 553 | 12.4 | 2.1% | 54.7 |
| H30 | 283 | 87 | 4.6 | 59.1% | 67.9 |
| H31 | 104 | 57 | 5.4 | 39.5% | 17.8 |
| H32 | 38 | 16 | 15.5 | 0.0% | 106.0 |
| H33 | 11,502 | 9,781 | 9.5 | 0.2% | 37.3 |
| H34 | 548 | 299 | 1.7 | 29.2% | 110.3 |
| H35 | 899 | 342 | 2.5 | 16.7% | 153.4 |
| H36 | 485 | 190 | 3.7 | 5.8% | 106.6 |
| H37 | 15 | 6 | 0.0 | 100.0% | 33.3 |
| H40 | 190 | 129 | 1.5 | 60.2% | 51.8 |
| H41 | 4,194 | 2,684 | 1.9 | 53.7% | 56.0 |
| H42 | 1,166 | 524 | 1.7 | 52.3% | 105.4 |
| H44 | 3,600 | 1,989 | 2.0 | 82.1% | 66.4 |
| H46 | 2,868 | 1,403 | 0.9 | 93.0% | 62.6 |
| H47 | 16 | 8 | 4.7 | 50.0% | 85.3 |
| H48 | 8,718 | 3,732 | 0.1 | 94.0% | 75.1 |
| H49 | 557 | 376 | 0.3 | 90.8% | 77.9 |
| H50 | 288 | 239 | 3.9 | 12.8% | 96.3 |
| H51 | 8,256 | 3,243 | 0.3 | 86.7% | 91.1 |
| H52 | 20,018 | 8,963 | 0.1 | 97.5% | 55.6 |
| H53 | 361 | 134 | 0.8 | 83.2% | 91.0 |
| H54 | 800 | 464 | 0.9 | 85.6% | 59.2 |
| H55 | 13,277 | 7,603 | 0.4 | 90.6% | 79.4 |
| H56 | 10,140 | 4,948 | 0.2 | 94.2% | 69.1 |
| H57 | 113 | 32 | 0.5 | 89.4% | 130.6 |
| H58 | 2,508 | 1,850 | 1.5 | 72.6% | 31.3 |
| H59 | 5,556 | 2,171 | 0.4 | 85.1% | 86.5 |
| H60 | 2,820 | 1,807 | 0.6 | 85.3% | 57.0 |
| H62 | 1,202 | 618 | 1.9 | 79.5% | 48.8 |
| H66 | 354 | 162 | 2.9 | 72.6% | 105.2 |
| H68 | 323 | 205 | 0.6 | 96.4% | 64.5 |
| H69 | 1,274 | 684 | 0.3 | 96.5% | 62.8 |
| H70 | 48 | 19 | 1.2 | 79.7% | 59.5 |
| H71 | 0 | 48 |  | 0.0% | 0.0 |
| J01 | 385 | 227 | 13.2 | 0.3% | 95.7 |
| J02 | 2,533 | 2,221 | 6.9 | 0.8% | 32.3 |
| J03 | 425 | 288 | 5.7 | 2.8% | 38.9 |
| J04 | 11 | 4 | 5.8 | 27.0% | 52.8 |
| J05 | 86 | 67 | 6.6 | 10.5% | 28.4 |
| J06 | 20 | 19 | 5.7 | 5.0% | 19.6 |
| J07 | 9 | 15 | 11.6 | 0.0% | 34.2 |
| J08 | 79 | 50 | 3.3 | 8.9% | 43.6 |
| J09 | 279 | 210 | 1.9 | 58.2% | 27.3 |
| J10 | 1,887 | 1,659 | 2.3 | 4.6% | 21.1 |
| J11 | 469 | 378 | 4.6 | 26.0% | 24.6 |
| J12 | 1,507 | 1,547 | 7.0 | 11.5% | 23.0 |
| J13 | 13,173 | 10,155 | 2.2 | 70.8% | 20.8 |
| J14 | 1,022 | 700 | 3.0 | 58.6% | 20.5 |
| J15 | 12 | 23 | 7.5 | 0.0% | 39.5 |
| J16 | 11 | 2 | 4.5 | 53.8% | 82.8 |
| J17 | 284 | 253 | 1.0 | 81.4% | 18.8 |
| J18 | 65,581 | 37,868 | 1.5 | 57.6% | 90.6 |
| J19 | 8 | 5 | 4.1 | 51.9% | 111.2 |
| J20 | 22 | 23 | 4.5 | 18.3% | 64.5 |
| J21 | 161 | 166 | 9.8 | 15.6% | 146.5 |
| J23 | 34 | 24 | 3.0 | 56.4% | 64.8 |
| J24 | 77 | 128 | 10.8 | 24.7% | 19.8 |
| J25 | 35 | 41 | 2.3 | 69.4% | 16.1 |
| J26 | 21 | 21 | 4.3 | 51.9% | 70.0 |
| J27 | 80 | 40 | 7.6 | 1.3% | 37.8 |
| J28 | 29 | 16 | 5.1 | 13.9% | 41.4 |
| J29 | 187 | 135 | 12.0 | 0.0% | 43.6 |
| J30 | 53 | 19 | 7.8 | 11.3% | 47.4 |
| J31 | 32 | 42 | 4.1 | 59.8% | 41.1 |
| J32 | 17 | 3 | 7.6 | 17.5% | 87.0 |
| J33 | 84 | 68 | 8.5 | 8.3% | 48.4 |
| J34 | 157 | 46 | 2.0 | 60.0% | 44.4 |
| J35 | 60 | 53 | 3.1 | 40.1% | 27.5 |
| J36 | 117 | 121 | 1.2 | 74.6% | 35.1 |
| J37 | 85 | 33 | 3.7 | 61.4% | 43.3 |
| J38 | 18,379 | 14,319 | 3.6 | 51.2% | 24.5 |
| J39 | 298 | 1,140 | 2.5 | 52.3% | 43.5 |
| J40 | 6,007 | 4,263 | 3.6 | 50.0% | 28.0 |
| J41 | 4,027 | 2,774 | 1.9 | 60.5% | 42.9 |
| J42 | 847 | 647 | 2.8 | 46.5% | 26.4 |
| J43 | 2,385 | 1,674 | 3.3 | 52.5% | 25.2 |
| J44 | 498 | 308 | 2.5 | 55.1% | 31.4 |
| J45 | 131 | 123 | 2.9 | 61.2% | 21.8 |
| J46 | 62 | 58 | 6.0 | 6.4% | 16.5 |
| J47 | 731 | 578 | 10.0 | 6.3% | 15.2 |
| J48 | 903 | 735 | 9.5 | 13.2% | 16.5 |
| J49 | 10 | 7 | 6.3 | 20.7% | 125.3 |
| J50 | 447 | 416 | 6.8 | 13.4% | 13.7 |
| J51 | 13 | 9 | 0.5 | 61.5% | 106.0 |
| J52 | 15 | 6 | 6.7 | 48.0% | 28.4 |
| J53 | 2,759 | 1,647 | 0.4 | 91.3% | 30.8 |
| J54 | 10 | 0 | 7.2 | 0.0% | 0.0 |
| J55 | 119 | 81 | 15.8 | 0.0% | 33.5 |
| J56 | 1,199 | 1,077 | 12.7 | 0.1% | 25.9 |
| J57 | 611 | 572 | 15.8 | 2.1% | 31.6 |
| J58 | 38 | 12 | 7.3 | 15.9% | 34.2 |
| J59 | 29 | 7 | 7.9 | 3.5% | 73.0 |
| J60 | 94 | 87 | 2.4 | 77.6% | 37.9 |
| J61 | 414 | 438 | 8.0 | 37.5% | 19.1 |
| J62 | 0 | 0 | - | 0.0% | 0.0 |
| J63 | 3 | 0 | 10.3 | 66.7% | 23.5 |
| J65 | 36 | 85 | 7.1 | 41.5% | 24.0 |
| J66 | 286 | 166 | 11.2 | 43.3% | 21.7 |
| J67 | 193 | 147 | 2.4 | 69.3% | 14.0 |
| J68 | 29 | 49 | 0.5 | 73.2% | 51.1 |
| J69 | 310 | 180 | 7.2 | 0.3% | 52.3 |
| J70 | 53 | 19 | 2.9 | 11.4% | 64.2 |
| J72 | 242 | 225 | 5.4 | 36.8% | 21.1 |
| J73 | 51 | 44 | 0.2 | 90.2% | 28.4 |
| J74 | 9,951 | 7,069 | 0.4 | 92.2% | 21.9 |
| J77 | 0 | 0 | 1.0 | 0.0% | 0.0 |
| K01 | 3 | 2 | 28.2 | 0.0% | 0.0 |
| K02 | 72 | 73 | 60.3 | 4.2% | 51.8 |
| K04 | 214 | 206 | 5.7 | 0.0% | 60.8 |
| K05 | 2 | 4 | 20.5 | 0.0% | 34.0 |
| K06 | 138 | 124 | 8.8 | 0.0% | 98.8 |
| K07 | 36 | 23 | 8.7 | 0.0% | 34.9 |
| K08 | 25 | 20 | 11.1 | 0.0% | 79.0 |
| K09 | 211 | 163 | 8.6 | 0.9% | 82.3 |
| K10 | 379 | 219 | 5.8 | 0.8% | 81.9 |
| K11 | 431 | 351 | 6.3 | 0.5% | 47.8 |
| K12 | 4 | 3 | 5.6 | 28.6% | 126.0 |
| K13 | 653 | 427 | 1.5 | 9.5% | 89.2 |
| K14 | 26 | 15 | 7.8 | 3.9% | 75.3 |
| K15 | 3 | 2 | 5.8 | 40.0% | 24.0 |
| K16 | 426 | 909 | 2.4 | 34.7% | 55.3 |
| K17 | 200 | 158 | 14.4 | 0.0% | 93.5 |
| K18 | 122 | 118 | 6.4 | 2.5% | 93.0 |
| K19 | 79 | 29 | 9.3 | 1.3% | 77.7 |
| K20 | 72 | 39 | 4.2 | 4.2% | 72.5 |
| K22 | 160 | 124 | 7.6 | 8.1% | 44.2 |
| K23 | 53 | 199 | 9.0 | 28.3% | 30.5 |
| K24 | 210 | 167 | 7.7 | 2.4% | 88.2 |
| K25 | 3,243 | 2,070 | 11.8 | 0.5% | 86.0 |
| K26 | 8,936 | 7,677 | 7.8 | 0.1% | 61.2 |
| K27 | 161 | 86 | 12.6 | 0.6% | 97.4 |
| K28 | 176 | 110 | 5.7 | 1.7% | 104.8 |
| K29 | 14 | 14 | 15.0 | 14.4% | 41.4 |
| K30 | 373 | 278 | 12.1 | 0.8% | 100.6 |
| K31 | 23 | 23 | 5.1 | 4.3% | 79.4 |
| K32 | 0 | 3 | - | 0.0% | 0.0 |
| K33 | 397 | 302 | 10.4 | 0.0% | 105.6 |
| K34 | 93 | 55 | 11.5 | 1.1% | 80.2 |
| K35 | 894 | 561 | 5.3 | 13.9% | 62.6 |
| K36 | 1 | 2 | 4.5 | 0.0% | 95.0 |
| K37 | 127 | 91 | 4.1 | 0.0% | 84.2 |
| K38 | 6 | 9 | 4.1 | 0.0% | 77.3 |
| K40 | 1,695 | 940 | 11.7 | 0.2% | 76.9 |
| K41 | 15 | 16 | 7.3 | 0.0% | 56.6 |
| K42 | 3 | 1 | 2.4 | 80.0% | 176.0 |
| K43 | 0 | 3 | - | 0.0% | 0.0 |
| K44 | 28 | 32 | 7.6 | 10.5% | 54.6 |
| K45 | 13,829 | 8,141 | 9.2 | 0.0% | 67.5 |
| K46 | 1 | 3 | 9.0 | 0.0% | 0.0 |
| K47 | 15 | 12 | 8.7 | 13.2% | 85.0 |
| K48 | 34 | 24 | 8.7 | 5.9% | 67.1 |
| K49 | 2,606 | 2,109 | 2.5 | 44.7% | 61.3 |
| K50 | 119 | 77 | 3.0 | 30.1% | 55.1 |
| K51 | 543 | 367 | 0.9 | 77.0% | 54.1 |
| K52 | 120 | 52 | 3.9 | 9.2% | 60.7 |
| K53 | 9 | 4 | 4.7 | 31.8% | 109.8 |
| K54 | 99 | 59 | 45.8 | 0.0% | 4.8 |
| K55 | 29 | 36 | 8.4 | 6.8% | 35.5 |
| K56 | 146 | 92 | 5.5 | 10.3% | 46.3 |
| K57 | 11,290 | 7,204 | 0.9 | 51.7% | 101.7 |
| K58 | 2,638 | 1,686 | 0.9 | 75.4% | 95.6 |
| K59 | 10,578 | 8,645 | 2.5 | 52.8% | 60.9 |
| K60 | 33,064 | 28,817 | 2.4 | 51.8% | 49.3 |
| K61 | 1,739 | 1,666 | 3.7 | 40.5% | 45.2 |
| K62 | 10,003 | 7,507 | 0.8 | 42.6% | 119.7 |
| K63 | 113,544 | 69,061 | 1.5 | 71.8% | 46.0 |
| K64 | 29 | 19 | 1.3 | 55.4% | 97.0 |
| K65 | 2,735 | 1,862 | 2.6 | 52.5% | 45.0 |
| K66 | 5 | 7 | 2.3 | 80.0% | 47.5 |
| K67 | 110 | 73 | 9.9 | 0.9% | 54.5 |
| K68 | 270 | 213 | 5.9 | 5.5% | 27.4 |
| K69 | 113 | 73 | 8.2 | 0.9% | 31.2 |
| K71 | 29 | 23 | 5.1 | 14.0% | 17.5 |
| K73 | 2,600 | 2,437 | 0.5 | 83.5% | 57.4 |
| K74 | 352 | 537 | 0.7 | 77.9% | 43.2 |
| K75 | 36,418 | 27,012 | 2.2 | 48.9% | 52.8 |
| K76 | 87 | 74 | 6.0 | 12.7% | 74.2 |
| K77 | 126 | 114 | 5.7 | 12.7% | 43.5 |
| K78 | 8 | 3 | 1.6 | 12.7% | 71.0 |
| L01 | 32 | 26 | 12.6 | 9.4% | 33.6 |
| L02 | 175 | 117 | 10.5 | 16.0% | 38.2 |
| L03 | 504 | 390 | 2.1 | 25.0% | 77.0 |
| L04 | 191 | 111 | 14.3 | 0.0% | 1.0 |
| L05 | 10 | 7 | 18.2 | 20.0% | 44.8 |
| L06 | 20 | 8 | 8.9 | 0.0% | 69.1 |
| L07 | 32 | 16 | 10.6 | 0.0% | 69.3 |
| L08 | 16 | 15 | 13.1 | 6.3% | 44.0 |
| L09 | 141 | 125 | 9.1 | 0.0% | 45.9 |
| L10 | 43 | 29 | 6.4 | 0.0% | 53.5 |
| L12 | 104 | 69 | 10.6 | 0.0% | 45.2 |
| L13 | 583 | 470 | 3.0 | 18.0% | 84.4 |
| L16 | 106 | 80 | 12.8 | 0.0% | 44.3 |
| L18 | 80 | 82 | 10.9 | 2.5% | 45.0 |
| L19 | 2,357 | 1,545 | 10.2 | 0.1% | 58.9 |
| L20 | 42 | 55 | 14.0 | 0.0% | 22.7 |
| L21 | 198 | 111 | 11.7 | 0.5% | 63.4 |
| L22 | 46 | 36 | 15.1 | 4.4% | 45.1 |
| L23 | 393 | 295 | 7.1 | 0.0% | 53.1 |
| L25 | 84 | 45 | 6.2 | 2.4% | 54.0 |
| L26 | 793 | 496 | 2.7 | 45.5% | 65.1 |
| L27 | 3,041 | 1,765 | 4.6 | 0.5% | 51.2 |
| L28 | 84 | 62 | 5.0 | 6.0% | 41.6 |
| L29 | 3,136 | 2,129 | 3.1 | 0.2% | 14.8 |
| L30 | 103 | 52 | 4.4 | 10.7% | 58.2 |
| L31 | 456 | 415 | 3.2 | 41.9% | 37.3 |
| L33 | 348 | 268 | 13.1 | 1.4% | 72.6 |
| L34 | 16 | 15 | 4.1 | 6.4% | 55.8 |
| L35 | 3,405 | 2,410 | 2.3 | 65.9% | 39.8 |
| L37 | 57 | 38 | 6.1 | 0.0% | 47.5 |
| L38 | 91 | 75 | 4.9 | 25.3% | 36.1 |
| L39 | 318 | 252 | 3.2 | 46.2% | 45.0 |
| L41 | 4 | 3 | 8.0 | 0.0% | 54.0 |
| L42 | 8 | 4 | 4.6 | 12.4% | 47.3 |
| L43 | 824 | 563 | 4.1 | 31.3% | 37.9 |
| L45 | 16 | 26 | 8.6 | 0.0% | 54.9 |
| L46 | 14 | 9 | 7.9 | 22.0% | 41.6 |
| L47 | 458 | 338 | 5.5 | 24.5% | 35.4 |
| L48 | 1 | 3 | 9.4 | 0.0% | 0.0 |
| L49 | 56 | 26 | 8.3 | 3.6% | 39.7 |
| L50 | 8 | 6 | 14.9 | 0.0% | 55.6 |
| L51 | 456 | 221 | 8.6 | 0.0% | 54.9 |
| L52 | 90 | 36 | 5.7 | 0.0% | 36.9 |
| L53 | 48 | 39 | 8.0 | 8.3% | 71.6 |
| L54 | 4,130 | 2,717 | 2.3 | 58.3% | 55.1 |
| L56 | 6 | 1 | 13.2 | 0.0% | 24.0 |
| L57 | 246 | 154 | 6.6 | 2.8% | 49.4 |
| L58 | 49 | 62 | 15.1 | 2.0% | 28.5 |
| L59 | 2,180 | 1,405 | 9.8 | 0.8% | 39.1 |
| L60 | 2,147 | 1,297 | 6.5 | 0.4% | 52.5 |
| L62 | 282 | 181 | 8.4 | 4.6% | 41.1 |
| L63 | 10,703 | 7,213 | 3.6 | 53.4% | 41.2 |
| L65 | 22 | 10 | 11.9 | 8.9% | 31.4 |
| L66 | 1,427 | 781 | 4.3 | 49.9% | 39.2 |
| L67 | 4,019 | 2,157 | 0.7 | 93.0% | 9.6 |
| L68 | 139 | 118 | 3.1 | 52.4% | 20.6 |
| L69 | 75 | 78 | 5.5 | 16.0% | 60.9 |
| L70 | 3,700 | 1,391 | 1.2 | 81.8% | 98.6 |
| L71 | 3,877 | 2,230 | 4.1 | 21.9% | 57.8 |
| L72 | 470 | 304 | 4.0 | 48.6% | 40.9 |
| L73 | 8 | 1 | 1.0 | 37.5% | 55.3 |
| L74 | 14,277 | 11,067 | 1.3 | 72.4% | 38.6 |
| L75 | 1,691 | 827 | 2.4 | 64.5% | 77.8 |
| L76 | 4 | 0 | 7.0 | 0.0% | 68.0 |
| L77 | 4 | 13 | 20.5 | 0.0% | 129.0 |
| L79 | 1,801 | 1,198 | 4.8 | 53.5% | 40.1 |
| L80 | 31 | 27 | 5.8 | 16.4% | 75.9 |
| L81 | 16 | 6 | 7.8 | 12.4% | 82.3 |
| L82 | 9 | 8 | 2.6 | 33.8% | 83.3 |
| L83 | 48 | 10 | 2.1 | 83.3% | 98.9 |
| L84 | 3,077 | 586 | 0.1 | 90.3% | 97.7 |
| L85 | 254 | 90 | 0.3 | 86.3% | 100.2 |
| L86 | 6,123 | 1,886 | 0.1 | 99.1% | 94.3 |
| L87 | 4,471 | 1,264 | 0.1 | 95.7% | 91.4 |
| L88 | 14,886 | 4,600 | 0.1 | 97.9% | 100.0 |
| L90 | 25 | 17 | 6.9 | 32.0% | 33.1 |
| L93 | 379 | 209 | 2.5 | 71.8% | 57.2 |
| L94 | 10,297 | 10,576 | 1.6 | 81.0% | 26.4 |
| L95 | 1,549 | 766 | 1.5 | 79.0% | 53.2 |
| L96 | 49 | 38 | 5.2 | 22.3% | 8.3 |
| L97 | 471 | 209 | 4.7 | 46.1% | 70.2 |
| L98 | 21 | 14 | 2.0 | 60.5% | 100.8 |
| L99 | 24,285 | 21,446 | 7.6 | 50.5% | 14.4 |
| M01 | 1,064 | 516 | 8.6 | 0.0% | 36.9 |
| M02 | 7,013 | 5,235 | 4.4 | 0.3% | 40.8 |
| M03 | 1,803 | 1,527 | 3.4 | 0.4% | 42.1 |
| M04 | 157 | 81 | 1.9 | 21.7% | 96.9 |
| M05 | 625 | 467 | 2.7 | 2.1% | 71.1 |
| M06 | 358 | 333 | 6.9 | 20.4% | 42.7 |
| M07 | 0 | 3,265 | - | 0.0% | 0.0 |
| M08 | 57 | 43 | 4.6 | 35.0% | 48.0 |
| M09 | 8,956 | 4,238 | 1.0 | 52.1% | 81.6 |
| M10 | 1,045 | 606 | 1.9 | 19.5% | 83.2 |
| M11 | 913 | 640 | 0.8 | 64.6% | 63.6 |
| M12 | 6 | 0 | 4.0 | 83.3% | 20.5 |
| M13 | 13,869 | 11,130 | 4.3 | 49.6% | 21.4 |
| M14 | 16,278 | 9,222 | 0.0 | 98.8% | 46.7 |
| M15 | 387 | 271 | 3.7 | 49.7% | 22.1 |
| M16 | 6,758 | 6,210 | 2.5 | 54.4% | 63.0 |
| M17 | 108 | 28 | 0.2 | 98.7% | 23.2 |
| M18 | 139 | 99 | 7.4 | 2.9% | 45.8 |
| M19 | 480 | 299 | 7.8 | 21.0% | 98.1 |
| M20 | 231 | 191 | 5.4 | 2.2% | 94.0 |
| M21 | 74 | 53 | 6.6 | 2.7% | 74.7 |
| M22 | 6 | 5 | 0.9 | 31.3% | 83.3 |
| M23 | 23 | 15 | 1.8 | 39.1% | 49.9 |
| M24 | 0 | 22 | - | 0.0% | 0.0 |
| M25 | 244 | 230 | 2.6 | 30.0% | 95.9 |
| M26 | 317 | 271 | 2.7 | 41.6% | 51.9 |
| M27 | 16,199 | 13,763 | 1.2 | 58.5% | 62.8 |
| M28 | 1,222 | 11 | 1.3 | 54.4% | 58.7 |
| M29 | 23,155 | 16,862 | 1.7 | 68.2% | 53.5 |
| M30 | 1,479 | 1,064 | 0.8 | 67.0% | 62.5 |
| M31 | 2,851 | 3,970 | 0.2 | 95.6% | 20.7 |
| M32 | 684 | 522 | 0.5 | 75.8% | 80.7 |
| M33 | 1,769 | 1,840 | 4.8 | 40.3% | 33.8 |
| M34 | 1,871 | 1,535 | 9.3 | 1.4% | 39.3 |
| M35 | 142 | 118 | 5.5 | 4.2% | 72.2 |
| M36 | 78 | 33 | 9.2 | 0.0% | 123.0 |
| M37 | 115 | 72 | 8.6 | 3.5% | 88.5 |
| M38 | 3,133 | 1,643 | 3.1 | 44.9% | 88.2 |
| M39 | 184 | 86 | 5.6 | 2.7% | 106.1 |
| M41 | 62 | 54 | 2.9 | 25.9% | 102.1 |
| M42 | 36,776 | 30,973 | 1.5 | 37.3% | 36.3 |
| M43 | 11,846 | 5,331 | 0.2 | 91.6% | 111.0 |
| M44 | 5,229 | 3,139 | 1.9 | 36.6% | 94.8 |
| M45 | 94,771 | 59,570 | 0.4 | 91.8% | 45.5 |
| M48 | 296 | 165 | 2.0 | 71.4% | 71.2 |
| M49 | 58,213 | 31,974 | 0.4 | 96.6% | 53.6 |
| M51 | 81 | 21 | 4.5 | 4.9% | 129.0 |
| M52 | 854 | 490 | 2.5 | 2.5% | 121.7 |
| M53 | 1,188 | 191 | 1.0 | 51.8% | 111.0 |
| M54 | 30 | 10 | 6.0 | 23.4% | 108.8 |
| M55 | 47 | 13 | 3.3 | 21.3% | 107.5 |
| M56 | 3,006 | 1,392 | 0.1 | 90.9% | 105.4 |
| M57 | 70 | 32 | 1.2 | 51.4% | 95.9 |
| M58 | 144 | 51 | 0.1 | 89.6% | 82.1 |
| M60 | 70 | 30 | 3.2 | 12.9% | 88.9 |
| M61 | 9,778 | 6,226 | 1.7 | 2.0% | 41.4 |
| M62 | 34 | 24 | 3.6 | 53.2% | 62.3 |
| M64 | 541 | 161 | 1.8 | 12.0% | 113.8 |
| M65 | 21,043 | 11,074 | 2.0 | 8.5% | 106.1 |
| M66 | 1,396 | 699 | 1.1 | 30.0% | 101.9 |
| M67 | 531 | 340 | 1.6 | 59.0% | 75.8 |
| M68 | 1,317 | 546 | 0.2 | 84.7% | 93.3 |
| M70 | 42,833 | 27,558 | 0.1 | 93.5% | 31.0 |
| M71 | 589 | 425 | 0.9 | 46.5% | 38.3 |
| M72 | 475 | 253 | 1.3 | 30.1% | 104.8 |
| M73 | 3,086 | 1,181 | 0.7 | 56.4% | 146.3 |
| M75 | 90 | 46 | 1.2 | 53.2% | 72.9 |
| M76 | 12,132 | 6,765 | 0.5 | 77.2% | 76.2 |
| M77 | 1,620 | 1,038 | 0.4 | 92.6% | 40.8 |
| M79 | 6,764 | 2,773 | 0.3 | 86.9% | 69.9 |
| M81 | 2,950 | 1,603 | 0.4 | 87.2% | 72.8 |
| M83 | 95 | 82 | 0.4 | 89.3% | 19.2 |
| M85 | 185 | 80 | 1.4 | 73.1% | 94.7 |
| M86 | 154 | 83 | 1.2 | 65.8% | 71.4 |
| N01 | 1,378 | 624 | 1.0 | 91.1% | 82.2 |
| N03 | 567 | 302 | 2.1 | 69.6% | 75.1 |
| N05 | 93 | 46 | 0.8 | 63.2% | 54.6 |
| N06 | 3,629 | 2,774 | 0.8 | 76.9% | 38.0 |
| N07 | 245 | 142 | 0.6 | 87.3% | 92.7 |
| N08 | 943 | 585 | 0.4 | 85.2% | 112.7 |
| N09 | 5,500 | 2,654 | 0.2 | 92.7% | 107.0 |
| N10 | 241 | 71 | 0.2 | 86.4% | 117.0 |
| N11 | 5,844 | 2,764 | 0.2 | 88.2% | 98.4 |
| N13 | 521 | 325 | 0.7 | 84.1% | 77.6 |
| N15 | 2,823 | 1,142 | 0.2 | 93.5% | 99.8 |
| N17 | 8,775 | 2,499 | 0.0 | 99.3% | 87.6 |
| N18 | 26 | 17 | 0.5 | 84.6% | 95.0 |
| N19 | 1,674 | 918 | 0.1 | 96.4% | 70.8 |
| N20 | 235 | 82 | 0.3 | 89.5% | 78.5 |
| N22 | 43 | 9 | 0.6 | 74.4% | 67.0 |
| N24 | 432 | 183 | 3.8 | 64.4% | 71.3 |
| N26 | 465 | 377 | 4.4 | 10.3% | 21.1 |
| N27 | 1,453 | 734 | 0.5 | 86.9% | 63.0 |
| N28 | 3,277 | 1,078 | 0.5 | 81.5% | 107.9 |
| N29 | 501 | 148 | 2.1 | 4.4% | 100.2 |
| N30 | 28,358 | 12,766 | 0.2 | 94.6% | 90.6 |
| N32 | 1,944 | 1,095 | 0.6 | 90.8% | 55.0 |
| N34 | 569 | 220 | 0.0 | 96.7% | 85.3 |
| O01 | 838 | 623 | 9.9 | 4.5% | 48.2 |
| O02 | 294 | 168 | 10.5 | 2.4% | 61.6 |
| O03 | 368 | 303 | 6.2 | 0.8% | 56.5 |
| O04 | 348 | 320 | 6.4 | 8.0% | 55.3 |
| O05 | 245 | 199 | 7.5 | 1.2% | 40.0 |
| O06 | 143 | 33 | 4.1 | 2.1% | 118.9 |
| O07 | 832 | 309 | 2.1 | 0.5% | 127.6 |
| O08 | 48 | 11 | 2.9 | 2.1% | 102.0 |
| O09 | 26 | 31 | 2.6 | 53.8% | 111.8 |
| O10 | 59 | 15 | 0.7 | 67.8% | 98.7 |
| O15 | 1,011 | 702 | 14.6 | 6.5% | 0.0 |
| O17 | 1,381 | 962 | 2.3 | 71.6% | 10.0 |
| O18 | 173 | 71 | 6.2 | 1.2% | 117.0 |
| O19 | 465 | 167 | 0.2 | 89.0% | 85.7 |
| O20 | 1 | 1 | 0.0 | 100.0% | 113.0 |
| O21 | 480 | 200 | 4.4 | 1.5% | 123.6 |
| O22 | 22 | 8 | 2.4 | 0.0% | 89.9 |
| O23 | 80 | 57 | 6.4 | 10.0% | 87.1 |
| O24 | 46 | 20 | 2.6 | 28.5% | 49.3 |
| O25 | 229 | 217 | 2.0 | 37.9% | 64.5 |
| O26 | 134 | 131 | 2.1 | 44.1% | 51.0 |
| O27 | 4,560 | 2,117 | 0.5 | 69.9% | 99.8 |
| O29 | 19,760 | 5,750 | 0.2 | 81.6% | 95.9 |
| O32 | 961 | 340 | 2.9 | 2.9% | 128.8 |
| O35 | 0 | 30 | - | 0.0% | 0.0 |
| O37 | 0 | 36 | - | 0.0% | 0.0 |
| O38 | 0 | 84 | - | 0.0% | 0.0 |
| O39 | 0 | 54 | - | 0.0% | 0.0 |
| O40 | 0 | 111 | - | 0.0% | 0.0 |
| P01 | 93 | 49 | 0.5 | 74.4% | 61.8 |
| P03 | 1,866 | 878 | 0.5 | 87.3% | 59.5 |
| P05 | 5,064 | 2,950 | 1.1 | 74.4% | 47.4 |
| P06 | 1,561 | 800 | 0.2 | 92.3% | 56.1 |
| P07 | 241 | 98 | 0.3 | 88.5% | 83.2 |
| P09 | 4,206 | 1,812 | 0.6 | 90.7% | 42.6 |
| P11 | 463 | 168 | 1.2 | 84.3% | 73.8 |
| P13 | 1,896 | 644 | 1.0 | 82.0% | 83.9 |
| P14 | 28 | 15 | 0.4 | 82.1% | 90.5 |
| P15 | 461 | 228 | 0.2 | 94.1% | 74.9 |
| P17 | 129 | 77 | 1.7 | 58.1% | 68.0 |
| P18 | 383 | 156 | 1.9 | 11.0% | 108.2 |
| P19 | 208 | 95 | 0.3 | 76.5% | 71.3 |
| P20 | 2,414 | 1,433 | 0.4 | 85.6% | 64.1 |
| P21 | 61 | 29 | 3.4 | 47.7% | 89.3 |
| P22 | 257 | 76 | 1.5 | 7.8% | 131.6 |
| P23 | 18,069 | 6,023 | 1.5 | 8.8% | 115.8 |
| P24 | 1,216 | 337 | 1.8 | 4.7% | 130.5 |
| P25 | 415 | 135 | 2.0 | 24.3% | 110.7 |
| P26 | 669 | 492 | 2.0 | 73.2% | 60.8 |
| P27 | 518 | 279 | 0.7 | 88.5% | 46.3 |
| P28 | 69 | 27 | 0.9 | 52.2% | 91.0 |
| P29 | 1,547 | 780 | 0.7 | 79.3% | 47.9 |
| P30 | 29 | 36 | 3.0 | 37.9% | 80.6 |
| P31 | 1,234 | 685 | 0.5 | 78.3% | 93.8 |
| P32 | 26 | 9 | 1.1 | 50.0% | 118.4 |
| Q01 | 10,670 | 6,692 | 0.1 | 94.8% | 37.5 |
| Q02 | 2,232 | 650 | 0.1 | 96.5% | 62.0 |
| Q03 | 2,764 | 1,221 | 0.5 | 93.6% | 31.7 |
| Q05 | 873 | 531 | 0.5 | 83.9% | 53.5 |
| Q07 | 28,748 | 19,517 | 2.8 | 2.0% | 75.3 |
| Q08 | 5,305 | 2,889 | 1.7 | 3.0% | 91.1 |
| Q09 | 2,965 | 1,464 | 2.5 | 9.6% | 110.5 |
| Q10 | 1,884 | 1,141 | 0.3 | 90.9% | 34.1 |
| Q16 | 8,981 | 3,698 | 0.1 | 92.9% | 75.5 |
| Q17 | 30,458 | 17,991 | 0.2 | 90.8% | 50.9 |
| Q18 | 39,233 | 21,084 | 0.1 | 94.1% | 44.9 |
| Q19 | 3 | 0 | 0.0 | 100.0% | 112.7 |
| Q20 | 1,550 | 777 | 1.0 | 86.2% | 59.9 |
| Q22 | 8,106 | 5,482 | 1.3 | 45.1% | 76.1 |
| Q23 | 6,107 | 3,710 | 1.6 | 37.7% | 76.4 |
| Q24 | 251 | 188 | 1.4 | 52.9% | 75.1 |
| Q25 | 272 | 163 | 1.1 | 66.1% | 79.1 |
| Q26 | 2 | 2 | 0.0 | 100.0% | 60.5 |
| Q27 | 252 | 88 | 0.2 | 82.5% | 92.8 |
| Q28 | 73 | 27 | 0.3 | 84.9% | 69.4 |
| Q29 | 9 | 5 | 0.9 | 33.3% | 45.7 |
| Q30 | 115 | 40 | 0.8 | 78.5% | 98.2 |
| Q31 | 140 | 77 | 2.2 | 51.5% | 89.8 |
| Q32 | 191 | 86 | 0.6 | 75.9% | 80.4 |
| Q34 | 12 | 10 | 2.9 | 66.7% | 104.2 |
| Q35 | 6,006 | 1,757 | 0.1 | 88.0% | 88.0 |
| Q36 | 157 | 41 | 0.1 | 89.8% | 94.9 |
| Q37 | 37 | 15 | 0.1 | 83.8% | 69.3 |
| Q38 | 310 | 101 | 0.6 | 78.3% | 92.7 |
| Q39 | 82 | 18 | 0.4 | 87.8% | 40.3 |
| Q41 | 4,206 | 1,342 | 0.1 | 91.5% | 70.5 |
| Q43 | 1,561 | 962 | 4.1 | 36.3% | 80.5 |
| Q44 | 88 | 44 | 1.0 | 74.8% | 87.9 |
| Q45 | 21 | 11 | 1.4 | 67.4% | 56.5 |
| Q47 | 204 | 171 | 3.3 | 41.2% | 63.2 |
| Q49 | 7,940 | 4,342 | 0.9 | 62.2% | 90.2 |
| Q50 | 159 | 151 | 2.1 | 64.8% | 43.7 |
| Q51 | 151 | 94 | 3.1 | 67.4% | 49.5 |
| Q52 | 203 | 100 | 0.7 | 66.1% | 93.9 |
| Q54 | 1,553 | 795 | 0.6 | 59.4% | 100.6 |
| Q55 | 7,103 | 4,495 | 1.2 | 81.9% | 44.0 |
| Q56 | 15 | 16 | 0.5 | 46.7% | 94.4 |
| Q57 | 8 | 3 | 0.4 | 75.0% | 80.8 |
| S01 | 547 | 175 | 0.4 | 74.0% | 115.0 |
| S02 | 477 | 101 | 4.4 | 6.1% | 124.1 |
| S03 | 136 | 48 | 1.0 | 59.6% | 108.0 |
| S04 | 1,032 | 260 | 0.6 | 79.1% | 79.2 |
| S05 | 2,630 | 1,943 | 0.0 | 98.3% | 92.7 |
| S06 | 203,861 | 141,448 | 0.1 | 96.6% | 51.8 |
| S08 | 6,905 | 5,251 | 0.2 | 98.1% | 45.6 |
| S10 | 320 | 173 | 0.6 | 94.3% | 59.0 |
| S11 | 400 | 231 | 0.7 | 89.3% | 62.3 |
| S17 | 42 | 28 | 17.2 | 4.7% | 61.7 |
| S18 | 126 | 93 | 10.3 | 1.6% | 77.0 |
| S19 | 8 | 12 | 9.6 | 35.5% | 52.8 |
| S20 | 30 | 24 | 16.3 | 13.4% | 116.8 |
| S21 | 5 | 2 | 11.9 | 21.4% | 29.5 |
| S22 | 18 | 15 | 2.2 | 66.6% | 3.0 |
| S23 | 399 | 138 | 0.4 | 83.0% | 139.0 |
| S24 | 80 | 56 | 18.0 | 17.4% | 48.8 |
| S25 | 90 | 43 | 10.0 | 15.6% | 44.2 |
| S26 | 86 | 47 | 7.5 | 60.4% | 32.3 |
| S27 | 1,218 | 1,141 | 1.6 | 90.9% | 51.5 |
| S28 | 5 | 5 | 0.6 | 80.0% | 25.3 |
| S29 | 0 | 4 | - | 0.0% | 0.0 |
| S30 | 303 | 145 | 1.3 | 72.6% | 63.7 |
| S31 | 405 | 180 | 2.1 | 74.2% | 108.3 |
| S32 | 0 | 1 | - | 0.0% | 0.0 |
| S33 | 264 | 0 | 0.1 | 100.0% | 135.2 |
| S34 | 0 | 0 | - | 0.0% | 0.0 |
| S35 | 2,488 | 1,309 | 6.5 | 52.0% | 24.6 |
| S36 | 843 | 670 | 1.2 | 91.7% | 49.5 |
| S37 | 105 | 57 | 9.9 | 57.0% | 57.7 |
| S38 | 1 | 1 | 0.0 | 100.0% | 228.0 |
| S39 | 36 | 9 | 0.5 | 69.4% | 123.1 |
| S40 | 66 | 56 | 1.8 | 74.4% | 18.4 |
| S41 | 765 | 581 | 1.4 | 86.4% | 11.1 |
| S42 | 4,850 | 3,569 | 1.2 | 86.7% | 4.9 |
| S43 | 2,000 | 974 | 1.5 | 87.8% | 51.6 |
| S47 | 8,141 | 5,094 | 2.2 | 73.0% | 21.6 |
| S48 | 226 | 48 | 1.1 | 37.2% | 105.3 |
| S49 | 440 | 127 | 0.9 | 65.6% | 67.4 |
| S54 | 176 | 85 | 3.1 | 63.6% | 29.9 |
| S55 | 1,655 | 943 | 2.3 | 73.3% | 4.6 |
| S56 | 666 | 365 | 2.6 | 74.7% | 24.5 |
| S57 | 12,819 | 9,758 | 5.4 | 62.6% | 19.8 |
| S60 | 3,125 | 1,235 | 0.8 | 84.2% | 90.7 |
| S62 | 1,477 | 628 | 0.5 | 81.7% | 109.9 |
| S63 | 13 | 11 | 10.1 | 61.5% | 88.7 |
| S64 | 2,268 | 803 | 0.1 | 96.7% | 68.5 |
| S66 | 5,217 | 3,223 | 0.2 | 96.9% | 15.5 |
| S68 | 3,630 | 1,507 | 0.3 | 96.3% | 65.6 |
| S70 | 1,941 | 920 | 0.4 | 96.0% | 50.4 |
| T01 | 700 | 481 | 2.5 | 55.4% | 47.7 |
| T02 | 555 | 200 | 4.1 | 18.2% | 108.2 |
| T03 | 212 | 100 | 10.0 | 2.8% | 43.5 |
| T05 | 981 | 646 | 5.0 | 28.3% | 45.5 |
| T07 | 813 | 424 | 9.4 | 0.0% | 23.6 |
| T08 | 336 | 211 | 8.0 | 3.6% | 69.4 |
| T09 | 713 | 411 | 5.5 | 3.6% | 18.5 |
| T10 | 1,753 | 1,018 | 6.1 | 4.6% | 12.9 |
| T11 | 973 | 735 | 2.6 | 21.2% | 12.6 |
| T14 | 884 | 824 | 2.5 | 65.7% | 10.3 |
| T15 | 0 | 5 | 6.9 | 0.0% | 0.0 |
| T16 | 294 | 141 | 8.6 | 0.0% | 54.0 |
| T17 | 34 | 43 | 3.5 | 8.8% | 53.8 |
| T19 | 4,866 | 2,652 | 0.9 | 83.0% | 78.5 |
| T20 | 61,350 | 26,396 | 0.5 | 79.3% | 82.7 |
| T21 | 5,115 | 2,002 | 0.8 | 71.7% | 90.5 |
| T22 | 1,754 | 766 | 2.8 | 72.0% | 73.4 |
| T23 | 160 | 56 | 2.0 | 68.8% | 87.5 |
| T24 | 19,454 | 7,430 | 0.7 | 83.6% | 86.4 |
| T25 | 7,747 | 2,935 | 3.5 | 28.5% | 113.2 |
| T26 | 1,424 | 513 | 4.0 | 22.3% | 121.4 |
| T27 | 6,533 | 2,331 | 1.2 | 73.8% | 91.5 |
| T28 | 418 | 202 | 9.1 | 22.2% | 109.7 |
| T29 | 735 | 410 | 1.1 | 81.1% | 77.2 |
| T30 | 691 | 416 | 7.4 | 17.5% | 66.4 |
| T31 | 2,338 | 1,573 | 4.0 | 61.1% | 56.6 |
| T32 | 391 | 225 | 7.9 | 5.4% | 153.7 |
| T33 | 284 | 222 | 7.0 | 18.3% | 53.5 |
| T34 | 192 | 131 | 9.1 | 24.5% | 47.1 |
| T36 | 1,774 | 1,668 | 3.4 | 54.4% | 21.9 |
| T37 | 236 | 209 | 4.7 | 34.3% | 32.1 |
| T38 | 72 | 56 | 4.8 | 25.0% | 32.6 |
| T39 | 393 | 381 | 6.7 | 28.3% | 29.7 |
| T41 | 1,925 | 1,342 | 8.1 | 31.0% | 67.7 |
| T42 | 9,049 | 4,578 | 1.2 | 70.2% | 92.7 |
| T43 | 14,430 | 6,717 | 0.8 | 81.3% | 69.1 |
| T45 | 1,522 | 1,277 | 10.1 | 39.6% | 15.8 |
| T48 | 206 | 226 | 1.9 | 81.9% | 32.3 |
| T50 | 11 | 3 | 1.4 | 9.1% | 85.1 |
| T51 | 10 | 10 | 6.7 | 60.0% | 141.2 |
| T52 | 14,308 | 5,098 | 0.1 | 95.5% | 99.4 |
| T53 | 450 | 384 | 0.9 | 78.0% | 61.0 |
| T54 | 1,904 | 860 | 0.5 | 95.6% | 94.2 |
| T55 | 617 | 184 | 4.1 | 68.1% | 107.6 |
| T56 | 737 | 309 | 0.3 | 87.1% | 115.3 |
| T57 | 2,122 | 507 | 0.4 | 95.6% | 77.8 |
| T59 | 7,315 | 2,102 | 0.0 | 97.7% | 75.6 |
| T60 | 267 | 99 | 0.1 | 95.1% | 95.5 |
| T61 | 616 | 178 | 0.1 | 99.1% | 66.8 |
| T62 | 12,317 | 3,589 | 0.4 | 96.2% | 83.3 |
| T64 | 1,686 | 742 | 0.8 | 64.4% | 112.3 |
| T65 | 943 | 436 | 0.8 | 82.4% | 82.1 |
| T67 | 10,607 | 7,386 | 0.9 | 77.3% | 30.2 |
| T68 | 447 | 232 | 1.5 | 67.6% | 80.3 |
| T69 | 2,870 | 908 | 0.2 | 89.1% | 94.6 |
| T70 | 4,323 | 1,684 | 0.7 | 71.2% | 103.0 |
| T71 | 480 | 156 | 0.3 | 93.1% | 80.0 |
| T72 | 11,614 | 4,692 | 0.4 | 96.3% | 77.7 |
| T74 | 7,994 | 2,432 | 0.2 | 97.1% | 76.9 |
| T76 | 90 | 30 | 10.2 | 6.6% | 135.4 |
| T77 | 2,109 | 1,367 | 2.8 | 49.5% | 71.3 |
| T79 | 9,691 | 4,310 | 0.5 | 70.9% | 78.8 |
| T80 | 524 | 222 | 0.6 | 70.0% | 110.8 |
| T81 | 2,221 | 1,359 | 2.0 | 83.7% | 70.4 |
| T83 | 830 | 666 | 6.7 | 63.8% | 81.7 |
| T85 | 4,593 | 4,245 | 3.1 | 21.1% | 20.9 |
| T86 | 342 | 238 | 2.0 | 65.1% | 18.5 |
| T87 | 14,277 | 11,131 | 2.1 | 73.6% | 16.7 |
| T88 | 162 | 106 | 3.2 | 70.6% | 17.5 |
| T89 | 28 | 15 | 11.3 | 46.7% | 175.7 |
| T90 | 5 | 13 | 0.0 | 40.0% | 0.0 |
| T91 | 108 | 51 | 0.6 | 78.7% | 17.9 |
| T92 | 309 | 161 | 2.3 | 70.6% | 65.6 |
| T94 | 610 | 378 | 0.9 | 41.2% | 74.7 |
| T96 | 6,870 | 4,468 | 1.8 | 79.7% | 63.9 |
| T97 | 1,237 | 470 | 1.1 | 65.5% | 92.5 |
| T98 | 588 | 222 | 1.5 | 62.8% | 96.5 |
| V01 | 914 | 487 | 8.8 | 2.3% | 86.8 |
| V02 | 127 | 78 | 5.5 | 2.4% | 88.5 |
| V03 | 211 | 180 | 16.2 | 1.9% | 49.8 |
| V04 | 20 | 7 | 0.4 | 80.0% | 45.7 |
| V05 | 563 | 324 | 5.0 | 32.1% | 72.5 |
| V06 | 493 | 405 | 5.8 | 22.1% | 37.7 |
| V07 | 225 | 124 | 2.3 | 56.9% | 70.2 |
| V08 | 104 | 73 | 2.8 | 15.4% | 5.4 |
| V09 | 6,816 | 1,522 | 0.3 | 86.1% | 6.0 |
| V10 | 1,936 | 506 | 1.6 | 2.2% | 136.9 |
| V11 | 541 | 232 | 0.9 | 65.5% | 52.5 |
| V12 | 208 | 150 | 4.7 | 9.1% | 88.1 |
| V13 | 659 | 324 | 1.0 | 37.5% | 96.4 |
| V14 | 1,077 | 868 | 7.3 | 36.2% | 53.3 |
| V15 | 453 | 229 | 2.1 | 18.5% | 6.2 |
| V16 | 853 | 306 | 1.4 | 3.6% | 130.0 |
| V17 | 1,224 | 632 | 1.3 | 77.3% | 59.1 |
| V18 | 153 | 109 | 1.7 | 44.5% | 73.8 |
| V19 | 924 | 492 | 1.5 | 68.3% | 61.0 |
| V20 | 201 | 76 | 1.9 | 6.0% | 148.1 |
| V21 | 2,205 | 762 | 0.3 | 87.8% | 109.8 |
| V22 | 6,097 | 3,499 | 4.8 | 1.7% | 91.2 |
| V23 | 170 | 86 | 4.7 | 1.8% | 122.9 |
| V24 | 470 | 393 | 11.6 | 0.0% | 77.7 |
| V25 | 16,795 | 8,118 | 2.8 | 8.8% | 103.1 |
| V26 | 1,934 | 936 | 3.8 | 7.3% | 113.4 |
| V27 | 55 | 12 | 4.5 | 7.3% | 135.5 |
| V28 | 51 | 11 | 2.3 | 5.9% | 130.7 |
| V29 | 2,464 | 1,003 | 3.6 | 6.7% | 102.6 |
| V30 | 33 | 15 | 3.1 | 6.1% | 167.8 |
| V31 | 75 | 37 | 11.6 | 0.0% | 88.9 |
| V32 | 1 | 0 | 6.0 | 0.0% | 316.0 |
| V33 | 3,812 | 1,582 | 2.2 | 16.9% | 88.1 |
| V34 | 561 | 227 | 2.4 | 13.5% | 112.1 |
| V35 | 13 | 9 | 2.7 | 7.8% | 134.8 |
| V36 | 229 | 86 | 1.8 | 1.7% | 94.6 |
| V37 | 94 | 67 | 9.0 | 1.1% | 96.3 |
| V38 | 1,834 | 911 | 4.4 | 1.5% | 154.9 |
| V39 | 263 | 113 | 5.9 | 0.4% | 189.8 |
| V40 | 2,098 | 977 | 7.8 | 2.7% | 120.8 |
| V41 | 2,536 | 1,528 | 6.1 | 8.2% | 143.4 |
| V42 | 87 | 48 | 10.2 | 1.1% | 102.2 |
| V43 | 243 | 151 | 6.8 | 9.0% | 72.3 |
| V44 | 1,495 | 599 | 3.8 | 36.3% | 54.8 |
| V45 | 19 | 11 | 9.2 | 36.9% | 84.1 |
| V46 | 441 | 259 | 11.2 | 3.9% | 100.4 |
| V47 | 624 | 587 | 6.9 | 55.0% | 27.3 |
| V48 | 16,597 | 7,088 | 0.0 | 99.4% | 172.6 |
| V49 | 26 | 12 | 8.4 | 15.4% | 83.4 |
| V50 | 372 | 168 | 2.9 | 76.7% | 80.2 |
| V51 | 170 | 111 | 7.8 | 0.6% | 121.1 |
| V52 | 343 | 142 | 6.6 | 68.0% | 111.7 |
| V54 | 44,683 | 11,200 | 0.2 | 99.0% | 124.9 |
| V55 | 3 | 7 | 0.0 | 100.0% | 85.7 |
| V56 | 3 | 0 | 1.0 | 33.3% | 65.0 |
| V57 | 1 | 0 | 1.0 | 0.0% | 37.0 |
| V66 | 2 | 4 | 16.3 | 0.0% | 141.0 |
| V67 | 1,591 | 663 | 2.7 | 5.0% | 103.5 |
| V68 | 133 | 57 | 2.4 | 0.8% | 96.1 |
| V69 | 0 | 505 | - | 0.0% | 0.0 |
| V70 | 0 | 10 | - | 0.0% | 0.0 |
| W01 | 61 | 28 | 1.5 | 39.3% | 177.0 |
| W02 | 202 | 80 | 0.7 | 60.9% | 126.6 |
| W03 | 3,162 | 957 | 0.9 | 62.6% | 115.7 |
| W04 | 2,222 | 816 | 3.2 | 9.8% | 136.1 |
| W05 | 641 | 459 | 14.2 | 3.3% | 68.3 |
| W06 | 6,616 | 2,700 | 0.6 | 83.3% | 112.1 |
| W07 | 79 | 14 | 1.7 | 50.6% | 112.7 |
| W08 | 8,114 | 3,149 | 1.3 | 76.4% | 97.7 |
| W09 | 2,385 | 1,358 | 2.0 | 59.8% | 83.4 |
| W10 | 105 | 55 | 4.5 | 32.4% | 93.5 |
| W11 | 8 | 4 | 0.4 | 75.0% | 139.6 |
| W12 | 1,671 | 414 | 1.4 | 44.6% | 109.9 |
| W13 | 6,572 | 2,250 | 0.9 | 65.1% | 100.0 |
| W14 | 542 | 213 | 1.0 | 70.1% | 138.4 |
| W15 | 5,283 | 1,566 | 0.4 | 80.6% | 105.4 |
| W16 | 2,460 | 1,057 | 2.3 | 34.3% | 123.4 |
| W17 | 591 | 254 | 3.0 | 50.4% | 115.5 |
| W18 | 182 | 84 | 5.2 | 30.8% | 73.2 |
| W19 | 6,968 | 4,829 | 6.0 | 59.9% | 25.9 |
| W20 | 15,417 | 11,699 | 3.8 | 50.0% | 14.4 |
| W21 | 1,142 | 1,020 | 3.8 | 54.9% | 15.3 |
| W22 | 122 | 80 | 7.1 | 67.0% | 13.5 |
| W23 | 6,198 | 5,319 | 3.6 | 49.4% | 21.6 |
| W24 | 6,839 | 4,238 | 7.2 | 74.0% | 6.3 |
| W25 | 404 | 277 | 6.9 | 49.5% | 9.0 |
| W26 | 3,776 | 2,107 | 1.5 | 88.3% | 6.1 |
| W27 | 1,048 | 784 | 1.3 | 39.6% | 96.1 |
| W28 | 29,995 | 13,779 | 1.1 | 79.9% | 82.2 |
| W29 | 46 | 10 | 13.8 | 54.0% | 33.7 |
| W30 | 2,650 | 1,432 | 3.9 | 66.0% | 40.9 |
| W31 | 244 | 108 | 2.0 | 41.4% | 82.2 |
| W32 | 104 | 49 | 4.3 | 35.6% | 89.5 |
| W33 | 1,911 | 1,080 | 4.3 | 62.2% | 55.3 |
| W37 | 29,288 | 10,574 | 5.0 | 0.6% | 114.0 |
| W38 | 29,501 | 10,358 | 3.9 | 0.4% | 108.5 |
| W39 | 1,341 | 1,026 | 6.5 | 1.0% | 117.0 |
| W40 | 82,315 | 26,232 | 3.9 | 0.4% | 118.7 |
| W41 | 2,419 | 844 | 4.1 | 0.4% | 118.0 |
| W42 | 1,826 | 1,145 | 10.2 | 7.1% | 119.4 |
| W43 | 137 | 58 | 4.3 | 19.0% | 142.2 |
| W44 | 1,073 | 326 | 0.4 | 73.6% | 126.0 |
| W45 | 725 | 250 | 0.9 | 74.5% | 129.4 |
| W46 | 291 | 283 | 12.1 | 0.0% | 98.5 |
| W47 | 98 | 55 | 12.8 | 6.1% | 111.1 |
| W48 | 45 | 22 | 12.5 | 8.9% | 123.9 |
| W49 | 245 | 121 | 6.5 | 1.6% | 69.0 |
| W50 | 428 | 128 | 3.1 | 3.3% | 108.7 |
| W51 | 101 | 31 | 4.7 | 3.0% | 107.9 |
| W52 | 110 | 66 | 6.4 | 13.7% | 106.3 |
| W53 | 172 | 47 | 0.9 | 61.1% | 122.3 |
| W54 | 339 | 128 | 0.5 | 80.3% | 129.6 |
| W55 | 786 | 287 | 0.2 | 86.8% | 119.2 |
| W56 | 784 | 277 | 0.3 | 81.6% | 112.2 |
| W57 | 2,663 | 935 | 3.4 | 78.8% | 97.5 |
| W58 | 10,535 | 4,381 | 2.5 | 6.1% | 122.8 |
| W59 | 9,349 | 2,980 | 0.3 | 79.2% | 105.6 |
| W60 | 740 | 312 | 1.7 | 32.2% | 132.8 |
| W61 | 1,029 | 359 | 1.7 | 32.8% | 114.9 |
| W62 | 5,573 | 2,029 | 1.2 | 57.7% | 123.6 |
| W63 | 428 | 164 | 2.5 | 41.1% | 113.5 |
| W64 | 72 | 26 | 6.7 | 34.7% | 108.1 |
| W65 | 1,376 | 1,015 | 3.5 | 50.9% | 9.8 |
| W66 | 1,207 | 821 | 2.5 | 78.1% | 11.2 |
| W67 | 1,146 | 1,113 | 4.5 | 45.8% | 9.1 |
| W68 | 59 | 67 | 1.6 | 67.6% | 6.3 |
| W69 | 2,523 | 1,062 | 1.7 | 79.0% | 82.9 |
| W70 | 123 | 81 | 1.2 | 68.3% | 96.0 |
| W71 | 2,508 | 849 | 0.3 | 81.3% | 99.0 |
| W72 | 165 | 161 | 0.9 | 53.4% | 85.4 |
| W73 | 189 | 72 | 1.0 | 58.2% | 87.3 |
| W74 | 13,849 | 6,838 | 0.6 | 54.1% | 108.1 |
| W75 | 1,571 | 851 | 0.6 | 82.8% | 55.3 |
| W76 | 689 | 240 | 0.3 | 88.4% | 83.3 |
| W77 | 3,713 | 1,698 | 1.0 | 58.2% | 110.4 |
| W78 | 5,189 | 1,793 | 0.4 | 79.5% | 91.8 |
| W79 | 5,337 | 1,500 | 0.2 | 82.5% | 110.8 |
| W80 | 4,786 | 1,771 | 5.0 | 69.7% | 93.0 |
| W81 | 2,265 | 817 | 2.8 | 74.2% | 98.1 |
| W82 | 41,591 | 14,938 | 0.1 | 93.0% | 73.1 |
| W83 | 5,645 | 1,658 | 0.3 | 81.5% | 94.8 |
| W84 | 5,633 | 2,249 | 0.4 | 75.4% | 101.1 |
| W85 | 2,618 | 1,154 | 4.2 | 78.4% | 77.1 |
| W86 | 622 | 257 | 0.5 | 84.9% | 95.1 |
| W87 | 2,469 | 811 | 0.4 | 89.7% | 86.7 |
| W88 | 984 | 346 | 0.4 | 88.7% | 94.7 |
| W89 | 2,518 | 802 | 0.2 | 88.9% | 95.8 |
| W91 | 5,892 | 1,734 | 1.3 | 68.9% | 87.9 |
| W92 | 1,784 | 817 | 0.9 | 90.0% | 64.9 |
| W93 | 2,592 | 922 | 4.3 | 0.4% | 103.8 |
| W94 | 20,787 | 9,304 | 4.1 | 0.4% | 107.6 |
| W95 | 580 | 438 | 5.4 | 0.3% | 129.3 |
| W96 | 1,982 | 625 | 3.4 | 1.5% | 119.4 |
| W97 | 3,439 | 1,166 | 2.9 | 1.2% | 129.0 |
| W98 | 707 | 285 | 4.8 | 1.7% | 123.8 |
| X01 | 14 | 9 | 5.3 | 55.8% | 7.8 |
| X02 | 3 | 0 | 0.6 | 40.0% | 59.5 |
| X03 | 5 | 2 | 3.1 | 80.0% | 144.3 |
| X04 | 1 | 0 | 0.0 | 100.0% | 0.0 |
| X05 | 8 | 3 | 4.3 | 25.0% | 33.8 |
| X07 | 64 | 56 | 11.5 | 3.1% | 40.7 |
| X08 | 2,093 | 1,501 | 1.7 | 84.8% | 46.0 |
| X09 | 1,996 | 1,474 | 21.1 | 0.1% | 48.2 |
| X10 | 740 | 572 | 14.2 | 15.3% | 30.8 |
| X11 | 4,701 | 2,668 | 7.0 | 49.0% | 48.1 |
| X12 | 997 | 514 | 9.0 | 37.5% | 66.3 |
| X14 | 671 | 643 | 13.9 | 0.1% | 29.2 |
| X15 | 287 | 152 | 6.6 | 0.0% | 161.7 |
| X16 | 52 | 24 | 1.1 | 32.7% | 81.2 |
| X17 | 0 | 0 | 3.0 | 0.0% | 0.0 |
| X19 | 22 | 18 | 2.0 | 4.5% | 122.4 |
| X20 | 11 | 6 | 1.0 | 54.5% | 89.1 |
| X21 | 1,276 | 657 | 0.4 | 91.3% | 98.0 |
| X22 | 1,013 | 623 | 3.9 | 5.8% | 113.7 |
| X23 | 28 | 20 | 2.5 | 28.6% | 133.4 |
| X24 | 147 | 81 | 1.4 | 20.4% | 137.6 |
| X25 | 385 | 169 | 1.1 | 34.3% | 128.6 |
| X27 | 481 | 175 | 0.2 | 91.7% | 134.4 |
| X45 | 696 | 314 | 3.6 | 0.6% | 33.4 |
| X46 | 136 | 74 | 1.3 | 32.4% | 19.9 |
| X55 | 1,010 | 948 | 2.7 | 72.5% | 11.8 |

AHES-APC: Annual Hospital Episode Statistics Hospital Admitted Patient Care Activity
The number of elective procedures was estimated from raw AHES-APC data as described in the methods

**Supplementary Table 3:** Procedural costs mapped to Health Resource Groups

| Procedure | Health Resource Group | Cost per procedure |
| --- | --- | --- |
| **Breast surgery** |  |  |
| Excision of breast | Malignant Breast Disorders with Interventions, with CC Score 0-2 | £3,724 |
| Other daycase breast surgery | Non-Malignant Breast Disorders with Interventions | £27,863 |
| Other inpatient breast surgery | Non-Malignant Breast Disorders with Interventions | £27,863 |
| **Cardiac surgery** |  |  |
| Coronary artery surgery | Standard Coronary Artery Bypass Graft with CC Score 0-4 | £11,292 |
| Other daycase cardiac surgery | Standard, Other Operations on Heart or Pericardium, with CC Score 0-4 | £7,612 |
| Other inpatient cardiac surgery | Standard, Other Operations on Heart or Pericardium, with CC Score 0-4 | £7,612 |
| Surgery on the great vessels | Standard Repair of Aortic Root with CC Score 0-6 | £14,559 |
| Valvular heart surgery | Standard, Single Heart Valve Replacement or Repair, with CC Score 0-5 | £12,082 |
| **Cardiology** |  |  |
| Interventional cardiology | Standard Percutaneous Transluminal Coronary Angioplasty with CC Score 0-3 | £2,885 |
| **Colorectal surgery** |  |  |
| Colorectal resection | Malignant Gastrointestinal Tract Disorders with Single Intervention, with CC Score 0-2 | £3,544 |
| Ileostomy, colostomy | Non-Malignant Gastrointestinal Tract Disorders with Single Intervention, with CC Score 0-2 | £3,710 |
| Other daycase colorectal surgery | Intermediate Anal Procedures, 19 years and over, with CC Score 1-2 | £2,200 |
| Other inpatient colorectal surgery | Non-Malignant Gastrointestinal Tract Disorders with Single Intervention, with CC Score 0-2 | £3,710 |
| Proctology | Minor Anal Procedures | £1,728 |
| Sigmoidoscopy, colonoscopy | Diagnostic Colonoscopy with Biopsy, 19 years and over | £1,176 |
| **General surgery** |  |  |
| Diagnostic laparoscopy | Intermediate Therapeutic General Abdominal Procedures, 19 years and over, with CC Score 1-2 | £4,237 |
| Excision of lymph node | Minor Therapeutic or Diagnostic, General Abdominal Procedures, 19 years and over | £1,392 |
| Excision of thyroid, parathyroid | Thyroid Procedures with CC Score 0-1 | £4,515 |
| Groin hernia repair | Inguinal, Umbilical or Femoral Hernia Procedures, 19 years and over, with CC Score 1-2 | £3,132 |
| Other daycase endocrine surgery | Thyroid Procedures with CC Score 0-1 | £4,515 |
| Other daycase general surgery | Minor Therapeutic or Diagnostic, General Abdominal Procedures, 19 years and over | £1,392 |
| Other inpatient endocrine surgery | Adrenal Procedures with CC Score 0-1 | £6,123 |
| Other inpatient general surgery | Major General Abdominal Procedures, 19 years and over, with CC Score 1-2 | £5,785 |
| Other interventional radiology | Percutaneous Single Drainage of Abdominal Abscess, with CC Score 0-1 | £2,285 |
| Ventral hernia repair | Abdominal Hernia Procedures, 19 years and over, with CC Score 1-3 | £4,725 |
| **Gynaecology** |  |  |
| Excision of vulva, vagina, cervix | Major Open Lower Genital Tract Procedures with CC Score 0-2 | £3,484 |
| Female sterilisation | Minor, Laparoscopic or Endoscopic, Upper Genital Tract Procedures | £2,624 |
| Hysterectomy, salpingoophorectomy | Major Open Upper Genital Tract Procedures with CC Score 0-2 | £4,897 |
| Hysteroscopy & related procedures | Diagnostic Hysteroscopy | £1,853 |
| Other daycase gynaecology | Minor Lower Genital Tract Procedures | £2,002 |
| Other inpatient gynaecology | Intermediate, Laparoscopic or Endoscopic, Upper Genital Tract Procedures, with CC Score 0-1 | £3,194 |
| Ovarian surgery | Major, Laparoscopic or Endoscopic, Upper Genital Tract Procedures, with CC Score 0-1 | £4,507 |
| Repair of prolapse of vagina | Intermediate Open Lower Genital Tract Procedures with CC Score 0-2 | £2,829 |
| **Head & Neck surgery** |  |  |
| Adenoidectomy | Adenoidectomy | £1,961 |
| Laryngoscopy | Diagnostic, Laryngoscopy or Pharyngoscopy, 19 years and over | £1,854 |
| Maxillofacial surgery | Major Maxillofacial Procedures, 19 years and over, with CC Score 1+ | £6,101 |
| Operations on ear | Intermediate Ear Procedures, 19 years and over | £3,102 |
| Operations on larynx, pharynx | Major, Mouth or Throat Procedures, 19 years and over, with CC Score 0-1 | £3,818 |
| Operations on nose | Intermediate Nose Procedures | £2,666 |
| Oral surgery | Intermediate, Mouth or Throat Procedures, 19 years and over, with CC Score 0-1 | £2,743 |
| Tonsillectomy | Tonsillectomy, 4 years and over | £2,183 |
| **Hepatobiliary surgery** |  |  |
| Cholecystectomy | Laparoscopic Cholecystectomy, 19 years and over, with CC Score 1-3 | £3,939 |
| Diagnostic percutaneous operations on liver | Non-Malignant, Hepatobiliary or Pancreatic Disorders, with Single Intervention, with CC Score 0-3 | £3,195 |
| ERCP & related procedures | Intermediate Therapeutic Endoscopic Retrograde Cholangiopancreatography with CC Score 0-1 | £1,947 |
| Excision of liver, pancreas, spleen | Very Major, Hepatobiliary or Pancreatic Procedures, with CC Score 0-2 | £8,088 |
| Other daycase hepatobiliary surgery | Minor, Hepatobiliary or Pancreatic Procedures, with CC Score 1+ | £4,603 |
| Other inpatient hepatobiliary surgery | Very Major, Hepatobiliary or Pancreatic Procedures, with CC Score 0-2 | £8,088 |
| Other interventional radiology | Other Percutaneous Therapeutic, Hepatobiliary or Pancreatic Procedures, with CC Score 0-2 | £2,772 |
| **Neurosurgery** |  |  |
| Brain tissue surgery | Complex Intracranial Procedures, 18 years and under, with CC Score 0-3 | £12,140 |
| Brain ventricular surgery | Complex Intracranial Procedures, 18 years and under, with CC Score 0-3 | £12,140 |
| Cranial nerve surgery | Very Major Intracranial Procedures, 18 years and under, with CC Score 0-3 | £7,233 |
| Cranium and dural surgery | Very Major Intracranial Procedures, 18 years and under, with CC Score 0-3 | £7,233 |
| Neurovascular interventional radiology | Percutaneous Transluminal Embolisation of, Single Small or Medium, Intracranial or Extracranial Aneurysm, with CC Score 0-3 | £5,568 |
| Other inpatient neurosurgery | Major Intracranial Procedures, 18 years and under, with CC Score 0-3 | £8,188 |
| **Oesophagogastric surgery** |  |  |
| Gastrosocopy | Diagnostic Endoscopic Upper Gastrointestinal Tract Procedures, 19 years and over | £999 |
| Gastrostomy, jejunostomy | Radiological Insertion of Gastrostomy Tube, 19 years and over | £1,496 |
| Oesophagogastric resection | Complex, Oesophageal, Stomach or Duodenum Procedures, 19 years and over, with CC Score 0-1 | £6,840 |
| Other daycase oesophagogastric surgery | Intermediate Upper Gastrointestinal Tract Procedures, 19 years and over | £1,464 |
| Other inpatient oesophagogastric surgery | Major, Oesophageal, Stomach or Duodenum Procedures, 19 years and over, with CC Score 0-1 | £5,091 |
| **Ophthalmology** |  |  |
| Cataract surgery | Intermediate, Cataract or Lens Procedures, with CC Score 0-1 | £3,557 |
| Glaucoma surgery | Minor, Glaucoma or Iris Procedures | £1,432 |
| Operations on vitreous body of eye | Intermediate Vitreous Retinal Procedures, 19 years and over, with CC Score 0-1 | £1,320 |
| Other daycase ophthalmology | Intermediate Oculoplastics Procedures, 19 years and over, with CC Score 0-1 | £2,087 |
| Other inpatient ophthalmology | Major, Orbit or Lacrimal Procedures, 19 years and over, with CC Score 1+ | £3,096 |
| **Orthopaedics** |  |  |
| Arthroscopic procedures | Minor Knee Procedures for Non-Trauma, 19 years and over | £2,262 |
| Daycase procedures on joint | General orthopaedics (non-trauma)* | £3,960 |
| Decompression of cervical spine | Percutaneous Vertebroplasty of One Level of Spine | £3,697 |
| Decompression of lumbar spine | Percutaneous Vertebroplasty of One Level of Spine | £3,697 |
| Denervation of spinal facet joint | Denervation or Injection around Spinal Facet, for Pain Management | £916 |
| Division or excision of bone | Malignancy, of Bone or Connective Tissue, with CC Score 0-1 | £2,179 |
| Excision of ganglion | Soft Tissue Disorders with CC Score 0-2 | £1,001 |
| Fasciectomy, fasciotomy | Soft Tissue Disorders with CC Score 0-2 | £1,001 |
| Fracture-related surgery | General orthopaedics (trauma)* | £3,710 |
| Fusion of joint | General orthopaedics (non-trauma)* | £3,960 |
| Fusion of joint of toe | Major Foot Procedures for Non-Trauma, 19 years and over, with CC Score 0-1 | £4,975 |
| Lower limb joint replacement | Complex, Hip or Knee Procedures for Non-Trauma, with CC Score 0-1 | £9,775 |
| Operations on bursa | Minor Knee Procedures for Non-Trauma, 19 years and over | £2,262 |
| Other daycase orthopaedics | General orthopaedics (non-trauma)* | £3,960 |
| Other daycase spinal surgery | Intermediate Extradural Spinal Procedures with CC Score 0-1 | £4,884 |
| Other inpatient orthopaedics | General orthopaedics (non-trauma)* | £3,960 |
| Other inpatient spinal surgery | Major Extradural Spinal Procedures with CC Score 0-1 | £5,667 |
| Other reconstruction of joint | General orthopaedics (non-trauma)* | £3,960 |
| Procedures on nail, nail bed | Soft Tissue Disorders with CC Score 0-2 | £1,001 |
| Procedures on peripheral nerves | Minor Hand Procedures for Non-Trauma, 19 years and over | £2,052 |
| Reconstruction of foot | Major Foot Procedures for Trauma, 19 years and over, with CC Score 1 | £4,877 |
| Spinal fusion | Major Spinal Reconstructive Procedures with CC Score 0-1 | £11,154 |
| Surgery on ligament, tendon, muscle | Other Muscle, Tendon, Fascia or Ligament Procedures | £4,844 |
| Upper limb joint replacement | Complex, Foot, Hand, Shoulder or Elbow Procedures for Non-Trauma, with CC Score 0-1 | £7,942 |
| **Plastic surgery** |  |  |
| Excision of lesion of skin | Minor Skin Procedures, 19 years and over | £1,400 |
| Other daycase plastic surgery | Intermediate Skin Procedures, 19 years and over | £2,157 |
| Other inpatient plastic surgery | Major Skin Procedures | £7,680 |
| Procedure on eyelid | Minor Oculoplastics Procedures, 19 years and over | £1,220 |
| Procedure on lip | Minor, Mouth or Throat Procedures, 19 years and over | £2,063 |
| **Thoracic surgery** |  |  |
| Biopsy of lung | Minor Thoracic Procedures | £2,175 |
| Bronchoscopy | Diagnostic Bronchoscopy, 19 years and over | £2,054 |
| Lung resection | Major Thoracic Procedures, 19 years and over, with CC Score 0-2 | £5,424 |
| Other daycase thoracic surgery | Minor Thoracic Procedures | £2,175 |
| Other inpatient thoracic surgery | Intermediate Thoracic Procedures, 19 years and over, with CC Score 0-2 | £3,999 |
| **Urology** |  |  |
| Cystectomy | Cystectomy with Urinary Diversion and Reconstruction, with CC Score 0-2 | £11,425 |
| Extracorporeal fragmentation of calculus of kidney | Urinary Tract Stone Disease with Interventions, with CC Score 0-2 | £3,836 |
| Male sterilisation | Minor, Scrotum, Testis or Vas Deferens Procedures, 19 years and over | £2,368 |
| Nephrectomy, uretectomy | Major Laparoscopic, Kidney or Ureter Procedures, 19 years and over, with CC Score 0-2 | £6,902 |
| Operations on bladder | Intermediate Open Bladder Procedures | £4,577 |
| Other daycase urology | Ureteric or Bladder Disorders, with Interventions, with CC Score 0-3 | £2,723 |
| Other inpatient urology | Ureteric or Bladder Disorders, with Interventions, with CC Score 0-3 | £2,723 |
| Other interventional radiology | Unilateral, Percutaneous Insertion of, Ureteric Stent or Nephrostomy | £1,634 |
| Percutaneous puncture of kidney | Other Percutaneous Therapeutic, Hepatobiliary or Pancreatic Procedures, with CC Score 0-2 | £2,772 |
| Prepuceplasty | Minor Penis Procedures, 19 years and over | £2,171 |
| Prostatectomy | Transurethral Prostate Resection Procedures with CC Score 0-2 | £3,420 |
| Scrotal surgery | Scrotum, Testis or Vas Deferens Disorders, with Interventions, with CC Score 0-1 | £6,474 |
| Transplant surgery | Kidney Transplant, 19 years and over, from Live Donor | £12,840 |
| Ureteric stent procedures | Unilateral, Percutaneous Insertion of, Ureteric Stent or Nephrostomy | £1,634 |
| Ureteroscopy, cystoscopy & related procedures | Intermediate Endoscopic Ureter Procedures, 19 years and over | £2,115 |
| Urethral surgery | Minor or Intermediate, Urethra Procedures, 19 years and over | £1,723 |
| **Vascular surgery** |  |  |
| Amputation of lower limb | Amputation of Single Limb with CC Score 0-9 | £8,613 |
| Amputation of upper limb | Amputation of Single Limb with CC Score 0-9 | £8,613 |
| Aortic surgery | Single Open Procedure, on Aorta or Abdominal Blood Vessel, with CC Score 0-3 | £8,526 |
| Arteriovenous shunt | Open Arteriovenous Fistula, Graft or Shunt Procedures | £3,261 |
| Other daycase vascular surgery | Open Operations, on Other or Unspecified Blood Vessels, with CC Score 0-1 | £3,780 |
| Other inpatient vascular surgery | Open Operations, on Other or Unspecified Blood Vessels, with CC Score 0-1 | £3,780 |
| Surgery on arteries | Single Open Procedure on Blood Vessel of Lower Limb with CC Score 0-3 | £6,968 |
| Varicose vein surgery | Open Treatment of Primary Unilateral Varicose Veins | £2,669 |
| Vascular interventional radiology | Percutaneous Transluminal Angioplasty of Single Blood Vessel with CC Score 0-2 | £1,757 |

Costs are based on NHS reference costs for 2019-20
\*Health Resource Group (HRG) categorisation is more granular than some of our orthopaedic procedure categories, making it difficult to match our procedures to a single HRG code. Therefore, for non-trauma procedures we took an average cost for the following HRG codes: Intermediate Hip Procedures for Non-Trauma, 19 years and over, with CC Score 1, Intermediate Knee Procedures for Non-Trauma, between 6 and 18 years, with CC Score 1+, Intermediate Foot Procedures for Non-Trauma, 19 years and over, with CC Score 0-1, Intermediate Hand Procedures for Non-Trauma, 19 years and over, with CC Score 0-1, Intermediate Shoulder Procedures for Non-Trauma, 19 years and over, with CC Score 0-1, Intermediate Elbow Procedures for Non-Trauma, 19 years and over, with CC Score 0-1, resulting in an estimated cost per procedure of £3,960. Equivalent trauma-related codes were used to calculate an average cost (£3,710) for trauma procedures.

**Supplementary Table 4:** Results of expert survey

| **Specialty of respondents** |  |
| --- | --- |
| Anaesthetics | 2 |
| Breast surgery | 1 |
| Cardiac surgery | 3 |
| Colorectal surgery | 5 |
| General surgery | 9 |
| Gynaecology | 3 |
| Head & Neck surgery | 3 |
| Hepatobiliary surgery | 3 |
| Neurosurgery | 3 |
| Ophthalmology | 1 |
| Orthopaedics | 9 |
| Plastic surgery | 1 |
| Thoracic surgery | 1 |
| Urology | 2 |
| Vascular surgery | 1 |
| **Estimate for when elective procedures will return to pre-pandemic baseline volume** |  |
| Already at or above pre-pandemic baseline | 12 |
| July 2022 | 5 |
| December 2022 | 6 |
| July 2023 | 8 |
| December 2023 | 4 |
| July 2024 | 3 |
| December 2024 | 1 |
| July 2025 | 1 |
| December 2025 | 4 |
| July 2026 - July 2029 | 0 |
| December 2029 or later | 1 |
| Unlikely to ever return to pre-pandemic baseline | 2 |
| **Estimates for realistic annual increase in elective procedure volume after it has returned to its pre-pandemic baseline** |  |
| Increase above pre-pandemic baseline will not be possible | 22 |
| Up to 2.4% increase per year | 9 |
| 2.5-4.9% increase per year | 7 |
| 5.0-7.4% increase per year | 2 |
| 7.5-9.9% increase per year | 2 |
| 10.0-12.4% increase per year | 2 |
| 12.5-19.9% increase per year | 0 |
| 20.0% or greater increase per year | 3 |

Responses collected through anonymous online survey on 5-7 June 2022. Survey instrument is available in Supplement 1.

**Supplemental Table 5:** Projections for annual incident need for elective procedures in 2022-29

| **Period** | **Annual incident need**<br>**for elective procedures** |
| --- | --- |
| April – December 2022 | 4,480,571 |
| January – December 2023 | 6,038,213 |
| January – December 2024 | 6,099,468 |
| January – December 2025 | 6,157,013 |
| January – December 2026 | 6,215,214 |
| January – December 2027 | 6,270,663 |
| January – December 2028 | 6,323,103 |
| January – December 2029 | 6,372,834 |

**Supplemental Table 6:** Forecasts for total need for elective procedures in England in different scenarios

| Month | Current capacity | Pessimistic scenario | Central scenario | Optimistic scenario |
| --- | --- | --- | --- | --- |
| Mar-22 | 4,347,469 | 4,347,469 | 4,347,469 | 4,347,469 |
| Apr-22 | 4,439,973 | 4,435,513 | 4,432,044 | 4,432,044 |
| May-22 | 4,532,477 | 4,519,098 | 4,508,691 | 4,508,691 |
| Jun-22 | 4,624,981 | 4,598,222 | 4,577,409 | 4,577,409 |
| Jul-22 | 4,717,485 | 4,672,887 | 4,638,199 | 4,638,199 |
| Aug-22 | 4,809,989 | 4,743,091 | 4,691,060 | 4,691,060 |
| Sep-22 | 4,902,494 | 4,808,836 | 4,735,992 | 4,735,992 |
| Oct-22 | 4,994,998 | 4,870,121 | 4,772,995 | 4,772,995 |
| Nov-22 | 5,087,502 | 4,926,946 | 4,802,070 | 4,802,070 |
| Dec-22 | 5,180,006 | 4,979,312 | 4,823,216 | 4,823,216 |
| Jan-23 | 5,277,853 | 5,032,560 | 4,848,911 | 4,848,116 |
| Feb-23 | 5,375,700 | 5,081,349 | 4,873,811 | 4,871,428 |
| Mar-23 | 5,473,548 | 5,125,678 | 4,897,917 | 4,893,150 |
| Apr-23 | 5,571,395 | 5,165,547 | 4,921,228 | 4,913,283 |
| May-23 | 5,669,242 | 5,200,956 | 4,943,745 | 4,931,828 |
| Jun-23 | 5,767,090 | 5,231,905 | 4,965,468 | 4,948,783 |
| Jul-23 | 5,864,937 | 5,258,394 | 4,986,395 | 4,964,150 |
| Aug-23 | 5,962,784 | 5,284,883 | 5,006,529 | 4,977,927 |
| Sep-23 | 6,060,632 | 5,311,373 | 5,025,868 | 4,990,116 |
| Oct-23 | 6,158,479 | 5,337,862 | 5,044,412 | 5,000,715 |
| Nov-23 | 6,256,326 | 5,364,351 | 5,062,162 | 5,009,726 |
| Dec-23 | 6,354,174 | 5,390,841 | 5,079,117 | 5,017,147 |
| Jan-24 | 6,457,125 | 5,422,435 | 5,100,383 | 5,028,084 |
| Feb-24 | 6,560,077 | 5,454,029 | 5,120,854 | 5,037,432 |
| Mar-24 | 6,663,029 | 5,485,622 | 5,140,530 | 5,045,191 |
| Apr-24 | 6,765,981 | 5,517,216 | 5,159,413 | 5,051,362 |
| May-24 | 6,868,933 | 5,548,810 | 5,177,500 | 5,055,943 |
| Jun-24 | 6,971,885 | 5,580,404 | 5,194,793 | 5,058,935 |
| Jul-24 | 7,074,837 | 5,611,998 | 5,211,292 | 5,060,338 |
| Aug-24 | 7,177,788 | 5,643,592 | 5,226,996 | 5,060,152 |
| Sep-24 | 7,280,740 | 5,675,186 | 5,241,905 | 5,058,378 |
| Oct-24 | 7,383,692 | 5,706,780 | 5,256,020 | 5,055,014 |
| Nov-24 | 7,486,644 | 5,738,374 | 5,269,341 | 5,050,061 |
| Dec-24 | 7,589,596 | 5,769,967 | 5,281,867 | 5,043,519 |
| Jan-25 | 7,697,343 | 5,806,357 | 5,298,394 | 5,040,184 |
| Feb-25 | 7,805,090 | 5,842,746 | 5,314,126 | 5,035,260 |
| Mar-25 | 7,912,838 | 5,879,135 | 5,329,064 | 5,028,747 |
| Apr-25 | 8,020,585 | 5,915,525 | 5,343,208 | 5,020,644 |
| May-25 | 8,128,332 | 5,951,914 | 5,356,557 | 5,010,953 |
| Jun-25 | 8,236,079 | 5,988,303 | 5,369,112 | 4,999,673 |
| Jul-25 | 8,343,827 | 6,024,692 | 5,380,872 | 4,986,804 |
| Aug-25 | 8,451,574 | 6,061,082 | 5,391,837 | 4,972,346 |
| Sep-25 | 8,559,321 | 6,097,471 | 5,402,008 | 4,956,298 |
| Oct-25 | 8,667,069 | 6,133,860 | 5,411,385 | 4,938,662 |
| Nov-25 | 8,774,816 | 6,170,250 | 5,419,967 | 4,919,437 |
| Dec-25 | 8,882,563 | 6,206,639 | 5,427,754 | 4,898,623 |
| Jan-26 | 8,995,160 | 6,247,878 | 5,439,598 | 4,881,070 |
| Feb-26 | 9,107,758 | 6,289,118 | 5,450,646 | 4,861,928 |
| Mar-26 | 9,220,355 | 6,330,357 | 5,460,901 | 4,841,197 |
| Apr-26 | 9,332,953 | 6,371,596 | 5,470,360 | 4,818,877 |
| May-26 | 9,445,550 | 6,412,836 | 5,479,025 | 4,794,968 |
| Jun-26 | 9,558,147 | 6,454,075 | 5,486,896 | 4,769,470 |
| Jul-26 | 9,670,745 | 6,495,314 | 5,493,972 | 4,742,383 |
| Aug-26 | 9,783,342 | 6,536,554 | 5,500,254 | 4,713,707 |
| Sep-26 | 9,895,939 | 6,577,793 | 5,505,741 | 4,683,443 |
| Oct-26 | 10,008,537 | 6,619,033 | 5,510,434 | 4,651,589 |
| Nov-26 | 10,121,134 | 6,660,272 | 5,514,332 | 4,618,146 |
| Dec-26 | 10,233,731 | 6,701,511 | 5,517,436 | 4,583,114 |
| Jan-27 | 10,350,950 | 6,747,372 | 5,524,366 | 4,551,114 |
| Feb-27 | 10,468,168 | 6,793,232 | 5,530,502 | 4,517,525 |
| Mar-27 | 10,585,386 | 6,839,092 | 5,535,843 | 4,482,347 |
| Apr-27 | 10,702,604 | 6,884,952 | 5,540,390 | 4,445,580 |
| May-27 | 10,819,822 | 6,930,812 | 5,544,142 | 4,407,224 |
| Jun-27 | 10,937,040 | 6,976,672 | 5,547,099 | 4,367,279 |
| Jul-27 | 11,054,258 | 7,022,532 | 5,549,262 | 4,325,745 |
| Aug-27 | 11,171,476 | 7,068,393 | 5,550,631 | 4,282,622 |
| Sep-27 | 11,288,695 | 7,114,253 | 5,551,205 | 4,237,910 |
| Oct-27 | 11,405,913 | 7,160,113 | 5,550,985 | 4,191,609 |
| Nov-27 | 11,523,131 | 7,205,973 | 5,549,970 | 4,143,720 |
| Dec-27 | 11,640,349 | 7,251,833 | 5,548,160 | 4,094,241 |
| Jan-28 | 11,761,937 | 7,302,063 | 5,549,927 | 4,047,543 |
| Feb-28 | 11,883,525 | 7,352,293 | 5,550,898 | 3,999,256 |
| Mar-28 | 12,005,113 | 7,402,524 | 5,551,075 | 3,949,380 |
| Apr-28 | 12,126,701 | 7,452,754 | 5,550,458 | 3,897,915 |
| May-28 | 12,248,289 | 7,502,984 | 5,549,046 | 3,844,862 |
| Jun-28 | 12,369,878 | 7,553,214 | 5,546,840 | 3,790,219 |
| Jul-28 | 12,491,466 | 7,603,444 | 5,543,839 | 3,733,987 |
| Aug-28 | 12,613,054 | 7,653,674 | 5,540,044 | 3,676,166 |
| Sep-28 | 12,734,642 | 7,703,904 | 5,535,454 | 3,616,757 |
| Oct-28 | 12,856,230 | 7,754,134 | 5,530,070 | 3,555,758 |
| Nov-28 | 12,977,818 | 7,804,365 | 5,523,891 | 3,493,170 |
| Dec-28 | 13,099,406 | 7,854,595 | 5,516,918 | 3,428,994 |
| Jan-29 | 13,225,139 | 7,908,969 | 5,513,294 | 3,367,372 |
| Feb-29 | 13,350,871 | 7,963,343 | 5,508,876 | 3,304,162 |
| Mar-29 | 13,476,603 | 8,017,718 | 5,503,664 | 3,239,362 |
| Apr-29 | 13,602,336 | 8,072,092 | 5,497,657 | 3,172,974 |
| May-29 | 13,728,068 | 8,126,467 | 5,490,855 | 3,104,997 |
| Jun-29 | 13,853,801 | 8,180,841 | 5,483,259 | 3,035,430 |
| Jul-29 | 13,979,533 | 8,235,215 | 5,474,869 | 2,964,275 |
| Aug-29 | 14,105,265 | 8,289,590 | 5,465,684 | 2,891,531 |
| Sep-29 | 14,230,998 | 8,343,964 | 5,455,704 | 2,817,198 |
| Oct-29 | 14,356,730 | 8,398,339 | 5,444,931 | 2,741,275 |
| Nov-29 | 14,482,462 | 8,452,713 | 5,433,362 | 2,663,764 |
| Dec-29 | 14,608,195 | 8,507,087 | 5,420,999 | 2,584,664 |

## References

1. NHS England. Understanding Winter Pressures in A&E Departments. Accessed online on 14 June 2022 at https://www.england.nhs.uk/wp-content/uploads/2013/11/wint-press-rep.pdf.

2. NHS England. Cancelled Elective Operations Data. Accessed online on 14 June 2022 at https://www.england.nhs.uk/statistics/statistical-work-areas/cancelled-elective-operations/cancelled-ops-data/

3. COVIDSurg Collaborative. Global guidance for surgical care during the COVID-19 pandemic. Br J Surg. 2020 Aug;107(9):1097–1103.

4. COVIDSurg Collaborative. Elective surgery cancellations due to the COVID-19 pandemic: global predictive modelling to inform surgical recovery plans. Br J Surg. 2020 Oct;107(11):1440–1449.

5. Gregory A, Campbell D (2022, Apr 14). Covid disruption to NHS in England wreaks havoc with surgery backlog. The Guardian. Accessed online on 20 June 2022 at https://www.theguardian.com/society/2022/apr/14/covid-disruption-to-nhs-in-england-wreaks-havoc-with-surgery-backlog

6. COVIDSurg Collaborative. Projecting COVID-19 disruption to elective surgery. Lancet. 2022 Jan 15;399(10321):233–234.

7. NHS England. Consultant-led Referral to Treatment Waiting Times. Accessed online on 1 June 2022 at https://www.england.nhs.uk/statistics/statistical-work-areas/rtt-waiting-times/

8. NHS England (2022). Delivery plan for tackling the COVID-19 backlog of elective care. Accessed online on 11 June 2022 at https://www.england.nhs.uk/coronavirus/wp-content/uploads/sites/52/2022/02/C1466-delivery-plan-for-tackling-the-covid-19-backlog-of-elective-care.pdf

9. NHS Confederation. Reaching out to 7 million patients on ‘hidden’ waiting list requires a generational change. Accessed online on 20 June 2022 at https://www.nhsconfed.org/news/reaching-out-7-million-patients-hidden-waiting-list-requires-generational-change

10. COVIDSurg Collaborative, GlobalSurg Collaborative. Timing of surgery following SARS-CoV-2 infection: country income analysis. Anaesthesia. 2022 Jan;77(1):111–112.

11. NHS England. Hospital Episode Statistics (HES). Accessed online on 20 May 2022 at https://digital.nhs.uk/data-and-information/data-tools-and-services/data-services/hospital-episode-statistics

12. NHS England. National Cost Collection for the NHS. Accessed online on 20 May 2022 at https://www.england.nhs.uk/costing-in-the-nhs/national-cost-collection/

13. Office for National Statistics. Estimates of the population for the UK, England and Wales, Scotland and Northern Ireland. Accessed online on 20 May 2022 at https://www.ons.gov.uk/peoplepopulationandcommunity/populationandmigration/populationestimates/datasets/populationestimatesforukenglandandwalesscotlandandnorthernireland

14. Office for National Statistics. Principal projection - England population in age groups. Accessed online on 20 May 2022 at https://www.ons.gov.uk/peoplepopulationandcommunity/populationandmigration/populationprojections/datasets/tablea24principalprojectionenglandpopulationinagegroups

15. Office for National Statistics. Clinical commissioning group population estimates (National Statistics). Accessed online on 20 May 2022 at https://www.ons.gov.uk/peoplepopulationandcommunity/populationandmigration/populationestimates/datasets/clinicalcommissioninggroupmidyearpopulationestimates

16. Public Health England (2021). Health Equity Assessment Tool (HEAT): executive summary. https://www.gov.uk/government/publications/health-equity-assessment-tool-heat/health-equity-assessment-tool-heat-executive-summary

17. Getting it Right First Time (2021). Elective Recovery High Volume Low Complexity (HVLC) guide for systems. Accessed online on 27 May 2022 at https://www.gettingitrightfirsttime.co.uk/wp-content/uploads/2021/05/GIRFT-HVLC-Guide-Final-V6.pdf

18. British Medical Association. NHS backlog data analysis. Accessed online on 10 June 2022 at https://www.bma.org.uk/advice-and-support/nhs-delivery-and-workforce/pressures/nhs-backlog-data-analysis

19. Stoye G, Warner M, Zaranko B. The NHS backlog recovery plan and the outlook for waiting lists. Accessed online on 10 June 2022 at https://ifs.org.uk/publications/15941

20. Gardner T, Fraser C. Elective care: how has COVID-19 affected the waiting list? Accessed online on 10 June 2022 at https://www.health.org.uk/news-and-comment/charts-and-infographics/elective-care-how-has-covid-19-affected-the-waiting-list

