## Supplement 1 (Expert survey) for "Forecasting waiting lists for elective procedures and surgery in England: a modelling study"

### Forecasting the NHS waiting list in England: expert survey

We are currently compling a report "Forecasting the NHS waiting list in England: 2022-2030" which will be released in mid June 2022.

This report will include projections for the possible size of the NHS waiting lists through to 2030.

We would like to ask for your help please to inform the parameters in our projections. We would be very grateful if you please complete the short, anonymous survey below.

Dmitri Nepogodiev / Aneel Bhangu

#### Your practice

1) Your country

This survey is aimed at UK teams

- ◯ England
- ◯ Northern Ireland
- ◯ Scotland
- ◯ Wales

2) Your role

This survey is aimed at registrars/ consultants

- ◯ Consultant / equivalent
- ◯ Registrar / equivalent

3) Your specialty

Please select the best match

- ◯ Anaesthetics
- ◯ Breast surgery
- ◯ Cardiac surgery
- ◯ Colorectal surgery
- ◯ Endocrine surgery
- ◯ General surgery
- ◯ Gynaecology
- ◯ Hepatobiliary surgery
- ◯ Neurosurgery
- ◯ Oesophagogastric surgery
- ◯ Ophthalmology
- ◯ Oral and maxillofacial surgery
- ◯ Orthopaedics
- ◯ Otolaryngology (head and neck surgery)
- ◯ Paediatric surgery
- ◯ Plastic surgery
- ◯ Thoracic surgery
- ◯ Transplant surgery
- ◯ Urology
- ◯ Vascular surgery
- ◯ Other

#### Parameters for projections

In England, although the overall number of elective procedures performed each month has been steadily increasing, in March 2022 elective procedural activity was still approximately 15% lower than it was pre-pandemic.

4) When do you think your unit will return to its pre-pandemic baseline for ELECTIVE procedural activity?

Pre-pandemic baseline means the same number of procedures performed per month as before the pandemic

- ◯ Already at or above pre-pandemic baseline
- ◯ July 2022
- ◯ December 2022
- ◯ July 2023
- ◯ December 2023
- ◯ July 2024
- ◯ December 2024
- ◯ July 2025
- ◯ December 2025
- ◯ July 2026
- ◯ December 2026
- ◯ July 2027
- ◯ December 2027
- ◯ July 2028
- ◯ December 2028
- ◯ July 2029
- ◯ December 2029 or later
- ◯ Unlikely to ever return to pre-pandemic baseline

5) Once your unit has returned to it's pre-pandemic baseline for elective procedural volume, what do you consider a realistic target for increasing annual ELECTIVE procedural volume, as a proportion of pre-pandemic volume?

i.e. if prepandemic elective procedural volume was 1,000, a 2% increase would mean that each year the number of elective procedures would increase by 20 (2% of 1,000). So volume in year 1 would be 1,020, in year 2 volume would be 1,040, in year 3 volume would be 1,060 etc.

- ◯ Increase above pre-pandemic baseline will not be possible
- ◯ Up to 2.4% increase per year
- ◯ 2.5-4.9% increase per year
- ◯ 5.0-7.4% increase per year
- ◯ 7.5-9.9% increase per year
- ◯ 10.0-12.4% increase per year
- ◯ 12.5-14.9% increase per year
- ◯ 15.0-17.4% increase per year
- ◯ 17.5-19.9% increase per year
- ◯ 20.0% or greater increase per year

Once you have completed all the fields above, please press 'submit'.

Thank you very much for your support.
